# Global Levels and Trends in Child Discipline: Evidence from 88 Countries, 2005–2023

**DOI:** 10.64898/2026.02.13.26346262

**Authors:** John Egyir, Elisabetta De Cao, Katherina Thomas, Elisabetta Aurino

**Affiliations:** Universitat de Barcelona, Spain; Università di Bologna, Department of Economics, Italy; Institut d’Economia de Barcelona, Spain; Organisation for Economic Co-operation and Development

## Abstract

**Background:** Home disciplinary practices shape children’s health and development. Yet, comprehensive, up-to-date global evidence on their levels, trends, and socioeconomic and regional inequalities remains limited. This study provides the first global prevalence estimates of both violent and non-violent forms of discipline, examining regional disparities, variations by child and family characteristics, and changes over time.

**Methods:** We drew from 176 nationally-representative Multiple Indicator Cluster Surveys and Demographic and Health Surveys, collected between 2005 and 2023 across 83 low- and middle-income and 5 high-income countries (N= 1,544,000 1-14y-olds). We estimated weighted prevalence estimates for all types of discipline (exclusively or only non-violent, physical and severe physical punishment, emotional violence, exclusively or only physical punishment, exclusively or only emotional violence, both physical and emotional violence). Disparities by child age, sex, residence, maternal education, household wealth, and world regions were computed. We also assessed changes over time for countries with multiple surveys.

**Results:** Only 19.1% of children experienced exclusively non-violent discipline; 55.0% and 12.7% physical and severe physical punishment; and 64.0% emotional violence. Violent discipline was highest among 6-9y-olds, in Sub-Saharan Africa, and in poorer households. Sex differences were more limited. Use of only non-violent discipline slightly increased in 26 countries, while physical and emotional violence decreased in 33 and 31 countries, respectively. Yet, in some countries, violent discipline increased over time.

**Conclusions:** Despite policy efforts to increase its use, exclusive non-violent discipline remains low, and violent methods are widespread. Targeted and context-specific interventions for specific age groups and poorer households curb violence exposure at home.

## INTRODUCTION

Disciplinary practices - the strategies parents and caregivers use to manage children’s behaviour - are pivotal determinants of child health, development, and well-being globally.^1,2^ Corporal punishment, including slapping or hitting, and emotional abuse, such as yelling or name-calling, are typically perpetrated by parents or caregivers.^3–5^ These methods are associated with adverse long-term outcomes cross-culturally, including chronic pain, mental health problems such as depression or anxiety, and decreased cognitive performance, remaining a major global health challenge.^6–13^ Exposure to violent discipline at home also increases the likelihood of perpetrating it as a parent, highlighting the urgency of effective interventions to break this intergenerational cycle.^14^

Given these lifelong and intergenerational sequelae, safety from corporal and emotional violence at home is a global health policy goal across multiple frameworks, including in Targets 16.2 and 5.2 of the Sustainable Development Goals (SDGs), the World Health Organisation’s INSPIRE framework, and the WHO-UNICEF-Lancet Commission.^15,16^ As of October 2025, about 70 countries have enacted full legal prohibitions on corporal punishment, and more countries have committed to reforming national laws to achieve a ban.^17^

Despite this policy prominence, substantial evidence gaps remain. First, most global estimates focus predominantly on violent discipline, with fewer studies estimating the prevalence of exclusively non-violent practices.^10^ Understanding the extent and distribution of non-harmful discipline is key to designing effective policies and programs.^18,19^ Second, evidence concentrates heavily on discipline during early-childhood, with limited data on middle-childhood and early-adolescence.^10,20–22^ Yet, discipline practices often vary by age, and their consequences on children’s health and development may change based on children’s developmental stage.^10,23^ Additionally, the few studies that have examined discipline among 2-14y-olds also have some limitations, including not using the most updated data; limited examination of exclusively non-violent practices of age-based differences in children’s experiences of different disciplinary practices.^24–29^ Third, although the SDGs emphasise monitoring inequalities by sex, household backgrounds, and place of residence, few studies explore such disparities, hindering targeting efforts.^4,5,18,24^ Fourth, while globalisation, social media, parental education expansion, and urbanisation are all factors driving changes in disciplinary practices^30–33^, most existing studies are cross-sectional, which limits our understanding of how different disciplinary practices may be evolving over time across contexts. These data limitations hinder the monitoring of discipline practices over space and time, as well as the targeting of priority geographical areas or population groups.

This study addresses these gaps by providing the first global estimates of violent *and* non-violent discipline among 1-14y-olds, using data from 176 nationally-representative surveys (2005-2023) across 88 countries. Estimates are disaggregated by child sex, age, socioeconomic status, urbanicity and world regions, offering an overview of inequalities by children’s characteristics and global regions. We also analyse changes in the prevalence of different discipline practices over time.

## METHODS

### Data source, study design, and participants

We sourced all available surveys that included a module on primary caregivers’ reports of child discipline at home. These comprised the UNICEF Multiple Indicator Cluster Surveys (MICS), including MICS3 (2005-2009), MICS4 (2010-2012), MICS5 (2013-2016), and MICS6 (2017-2023), and the USAID’s Demographic and Health Surveys (DHS). Surveys were grouped into four rounds: round 1 (2005-2009), 2 (2010-2012), 3 (2013-2016), and 4 (2017 onward), following MICS/DHS implementation phases.^34^

We focused on surveys from 2005 onwards because the same child discipline module was included in both surveys in 2005 (UNICEF, 2010). MICS and DHS are highly comparable in terms of national representativeness and sampling approach; both employ a two-stage stratified cluster design, with probabilistic selection of enumeration areas in the first stage, and systematic selection of households in the second. In rounds 2 and 3, both MICS and DHS administered the discipline module for one randomly selected child aged 2-14 years in the household. In subsequent rounds-MICS 5 and all DHS surveys-the module instead referred to one randomly selected child aged 1-14 years. In MICS 6, the design changed again: the module was administered for all children aged 1-4 years and for one randomly selected child aged 5-14 years (with no corresponding change in DHS).

We excluded from the analytical sample: (i) surveys employing a subnational (rather than national) sampling framework (e.g., Kenya, Indonesia, Lebanon, Pakistan); and (ii) surveys without the child discipline module (e.g., Bhutan, Burundi, Egypt, Mali, Mexico). The analytical sample included over 1,544,000 children aged 1-14y with information on child discipline, drawn from 176 nationally-representative surveys across 83 low- and middle-income countries (LMICs) and 5 high-income countries (HICs). The median survey year was 2017, with an interquartile range from 2012 to 2019. We used the median survey year to classify countries by their income level based on the World Bank classification. In 2017, 138 countries with a gross national income per capita below US$12,475 were classified as LMICs, and 79 above that threshold as HICs (World Bank, 2025). The sample ranged from 590 in Saint Lucia to 87,808 in Bangladesh. Repeated cross-sectional data was available in 54 countries. Online supplementary appendix A Tables A1-A2 provide details on the sample, including countries and observations per survey-wave.

### Measures

We constructed measures of disciplinary practices using 11 items from the child discipline module, which was based on the Conflict Tactics Scale Parent-Child version (CTSPC).^35^

Primary caregivers, usually children’s mothers, reported whether they or another adult in the household had used any disciplinary methods in the past month: (1) took away privileges; (2) explained wrong behaviour; (3) gave something else to do; (4) shook child; (5) spanked, hit, or slapped on bottom with bare hand; (6) hit with belt, hairbrush, stick, or other hard object; (7) hit/slapped on the hand, arm or leg; (8) hit/slapped on the face, head or ears; (9) beat up, hit over and over as hard as possible; (10) shouted, yelled or screamed; and (11) called dumb, lazy or another name.

We created binary variables (coded as 1= yes, 0 = no) to capture children’s exposure to different forms of discipline. Any reported occurrence of items (4)-(9) was classified as “physical punishment”, with items (8)-(9) further categorised as “severe physical punishment”. Any of the items (10)-(11) indicated exposure to “emotional violence” “Exclusively or Only non-violent discipline” was defined as exposure to any of (1)-(3) and none of (4)-(11). We also constructed variables for: “exclusively or only physical punishment” (any of 4-9, and none of 10–11); “exclusively or only emotional violence” (any of 10-11, and none of 4-9); “both physical and emotional violence” (any of 4-9 and any of 10-11); and “both non-violent and violent methods” (any of 4-11 and any of 1-3). Children with no reported discipline were classified as receiving “neither violent nor non-violent discipline”.

For brevity, and given their higher prevalence compared with the other categories, the main text only reports on the following categories: “only non-violent”, “physical punishment”, and “emotional violence”. The prevalence of “severe physical violence”, “only physical punishment”, “only emotional violence”, and “both physical and emotional violence” is reported in online supplementary appendix B. All surveys included data on child age and sex, maternal education (defined as equal to 1 if the mother had at least secondary education), urban/rural residence, and survey-specific household wealth quintiles. Descriptive statistics for these variables are reported in online supplementary appendix A Table A3.

### Statistical analysis

Descriptive analyses used survey-provided household weights to ensure representativeness and consistency with previous estimates.^10^ We proceeded stepwise. First, we generated estimates for each country. Second, we computed average values of each indicator across age groups and regions. Children were categorised into three age groups: early-childhood (1-5y), mid-childhood (6-9y), and early-adolescence (10-14y). Countries were grouped into five geographical regions based on UN Statistics Division classifications: Asia and Pacific, Eastern Europe, Latin America and the Caribbean, Middle East and North Africa and Sub-Saharan Africa (SSA). To quantify differences or gaps by subgroups, we computed the absolute differences between population groups for each child discipline indicator. Within each region, we compared differences in discipline by child sex (male/female), urban/rural residence, household wealth (poorest/richest quintile), and maternal education (primary/lower vs secondary/higher). We assessed the statistical significance of the average gaps using two-sided t-tests (p≤0.05). Pairwise deletion was applied to handle missing data.

To assess changes over time, we used data from countries with at least two surveys (N=54). We tracked changes in country-level estimates and age-group differences over time, focusing on average differences across indicators between the earliest and latest rounds. Given that children at age 1 were sampled only starting in MICS 5, we restrict the analytical sample for the trends analyses to children aged 2-14 years. Analyses were conducted using Stata V.18.0 and Python V.3.10.

## RESULTS

Globally, only 19.1% of 1-14y-olds were exposed to only non-violent discipline (online supplementary appendix B Table B1). This global average concealed wide cross-country variation (Figure 1, Panel A), ranging from 45.6% in Bosnia and Herzegovina and 45.0% in Albania to 4.3% in Ghana and 4.6% in Egypt. In 28 countries, mostly in SSA, fewer than 10.0% of children were exclusively disciplined non-violently. In contrast, physical punishment was widespread, affecting 55.0% of children globally. The highest prevalence was in Kiribati (85.0%), with similarly high rates in Central African Republic, Tonga, and Samoa (>78.0%), and lowest in Uruguay and Kazakhstan (<25.0%) (Figure 1, Panel B). Emotional violence was even more prevalent than physical punishment, with 64.0% affected worldwide. Rates exceeded 85.0% in Egypt, Palestine, Ghana, and Burundi (Figure 1, Panel C). Comparison of Figure 1 and online supplementary appendix B Figure B1 shows that severe physical violence, though concentrated in certain countries, occurs far less frequently than milder forms of physical punishment. Online supplementary appendix C Figure C1 Panel A further reveals that most households combine violent and non-violent disciplinary methods. Only a small minority (7.58%) do not use any form of discipline (online supplementary appendix C Figure C1 Panel B).

**Figure 1.**
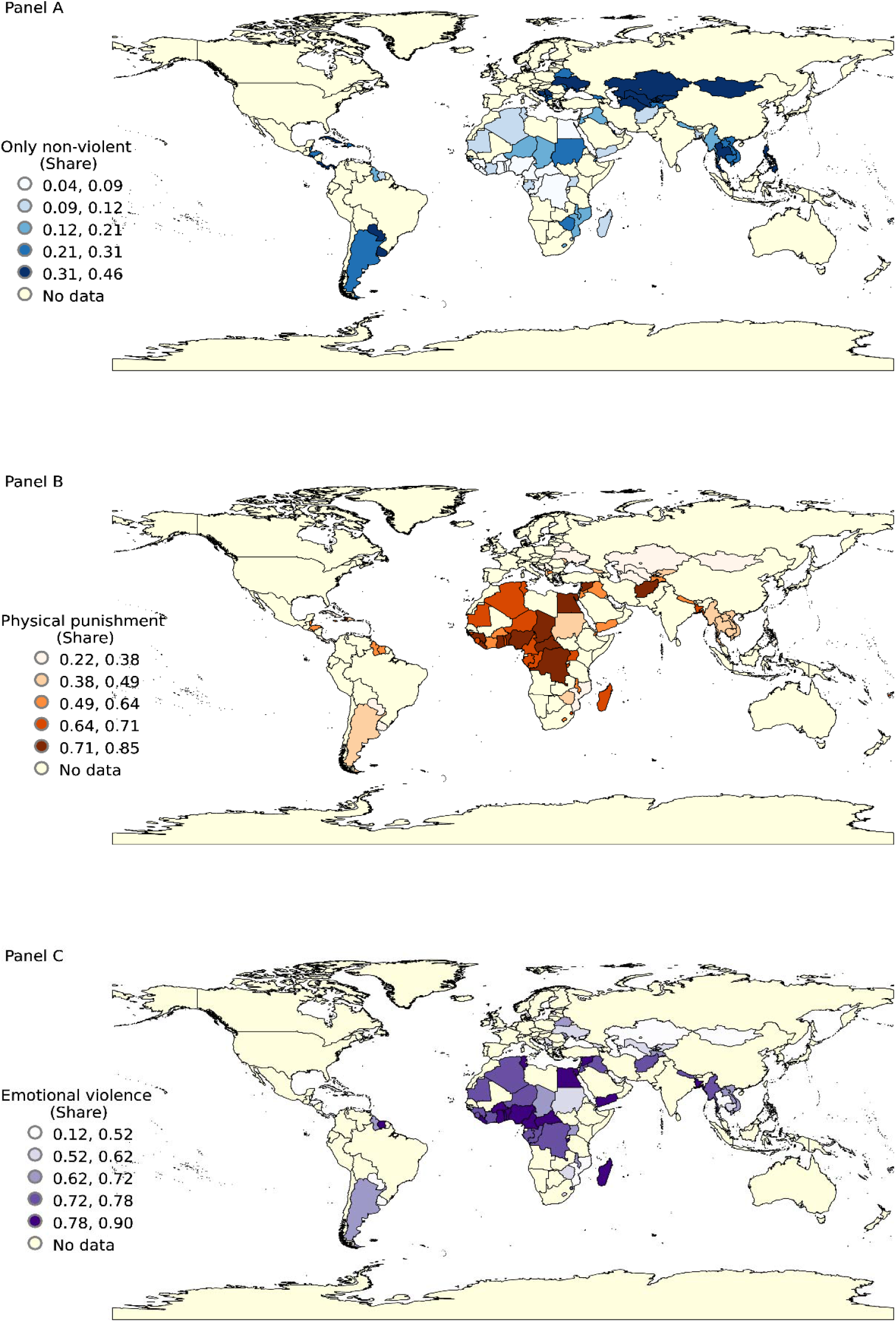
Share of 1-14y-olds exposed to each type of discipline, by country. Online supplementary appendix B Table B1 presents the details of the estimates by country.

### Estimates by age groups and regions

Figure 2 presents age-specific estimates for the three main indicators. Prevalences of *only non-violent* discipline were similar for early childhood (1-5y) and mid-childhood children (6-9y), while they were between 3-5 percentage points higher for early-adolescents (10-14y). Eastern Europe had the highest rates of exclusively non-violent discipline (33.3%), while Sub-Saharan Africa had the lowest (11.7%). Compared to younger children (1-5y), those aged 6-9y were less likely to experience only non-violent discipline in Sub-Saharan Africa (p<0.01), the Middle East and North Africa (p<0.10), and Asia and Pacific (p<0.10). Physical punishment was more prevalent among 6-9y-olds in Sub-Saharan Africa (73.6%) and the Middle-East and North Africa (69.4%), whereas it was highest among 1-5y-olds in the Asia and Pacific region (55.7%), Eastern Europe (39.8%), and Latin America and the Caribbean (43.6%). Globally, emotional violence was highest in mid-childhood (68.0%, online supplementary appendix B Table B2), with the highest prevalences in the Middle-East and North Africa (82.1%) and Sub-Saharan Africa (80.0%). In 68 countries, exposure to only non-violent discipline was frequent among 10-14y-olds, whereas physical and emotional violence was highest among 6-9y-olds (online supplementary appendix B Table B2).

**Figure 2.**
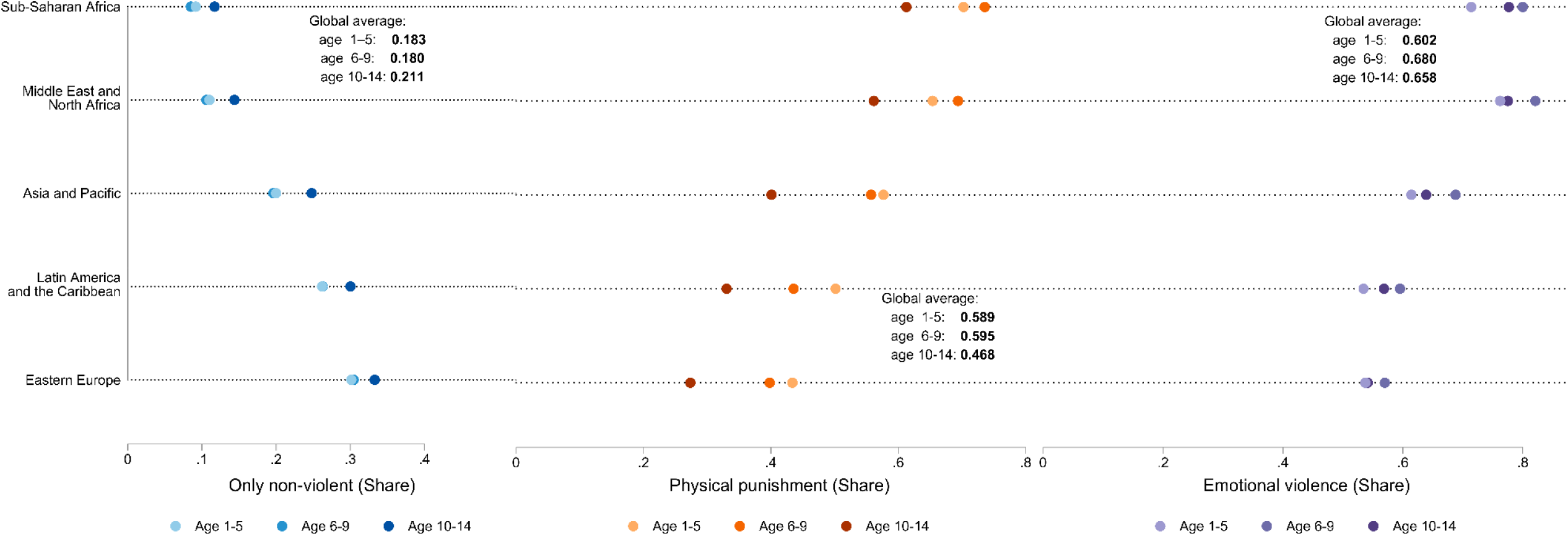
Share of 1-14y-olds exposed to each type of discipline by world region and child age. Online supplementary appendix B Table B2 presents estimates by country and child age. “Global average” gives the unweighted average of the 88 country-estimates.

### Estimates by child sex, household wealth, maternal education, and residence type

Figure 3 shows differences by child sex and region, with p-values of such differences provided in online supplementary appendix B Table B3. Except in Latin America and the Caribbean, boys were less likely than girls to experience only non-violent discipline, with an average boy-girl gap of −2.2 percentage points. Boys also experienced more physical and emotional punishment, with average boy-girl gaps of 4.3 percentage points and 2.9 percentage points, respectively. Across the three indicators, sex differences were smallest in Sub-Saharan Africa and largest in Eastern Europe. Wealthier households were more likely to adopt only non-violent discipline than poorer households in all regions (Figure 4). The wealth gap for only non-violent discipline averaged 6.0 percentage points, being highest in Latin America and the Caribbean (9.3 percentage points, p<0.01) and lowest in Sub-Saharan Africa (1.5 percentage points, p<0.01). There were also large differences between richest vs poorest households in physical punishment (−8.8 percentage points) and emotional violence (−3.4 percentage points), with gaps being largest in Latin America and the Caribbean for physical punishment (−15.5 percentage points, p<0.01) and in the Asia and Pacific region for emotional violence (−7.2 percentage points, p<0.01) (online supplementary appendix B Table B3).

**Figure 3.**
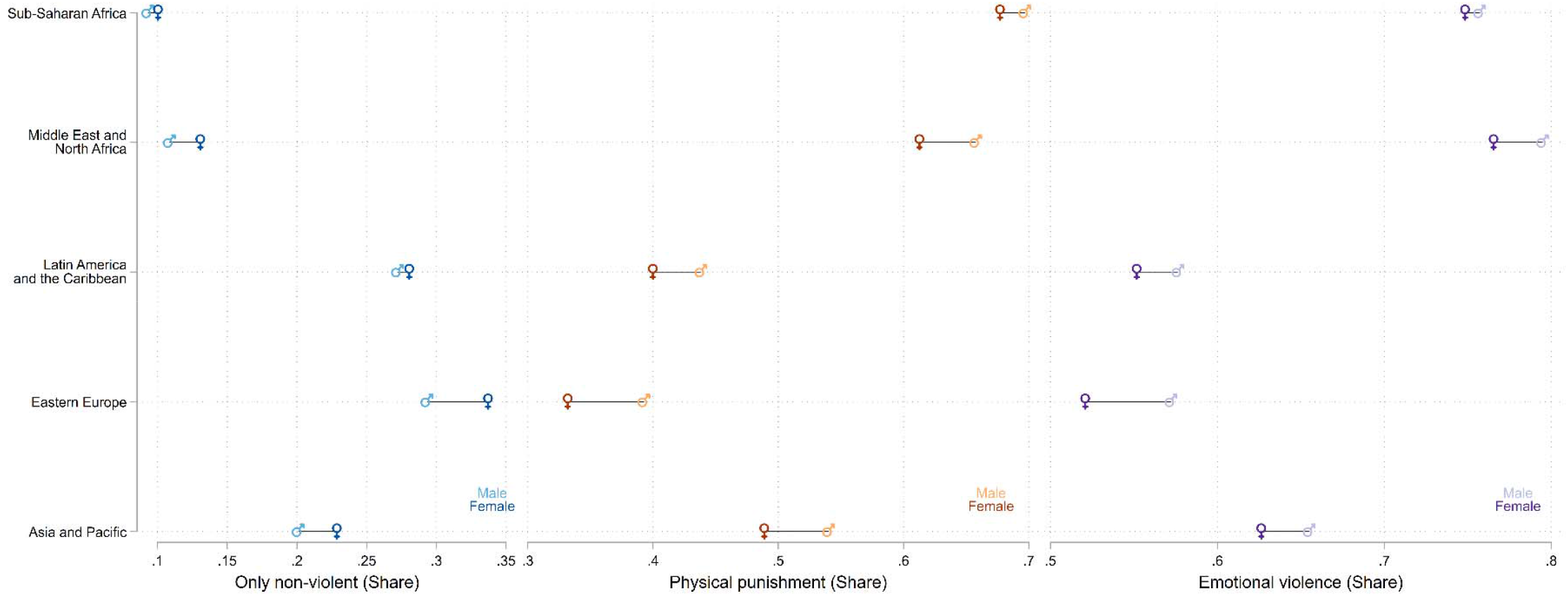
Share of 1-14y-olds exposed to each type of discipline by world region and child gender. Online supplementary appendix B Table B3 presents estimates on gaps and their corresponding p-values.

**Figure 4.**
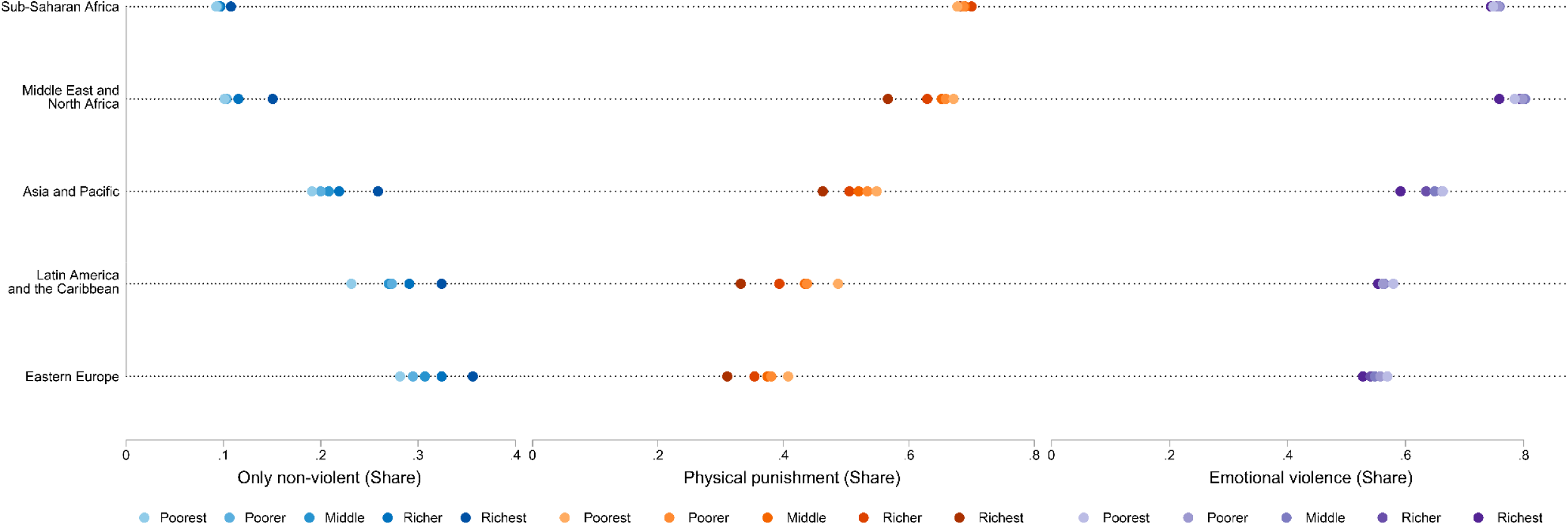
Share of 1-14y-olds exposed to each type of discipline by world region and household wealth. Online supplementary appendix B Table B3 presents estimates on gaps, and their corresponding p-values.

Mothers with secondary or higher education were more likely to use only non-violent methods than those with primary education or less in Eastern Europe (4.6 percentage points) and Asia and Pacific (11.4 percentage points). Less physical and emotional violence was also observed for children of more educated mothers. However, gaps by maternal education were generally smaller compared to wealth, except in the Asia and Pacific (online supplementary appendix B Table B4). Urban-rural differences were observed across nearly all regions and indicators (online supplementary appendix B Table B4). In Asia and Pacific, and the Middle-East and North Africa, urban children were more likely than rural peers to experience only non-violent discipline, with average gaps of 2.2 percentage points. Physical and emotional violence were also lower among urban children in most regions, with the highest gaps in the Asia and Pacific region (physical: −8.0 percentage points, p<0.01; emotional: −9.3 percentage points, p<0.01). Sub-Saharan Africa and Eastern Europe stand out as exceptions to these trends for physical and emotional violence, and emotional violence, respectively. Country-specific differences by sex, residence, maternal education, and wealth are provided in online supplementary appendix B Tables B5-B8.

### Trends

The x-axis of Figure 5 shows the cumulative share of each country’s sample population within a given wave, ordered from highest to lowest values of the discipline measure, while the y-axis reports the estimated prevalence of the outcome in that wave. For example, a value of 0.7 on the x-axis indicates that 70% of the sample lived in countries where the child discipline was at or above the corresponding y-axis level in that wave.

**Figure 5.**
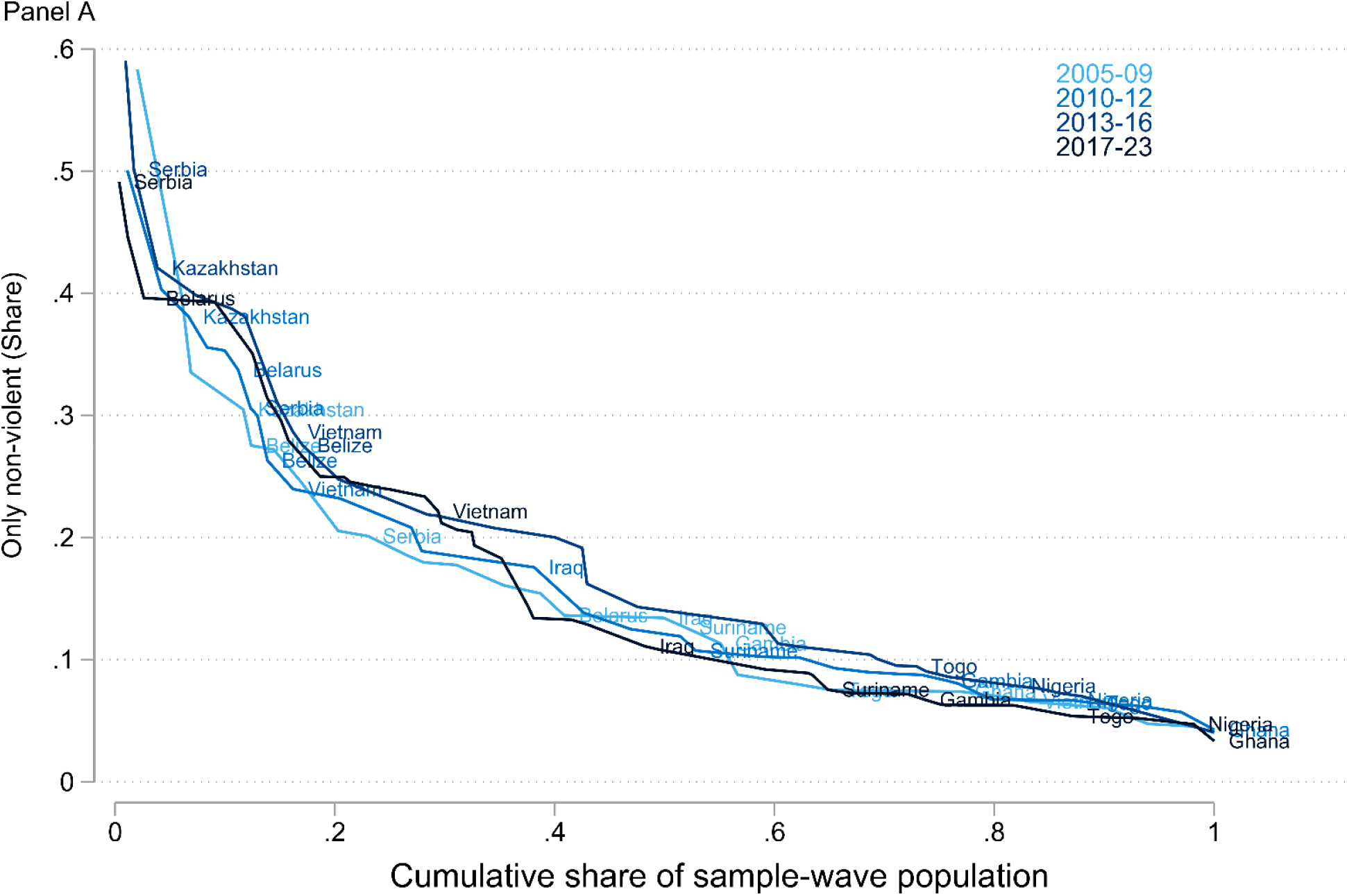
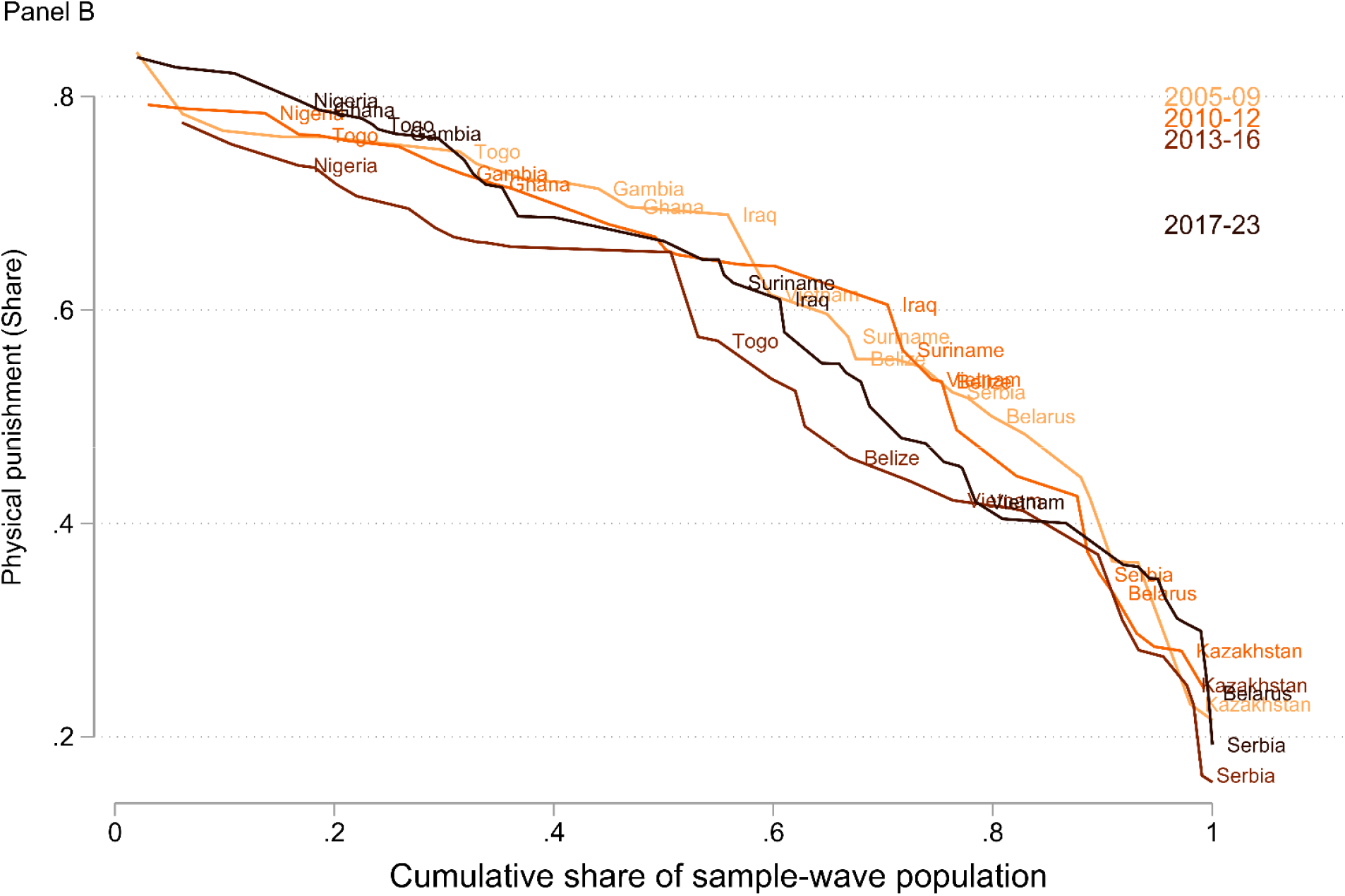
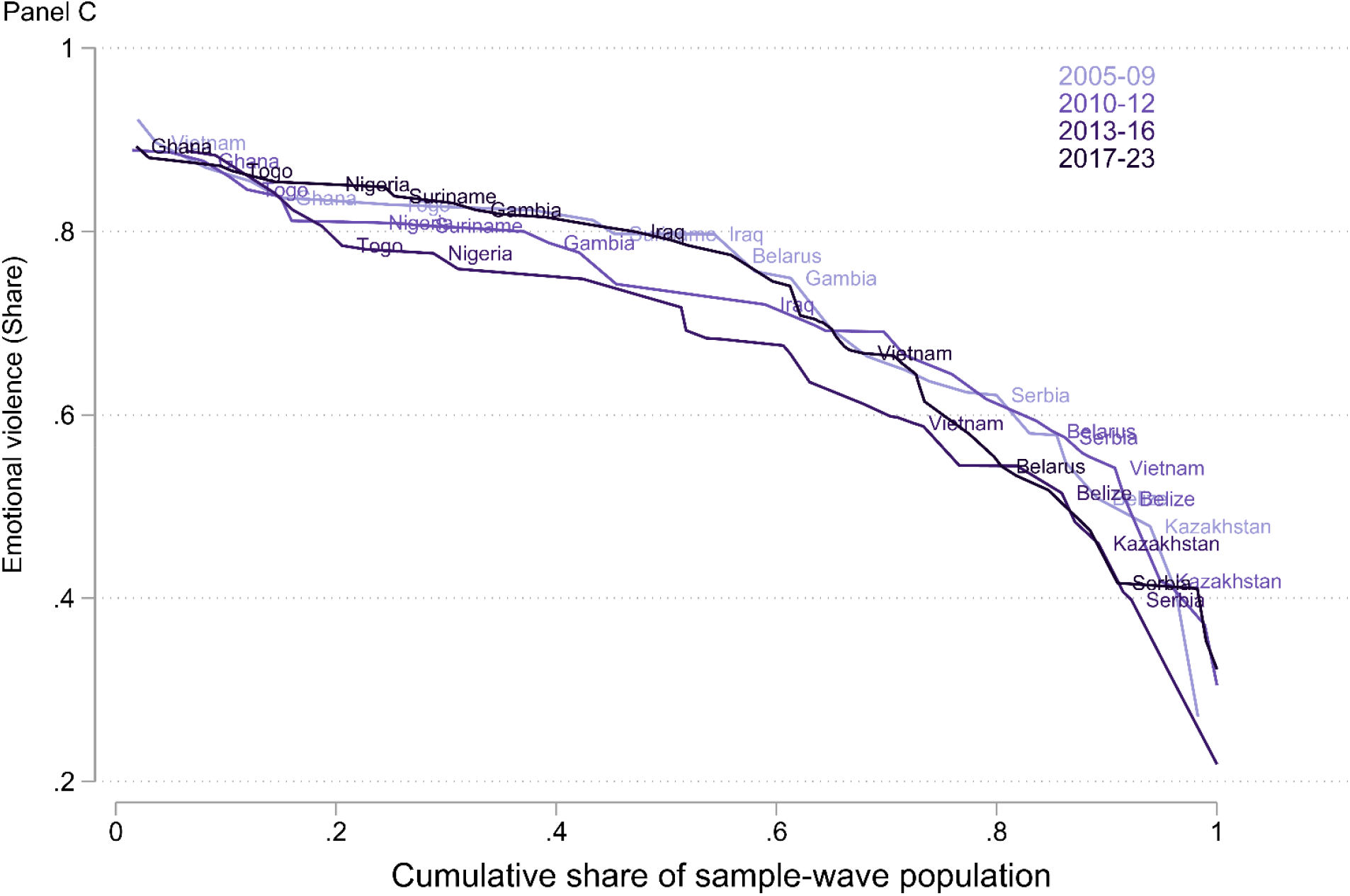
Trends in each type of discipline by country and survey wave. The x-axis represents the cumulative share of each country’s sample population within a given wave, ordered from highest to lowest values of the discipline type. Countries with at least three survey rounds are labelled. See online supplementary appendix B Figure B3 for estimates for the 54 countries.

#### Only non-violent discipline

Globally, progress in only non-violent discipline remained limited (Figure 5, Panel A and online supplementary appendix B Figure B3, Panel A). On average, 70% of the cumulative sample population aged 2-14y lived in countries where fewer than 10.0% experienced only non-violent discipline. Of the 54 countries with at least two surveys, 27 showed increases in only non-violent discipline use, led by Serbia (+29.0 percentage points) and Belarus (+26.0 percentage points), while Algeria (+0.01 percentage points) and Belize (+0.03 percentage points) showed minimal change. Trends in Sub-Saharan Africa were generally less positive, with 12 of 19 countries showing declines in non–violent discipline ranging from 0.4 percentage points to 6.7 percentage points. Bosnia and Herzegovina, and Turkmenistan saw the largest reduction in this indicator, from 58.3% in 2006 to 35.6% in 2011, and 59.0% in 2015 to 29.6% in 2019, respectively. In countries with increases in the use of only non-violent discipline, these were stronger among 2-5y-olds and 10-14y-olds (online supplementary appendix B Figures B4-B5, Panel A). Of 18 countries that experienced declines across all age groups, over half recorded the largest reductions in non-violent discipline use among 6-9y-olds.

#### Physical punishment

Physical punishment remained persistently high between 2005 and 2023. Put simply, about 20% of the cumulative sample lived in countries where more than 70.0% of children experienced physical punishment (Figure 5, Panel B and online supplementary appendix B Figure B3, Panel B). Of the 54 countries, 29 showed declines, with reductions exceeding 25.0 percentage points in Serbia, Yemen, and Belarus. Conversely, there were strong increases in Turkmenistan (+35.3 percentage points), Malawi (+20.7 percentage points), and Bosnia and Herzegovina (+15.8 percentage points). Focusing on age differences, physical punishment declined across all age groups in 25 countries, rose in 15, and showed mixed trends in 14 countries (online supplementary appendix B Figures B4-B5, Panel B). In countries with rising trends, 6-9y-olds were most affected, with increases from +7.2 percentage points in Zimbabwe to +36.9 percentage points in Turkmenistan.

#### Emotional violence

High persistence in emotional violence was also observed, with prevalence exceeding 75.0%, for roughly 20% of the cumulative sampled children (Figure 5, Panel C and online supplementary appendix B Figure B3, Panel C). Of 54 countries, 25 recorded declines, with nine countries showing reductions greater than −10.0 percentage points. Largest reductions occurred in Vietnam (−23.0 percentage points) and Serbia (−23.0 percentage points). Rates increased in 29 countries, ranging from +0.3 percentage points in Iraq to +28.6 percentage points in Kyrgyzstan. Age-specific trends showed declines in 19 countries across all age groups. In 22 countries, there were increasing trends, while mixed patterns were observed in 13 countries (online supplementary appendix B Figures B4-B5, Panel C). Among countries with declines, seven of them, including Belarus, Jordan, Argentina, and Mauritania, saw the greatest reductions among 2-5y-olds, while 10 countries, including Vietnam, Serbia, and Thailand, saw the largest drops among 10-14y-olds. In most countries with increases, emotional violence rose among 2-5y-olds and 6-9y-olds.

## DISCUSSION

This study offers a comprehensive, up-to-date view of home disciplinary practices for children aged 1-14 years and their evolution, highlighting priority populations for promoting non-violent discipline globally. We drew on all available nationally-representative surveys from the past two decades to provide prevalence estimates of: “only non-violent discipline”, “emotional violence”, and “physical and severe physical violence”. Further estimates (including “severe physical violence”, “only physical punishment”, “only emotional violence”, “both physical and emotional violence”, and “no discipline”) were reported as supplementary analyses. Estimates were disaggregated by child age and world regions, revealing disparities across age groups and socioeconomic contexts. Additionally, aligned with SDG 16.2’s focus on inequalities, we assessed within-region and within-country differences by child sex, household wealth, maternal education, and urban/rural residence, providing a detailed view of global patterns and inequities. We also assessed changes over time. To our knowledge, this is among the first studies to provide global estimates and trends in all forms of disciplinary practices, disaggregating them by child and household characteristics. These findings offer much-needed, up-to-date evidence to identify vulnerable groups and target policies promoting non-violent caregiving worldwide.

Only one in five children aged 1-14 years in the study sample were disciplined exclusively with non-violent practices: one in two experienced physical violence, and two in three experienced emotional violence. About one in eight experienced severe physical violence. While these estimates point to strikingly high prevalence of violent discipline at home, they likely underestimate children’s overall exposure to violence, as corporal or emotional punishment occurring in educational and community settings - where violence is often common^36,37^ - is not reflected in our data. Similarly, the MICS scale only elicits experiences of violence at home perpetrated by adults, while sibling violence may also occur.^37^ Thus, we cannot fully characterise the interlocking experiences of violence experienced by children, offering a likely “lower-bound”. Leveraging data from 88 countries, we highlight substantial geographic variation: children in Sub-Saharan Africa (SSA) were the least likely to experience only non-violent discipline, with countries like Ghana, Burundi, Cameroon, Benin, and Nigeria showing fewer than one in ten children disciplined without violence. Physical punishment, including severe forms, exceeded 65.0% in countries across Asia and Pacific, Middle East and North Africa, and SSA, with emotional violence also being prevalent among countries outside Eastern Europe and Central Asia, and Latin America and the Caribbean.

Aligned with the emphasis on uncovering disparities under SDG 16.2e, we identify stark inequities across child characteristics. Across all regions, children aged 10-14 years were more likely to experience only non-violent discipline compared to younger groups, while physical and emotional violence was most prevalent among 6-9y-olds. Interestingly, in countries where violent forms of discipline increased over time, they also increased the most in mid-childhood. Given that most 6-9y-olds attend school, where violence remains widespread^36^, children at this stage may face compounded risks of experiencing violence in multiple settings, potentially increasing the likelihood of lasting developmental harm. These findings thus highlight an important intervention target, as mid-childhood often receives less attention in parenting and community interventions compared with early-childhood or early-adolescence.^38–40^ Data also showed that girls faced less violent discipline than boys, consistent with prior research showing boys’ greater exposure to corporal punishment.^24,41,42^ There were starker inequalities in violent discipline by household wealth than maternal education. Urban caregivers were more likely to use non-violent discipline, though urban children in Sub-Saharan Africa faced higher physical violence, while those in Eastern Europe faced more emotional violence. These contextual patterns underscore entrenched cultural norms sustaining violent discipline beyond socioeconomic or educational factors.^42^ While emotional and physical punishment declined overall, with pronounced reductions in the latter, the use of only non-violent practices remained low over time, indicating slow progress in replacing violent methods. These trends highlight the urgent need to understand country-specific drivers to set up context-appropriate responses. Overall, our detailed disaggregation aligns with SDG 16.2 equity goals and identifies high-risk groups and regions: boys, children aged 6–9 years, those in poorest households, rural areas, and in Sub-Saharan Africa.

Given the differences in the methods and data used, direct comparison with prior estimates is challenging. Most previous studies were based on 2-4y-olds, a more limited set of LMICs, and reported data from a single time point.^5,10,20,29^ Our study, conversely, covers 88 LMICs and HICs with data from multiple rounds for 1-14y-olds. Therefore, we extended existing global estimates with estimates that span over almost two decades.

This study, however, has limitations. First, disciplinary practices may be misreported or underreported. Parents may understate their use of violent methods where corporal punishment is legally prohibited or downplay non-violent methods if harsh discipline is the social norm. Misreporting may also arise when primary caregivers are unaware of disciplinary actions taken by other adults at home, especially in collectivist settings where grandparents or relatives often reside in the households and share caregiving roles. Second, our trend analysis may be affected by cross-cultural differences in survey responses or changes in reporting behaviours, following legal bans on violent discipline or structural changes in other socio-demographic factors that drive discipline behaviours (e.g., urbanisation, maternal education, and so on). Third, while this study represents the most comprehensive analysis of globally available child discipline data to date, its coverage is limited to countries with accessible nationally-representative surveys. Available data have limited coverage of Asia and -Pacific, Eastern Europe, and Latin America and the Caribbean, which may lead to mis-estimation of the true global and regional prevalence of disciplinary practices. Fourth, we could not access some DHS datasets due to ongoing reviews of USAID programmes. Finally, although DHS and MICS are broadly comparable, there may be some instances in which sampling frames and implementation may vary. Similarly, there were changes in the sampling of children across waves, which excluded from the trend analysis children aged 1. Sensitivity checks by including this age group in the latest data round do not change the main conclusions (available upon request). Despite these limitations, this study demonstrates the persistent and ubiquitous nature of harsh discipline, with the aim of motivating further research into its drivers and informing efforts to promote non-violent discipline.

## CONCLUSIONS

Despite declines in violent methods, non-violent discipline remains low and harsh discipline persists at high levels, with large variation in prevalence based on world region of residence and children’s backgrounds. Given the well-established links between home discipline practices and child health and development, these findings underscore the need for culturally-appropriate and context-specific strategies to promote shifts to non-violent discipline globally.

## Acknowledgements

Elisabetta Aurino thanks the European Research Council, Jacobs Foundation and Ministerio de ciencia, innovacion y universidades for the support of her time on this study. John Egyir and Katherina Thomas also thank the afore-mentioned funders for their support of their time as postdoctoral researchers in the lab of Elisabetta Aurino. Elisabetta De Cao thanks the European Research Council and the Italian Ministry of Universities and Research (MUR) for her time on this study. The views and opinions expressed are those of the authors only and do not necessarily reflect those of the European Union or the European Research Council Executive Agency, or any of the other funders. Neither the European Union nor the granting authority can be held responsible for them. The opinion and argument expressed here also do not necessarily reflect the official views of the OECD or of its member states.

## Contributors

JE prepared the data for analysis, analyzed the data, developed the tables and figures, interpreted the results, drafted the original manuscript, and revised the paper. EDC contributed to the interpretation of results, writing, and critically revised the manuscript. KT contributed to the literature search and review, interpreted the results, participated in writing, and revised the manuscript. EA conceived the study idea, contributed to the literature search, interpretation of results, writing of the manuscript and revisions. All authors approved the final manuscript as submitted and agree to be accountable for all aspects of the work.

## Patient and public involvement

Patients and/or the public were not involved in the design, or conduct, or reporting, or dissemination plans of this research.

## Patient consent for publication

Not required.

## Ethics approval

Not required.

## Data availability statement

The project used publicly-accessible secondary data obtained from the IPUMS website (https://globalhealth.ipums.org/); DHS website (https://dhsprogram.com/data/available-datasets.cfm) and the MICS website (https://mics.unicef.org/surveys).

