## Supplementary Material for "Global Levels and Trends in Child Discipline: Evidence from 88 Countries, 2005–2023"

1. Online supplementary appendix A. Sample construction and descriptive statistics
2. Online supplementary appendix B. Additional tables and figures of child discipline
3. Online supplementary **a**ppendix C. Additional child discipline measures

### Online supplementary appendix A. Sample construction and descriptive statistics

Table A1: Samples countries and survey waves included

| | | Round 3 | | Round 4 | | Round 5 | | Round 6 | |
|---|---|---|---|---|---|---|---|---|---|
| World region | country | year | source | year | source | year | source | year | source |
| Asia and Pacific | Afghanistan | | | 2010 | mics | | | 2022 | mics |
| | Bangladesh | | | | | 2012 | mics | 2019 | mics |
| | Cambodia | | | | | | | 2021 | dhs |
| | Fiji | | | | | | | 2021 | mics |
| | Georgia | 2005 | mics | | | | | 2018 | mics |
| | Kazakhstan | 2006 | mics | 2010 | mics | 2015 | mics | | |
| | Kiribati | | | | | | | 2018 | mics |
| | Kyrgyzstan | 2005 | mics | | | 2014 | mics | 2018 | mics |
| | Lao People's Democratic Rep. | 2006 | mics | 2011 | mics | | | 2017 | mics |
| | Mongolia | | | 2010 | mics | 2013 | mics | 2018 | mics |
| | Myanmar | | | | | 2015 | dhs | | |
| | Nepal | | | | | 2014 | mics | 2019 | mics |
| | Philippines | | | | | | | 2022 | dhs |
| | Samoa | | | | | | | 2019 | mics |
| | Tajikistan | 2005 | mics | | | | | 2017 | dhs |
| | Thailand | | | | | 2015 | mics | 2019, 2022 | mics, mics |
| | Tonga | | | | | | | 2019 | mics |
| | Turkmenistan | | | | | 2015 | mics | 2019 | mics |
| | Tuvalu | | | | | | | 2019 | mics |
| | Uzbekistan | | | | | | | 2021 | mics |
| | Vietnam | 2006 | mics | 2010 | mics | 2013 | mics | 2020 | mics |
| Eastern Europe | Albania | 2005 | mics | | | | | | |
| | Belarus | 2005 | mics | 2012 | mics | | | 2019 | mics |

| | | | | | | | | | |
|---|---|---|---|---|---|---|---|---|---|
| | Bosnia and Herzegovina | 2006 | mics | 2011 | mics | | | | |
| | Kosovo | | | | | 2013 | mics | 2019 | mics |
| | Macedonia | 2005 | mics | 2011 | mics | | | 2018 | mics |
| | Moldova | | | 2012 | mics | | | | |
| | Montenegro | 2005 | mics | | | 2013 | mics | 2018 | mics |
| | Serbia | 2005 | mics | 2010 | mics | 2014 | mics | 2019 | mics |
| | Ukraine | 2005 | mics | 2012 | mics | | | | |
| Latin America and the Caribbean | Argentina | | | 2011 | mics | | | 2019 | mics |
| | Barbados | | | 2012 | mics | | | | |
| | Belize | 2006 | mics | 2011 | mics | 2015 | mics | | |
| | Costa Rica | | | 2011 | mics | | | 2018 | mics |
| | Cuba | | | | | 2014 | mics | 2019 | mics |
| | Dominican Rep. | | | | | 2014 | mics | 2019 | mics |
| | El Salvador | | | | | 2014 | mics | | |
| | Guyana | 2006 | mics | | | 2014 | mics | 2019 | mics |
| | Haiti | | | 2012 | dhs | 2016 | dhs | | |
| | Honduras | | | | | | | 2019 | mics |
| | Jamaica | 2005 | mics | 2011 | mics | | | 2022 | mics |
| | Panama | | | | | 2013 | mics | | |
| | Paraguay | | | | | 2016 | mics | | |
| | St. Lucia | | | 2012 | mics | | | | |
| | Suriname | 2006 | mics | 2010 | mics | | | 2018 | mics |
| | Trinidad and Tobago | 2006 | mics | 2011 | mics | | | 2022 | mics |
| | Turks and Caicos Islands | | | | | | | 2019 | mics |
| | Uruguay | | | 2012 | mics | | | | |
| Middle East and North Africa | Algeria | | | 2012 | mics | | | 2018 | mics |
| | Djibouti | 2006 | mics | | | | | | |
| | Egypt | | | | | 2014 | dhs | | |
| | Iraq | 2006 | mics | 2011 | mics | | | 2018 | mics |
| | Jordan | | | 2012 | dhs | | | 2017, 2023 | dhs, dhs |

| | | | | | | | | | |
|---|---|---|---|---|---|---|---|---|---|
| | Qatar | | | 2012 | mics | | | | |
| | Palestine | | | 2010 | mics | 2014 | mics | 2019 | mics |
| | Sudan | | | | | 2014 | mics | | |
| | Syria | 2006 | mics | | | | | | |
| | Tunisia | | | 2011 | mics | | | 2018, 2023 | mics, mics |
| | Yemen | 2006 | mics | | | | | 2022 | mics |
| Sub-Saharan Africa | Benin | | | | | 2014 | mics | 2017, 2021 | dhs, mics |
| | Burkina Faso | 2006 | mics | | | | | | |
| | Burundi | | | | | 2016 | dhs | | |
| | Cameroon | 2006 | mics | | | 2014 | mics | | |
| | Central African Rep. | 2006 | mics | 2010 | mics | | | 2018 | mics |
| | Chad | | | 2010 | mics | 2014 | dhs | 2019 | mics |
| | Comoros | | | | | | | 2022 | mics |
| | Congo | | | 2011 | dhs | 2014 | mics | | |
| | Congo Democratic Rep. | | | 2010 | mics | 2013 | dhs | 2017 | mics |
| | Cote d'Ivoire | 2006 | mics | | | 2016 | mics | 2021 | dhs |
| | Eswatini | | | 2010 | mics | 2014 | mics | 2021 | mics |
| | Gabon | | | | | | | 2019 | dhs |
| | Gambia | 2005 | mics | 2010 | mics | | | 2018 | mics |
| | Ghana | 2006 | mics | 2011 | mics | | | 2017 | mics |
| | Guinea | | | | | 2016 | mics | | |
| | Guinea Bissau | 2006 | mics | | | 2014 | mics | 2018 | mics |
| | Lesotho | | | | | | | 2018 | mics |
| | Liberia | 2007 | dhs | | | | | 2019 | dhs |
| | Madagascar | | | | | | | 2018 | mics |
| | Malawi | | | | | 2013 | mics | 2019 | mics |
| | Mauritania | | | 2011 | mics | 2015 | mics | | |
| | Mozambique | | | | | | | 2022 | dhs |
| | Niger | | | 2012 | dhs | | | | |
| | Nigeria | | | 2011 | mics | 2016 | mics | 2021 | mics |

| | | | | | | | | | |
|---|---|---|---|---|---|---|---|---|---|
| | Sao Tome and Principe | | | | | 2014 | mics | 2019 | mics |
| | Sierra Leone | 2005 | mics | 2010 | mics | | | 2017 | mics |
| | Togo | 2006 | mics | 2010 | mics | 2013 | dhs | 2017 | mics |
| | Uganda | | | | | 2016 | dhs | | |
| | Zimbabwe | | | | | 2014 | mics | 2019 | mics |
| | Countries with 1 wave | | | | | | | 34 | |
| | Countries with 2 waves | | | | | | | 27 | |
| | Countires with >3 waves | | | | | | | 27 | |

*Notes*: The table shows full information on the sampled countries and waves included in the analysis. Albania (DHS 2008, 2017); Armenia (DHS 2010, 2015); Azerbaijan (DHS 2006); and Bolivia (DHS 2003, 2008) are not included because, at the time of writing, the DHS data request was on pause due to the ongoing review of USAID programs. The MICS program does not have a CD module for Armenia, Azerbaijan and Bolivia. Colombia (DHS 1995, 2000, 2005, 2010, 2015) and Peru (DHS 2010, 2011, 2012) are not included in the analysis because, first, the CD module is different from what is provided in the MICS and DHS surveys from other countries. Further, the questionnaire does not include CD questions that ask if each child or a randomly selected child was disciplined. Instead, enumerators ask to women with at least one child living at home how children within the household were disciplined generally. Finally, the CD questions were not framed with time limit (i.e., in the past month), thus capturing regular practices rather than practices within a specific time frame. The MICS program does not have dataset for Colombia and Peru.

Table A2: Sample construction

| Country | (1) Year | (2) $N_{all}$ | (3) $N_{1-5}$ | (4) $N_{6-9}$ | (5) $N_{10-14}$ | (6) $N_{gender}$ | (7) $N_{rur-urb}$ | (8) $N_{momedu}$ | (9) $N_{wealth}$ |
|---|---|---|---|---|---|---|---|---|---|
| Afghanistan | 2010 | 11563 | 4149 | 3360 | 4054 | 11563 | 11563 | 11561 | 11563 |
| Afghanistan | 2022 | 42637 | 28638 | 7368 | 6631 | 42637 | 42637 | 42637 | 42626 |
| Albania | 2005 | 2478 | 589 | 702 | 1187 | 2478 | 2478 | 2478 | 2478 |
| Algeria | 2012 | 16792 | 5937 | 4736 | 6119 | 16792 | 16792 | 16788 | 16792 |
| Algeria | 2018 | 25511 | 13769 | 5825 | 5917 | 25511 | 25511 | 25510 | 25511 |
| Argentina | 2011 | 11785 | 4050 | 3288 | 4447 | 11784 | NA | 11710 | 11785 |
| Argentina | 2019 | 10429 | 5945 | 2270 | 2214 | 10429 | NA | 10413 | 10429 |
| Bangladesh | 2012 | 38723 | 12995 | 11140 | 14588 | 38723 | 38723 | 38721 | 38723 |
| Bangladesh | 2019 | 49085 | 21677 | 12071 | 15337 | 49085 | 49085 | 49085 | 48963 |
| Barbados | 2012 | 891 | 276 | 240 | 375 | 891 | 891 | 882 | 891 |
| Belarus | 2019 | 5317 | 3289 | 1069 | 959 | 5317 | 5317 | 5315 | 5317 |
| Belarus | 2012 | 3334 | 1835 | 698 | 801 | 3334 | 3334 | 3334 | 3334 |
| Belarus | 2005 | 3090 | 1600 | 640 | 850 | 3090 | 3090 | 3090 | 3090 |
| Belize | 2015 | 3024 | 1375 | 740 | 909 | 3024 | 3024 | 3016 | 3024 |
| Belize | 2006 | 1034 | 295 | 346 | 393 | 1034 | 1034 | 1028 | 1034 |
| Belize | 2011 | 2458 | 759 | 721 | 978 | 2458 | 2458 | 2451 | 2458 |
| Benin | 2014 | 9681 | 4154 | 2640 | 2887 | 9681 | 9681 | 9681 | 9681 |
| Benin | 2021 | 20282 | 11720 | 4396 | 4166 | 20282 | 20282 | 20279 | 20282 |
| Benin | 2017 | 10013 | 4426 | 2801 | 2786 | 10013 | 10013 | 7731 | 10013 |
| Bosnia and Herzegovina | 2006 | 2907 | 1553 | 713 | 641 | 2907 | 2907 | 2907 | 2907 |
| Bosnia and Herzegovina | 2011 | 2565 | 1272 | 586 | 707 | 2565 | 2565 | 2565 | 2565 |
| Burkina Faso | 2006 | 4328 | 1468 | 1326 | 1534 | 4328 | 4328 | 3699 | 4328 |
| Burundi | 2016 | 11228 | 4656 | 3084 | 3488 | 11228 | 11228 | 9392 | 11228 |
| Cambodia | 2021 | 14196 | 5023 | 3951 | 5222 | 14196 | 14196 | 1831 | 14196 |
| Cameroon | 2014 | 5985 | 2503 | 1645 | 1837 | 5985 | 5985 | 5982 | 5985 |
| Cameroon | 2006 | 5845 | 1900 | 1827 | 2118 | 5845 | 5845 | 5842 | 5845 |
| Central Africa Rep. | 2018 | 12308 | 7831 | 2441 | 2036 | 12308 | 12308 | 12302 | 12308 |
| Central Africa Rep. | 2010 | 8211 | 3395 | 2467 | 2349 | 8211 | 8211 | 8210 | 8211 |
| Central Africa Rep. | 2006 | 7828 | 3111 | 2452 | 2265 | 7828 | 7828 | 7783 | 7828 |
| Chad | 2010 | 12362 | 5129 | 3683 | 3550 | 12362 | 12362 | 12359 | 12362 |
| Chad | 2014 | 8584 | 3676 | 2516 | 2392 | 8584 | 8584 | 7190 | 8584 |
| Chad | 2019 | 30272 | 19502 | 6010 | 4760 | 30272 | 30272 | 30272 | 30272 |
| Comoros | 2022 | 6927 | 4032 | 1394 | 1501 | 6927 | 6927 | 6916 | 6927 |
| Congo | 2014 | 7867 | 3506 | 2130 | 2231 | 7867 | 7867 | 7865 | 7867 |
| Congo | 2011 | 7514 | 3025 | 2291 | 2198 | 7514 | 7514 | 5785 | 7514 |
| Congo Dem. Rep. | 2017 | 28751 | 18779 | 5330 | 4642 | 28751 | 28751 | 28750 | 28751 |
| Congo Dem. Rep. | 2013 | 6515 | 2866 | 1751 | 1898 | 6515 | 6515 | 5032 | 6515 |
| Congo Dem. Rep. | 2010 | 8796 | 3311 | 2488 | 2997 | 8796 | 8796 | 8789 | 8796 |
| Costa Rica | 2011 | 3042 | 1139 | 821 | 1082 | 3042 | 3042 | 3036 | 3042 |
| Costa Rica | 2018 | 6119 | 3390 | 1383 | 1346 | 6119 | 6119 | 6118 | 6119 |
| Cote d'Ivoire | 2021 | 10236 | 4084 | 2921 | 3231 | 10236 | 10236 | 1581 | 10236 |
| Cote d'Ivoire | 2006 | 6484 | 2262 | 2085 | 2137 | 6484 | 6484 | 6480 | 6484 |
| Cote d'Ivoire | 2016 | 7455 | 3218 | 2093 | 2144 | 7455 | 7455 | 7454 | 7455 |
| Cuba | 2019 | 7970 | 4893 | 1549 | 1528 | 7970 | 7970 | 7969 | 7970 |
| Cuba | 2014 | 5657 | 3855 | 803 | 999 | 5657 | 5657 | 5653 | NA |
| Djibouti | 2006 | 3119 | 877 | 947 | 1295 | 3119 | 3119 | 3117 | NA |
| Dominican Rep. | 2019 | 16818 | 7922 | 4301 | 4595 | 16818 | 16818 | 16777 | 16818 |
| Dominican Rep. | 2014 | 18961 | 10301 | 4003 | 4657 | 18958 | 18961 | 18917 | 18961 |
| Egypt | 2014 | 15664 | 7325 | 3692 | 4647 | 15664 | 15664 | 15387 | 15664 |
| El Salvador | 2014 | 8147 | 4276 | 1717 | 2154 | 8147 | 8147 | 8142 | 8147 |
| Eswatini | 2021 | 3698 | 1983 | 808 | 907 | 3698 | 3698 | 3694 | 3698 |
| Eswatini | 2014 | 3114 | 1126 | 916 | 1072 | 3114 | 3114 | 3094 | 3114 |
| Eswatini | 2010 | 2834 | 900 | 811 | 1123 | 2834 | 2834 | 2829 | 2834 |
| Fiji | 2021 | 3947 | 1939 | 941 | 1067 | 3947 | 3947 | 3942 | 3946 |
| Gabon | 2019 | 3842 | 1622 | 1070 | 1150 | 3842 | 3842 | 1060 | 3842 |
| Gambia | 2010 | 6230 | 2287 | 1903 | 2040 | 6230 | 6230 | 6229 | 6230 |
| Gambia | 2018 | 12877 | 8696 | 2227 | 1954 | 12877 | 12877 | 12840 | 12858 |
| Gambia | 2005 | 4715 | 1473 | 1476 | 1766 | 4715 | 4715 | 4712 | 4715 |
| Georgia | 2018 | 5025 | 2441 | 1330 | 1254 | 5025 | 5025 | 5024 | 5025 |
| Georgia | 2005 | 4311 | 1099 | 1193 | 2019 | 4311 | 4311 | 4311 | 4311 |
| Ghana | 2017 | 14427 | 7928 | 3004 | 3495 | 14427 | 14427 | 14422 | 14426 |
| Ghana | 2011 | 8156 | 2524 | 2458 | 3174 | 8156 | 8156 | 8156 | 8156 |
| Ghana | 2006 | 3936 | 1211 | 1210 | 1515 | 3936 | 3936 | 3936 | 3936 |
| Guinea | 2016 | 6138 | 2601 | 1765 | 1772 | 6138 | 6138 | 6137 | 6138 |
| Guinea Bissau | 2014 | 5158 | 2156 | 1460 | 1542 | 5158 | 5158 | 5134 | 5158 |
| Guinea Bissau | 2006 | 4742 | 1680 | 1429 | 1633 | 4742 | 4742 | 4721 | 4742 |
| Guinea Bissau | 2018 | 10865 | 6618 | 2161 | 2086 | 10865 | 10865 | 10858 | 10865 |

| | | | | | | | | | |
|---|---|---|---|---|---|---|---|---|---|
| Guyana | 2019 | 4786 | 2564 | 1082 | 1140 | 4786 | 4786 | 4707 | 4786 |
| Guyana | 2014 | 3118 | 1559 | 743 | 816 | 3118 | 3118 | 3082 | 3118 |
| Guyana | 2006 | 3122 | 937 | 979 | 1206 | 3115 | 3122 | 2926 | 3122 |
| Haiti | 2016 | 6021 | 2143 | 1766 | 2112 | 6021 | 6021 | 1376 | 6021 |
| Haiti | 2012 | 8503 | 2601 | 2528 | 3374 | 8503 | 8503 | 1051 | 8503 |
| Honduras | 2019 | 16307 | 7926 | 3998 | 4383 | 16307 | 16307 | 16295 | 16307 |
| Iraq | 2018 | 25755 | 14947 | 5453 | 5355 | 25755 | 25755 | 25750 | 25754 |
| Iraq | 2011 | 27908 | 10382 | 7552 | 9974 | 27908 | 27908 | 27905 | 27908 |
| Iraq | 2006 | 13001 | 4392 | 3661 | 4948 | 13001 | 13001 | 12999 | NA |
| Jamaica | 2011 | 2656 | 733 | 782 | 1141 | 2656 | 2656 | 2650 | 2656 |
| Jamaica | 2005 | 2224 | 626 | 670 | 928 | 2224 | 2224 | 2222 | NA |
| Jamaica | 2022 | 3130 | 1331 | 826 | 973 | 3130 | 3130 | 3122 | 3130 |
| Jordan | 2023 | 6056 | 2106 | 1628 | 2322 | 6056 | 6056 | 1671 | 6056 |
| Jordan | 2017 | 5765 | 2100 | 1620 | 2045 | 5765 | 5765 | 1712 | 5765 |
| Jordan | 2012 | 6312 | 2201 | 1652 | 2459 | 6312 | 6312 | 1699 | 6312 |
| Kazakhstan | 2015 | 7472 | 3059 | 2194 | 2219 | 7472 | 7472 | 7470 | 7472 |
| Kazakhstan | 2010 | 6782 | 2444 | 1895 | 2443 | 6782 | 6782 | 6782 | 6782 |
| Kazakhstan | 2006 | 6864 | 1883 | 1837 | 3144 | 6864 | 6864 | 6851 | 6864 |
| Kiribati | 2018 | 3635 | 1939 | 883 | 813 | 3635 | 3635 | 3632 | 3635 |
| Kosovo | 2019 | 2940 | 1422 | 670 | 848 | 2940 | 2940 | 2940 | 2940 |
| Kosovo | 2013 | 2178 | 803 | 510 | 865 | 2178 | 2178 | 2177 | 2178 |
| Kyrgyzstan | 2018 | 6000 | 3186 | 1395 | 1419 | 6000 | 6000 | 5999 | 5999 |
| Kyrgyzstan | 2005 | 3344 | 899 | 968 | 1477 | 3344 | 3344 | 3344 | 3344 |
| Kyrgyzstan | 2014 | 4248 | 2027 | 1094 | 1127 | 4248 | 4248 | 4244 | 4248 |
| Lao People’s Dem. Rep. | 2017 | 21615 | 10925 | 4981 | 5709 | 21615 | 21615 | 21612 | 21615 |
| Lao People’s Dem. Rep. | 2011 | 14485 | 4352 | 4196 | 5937 | 14485 | 14485 | 14483 | 14485 |
| Lao People’s Dem. Rep. | 2006 | 4901 | 1481 | 1432 | 1988 | 4901 | 4901 | 4901 | 4901 |
| Lesotho | 2018 | 6653 | 3132 | 1592 | 1929 | 6653 | 6653 | 6613 | 6616 |
| Liberia | 2007 | 5257 | 1947 | 1680 | 1630 | 5257 | 5257 | 3460 | 5257 |
| Liberia | 2019 | 6125 | 2386 | 1702 | 2037 | 6125 | 6125 | 4020 | 6125 |
| Macedonia | 2018 | 2387 | 1410 | 543 | 434 | 2387 | 2387 | 2387 | 2387 |
| Macedonia | 2011 | 1715 | 716 | 410 | 589 | 1715 | 1715 | 1715 | 1715 |
| Macedonia | 2005 | 3558 | 2239 | 682 | 637 | 3558 | 3558 | 3558 | 3558 |
| Madagascar | 2018 | 19842 | 11497 | 4045 | 4300 | 19842 | 19842 | 19826 | 19833 |
| Malawi | 2013 | 19271 | 7741 | 5499 | 6031 | 19271 | 19271 | 19201 | 19271 |
| Malawi | 2019 | 27004 | 14179 | 6102 | 6723 | 27004 | 27004 | 26990 | 27003 |
| Mauritania | 2015 | 8540 | 3475 | 2375 | 2690 | 8540 | 8540 | 8456 | 8540 |
| Mauritania | 2011 | 7853 | 2703 | 2545 | 2605 | 7853 | 7853 | 7837 | 7853 |
| Moldova | 2012 | 3119 | 1127 | 880 | 1112 | 3119 | 3119 | 3093 | 3119 |
| Mongolia | 2018 | 11147 | 5697 | 2853 | 2597 | 11147 | 11147 | 11137 | 11137 |
| Mongolia | 2013 | 7871 | 3355 | 1925 | 2591 | 7871 | 7871 | 7867 | 7871 |
| Mongolia | 2010 | 5659 | 1883 | 1556 | 2220 | 5659 | 5659 | 5548 | 5659 |
| Montenegro | 2005 | 1179 | 523 | 304 | 352 | 1179 | 1179 | 1179 | 1179 |
| Montenegro | 2018 | 1800 | 1049 | 397 | 354 | 1800 | 1800 | 1800 | 1800 |
| Montenegro | 2013 | 1578 | 788 | 359 | 431 | 1578 | 1578 | 1578 | 1578 |
| Mozambique | 2022 | 5299 | 1965 | 1500 | 1834 | 5299 | 5299 | 695 | 5299 |
| Myanmar | 2015 | 7778 | 2314 | 2316 | 3148 | 7778 | 7778 | 1521 | 7778 |
| Nepal | 2014 | 7523 | 2669 | 1971 | 2883 | 7523 | 7523 | 7461 | 7523 |
| Nepal | 2019 | 11718 | 6324 | 2538 | 2856 | 11718 | 11718 | 11715 | 11718 |
| Niger | 2012 | 8964 | 3464 | 2830 | 2670 | 8964 | 8964 | 7430 | 8964 |
| Nigeria | 2011 | 20349 | 7727 | 6029 | 6593 | 20348 | 20349 | 20323 | 20349 |
| Nigeria | 2021 | 39735 | 23834 | 7977 | 7924 | 39735 | 39735 | 39725 | 39724 |
| Nigeria | 2016 | 21553 | 9175 | 5862 | 6516 | 21553 | 21553 | 21490 | 21553 |
| Panama | 2013 | 6316 | 2585 | 1607 | 2124 | 6316 | 6316 | 6312 | 6316 |
| Paraguay | 2016 | 4652 | 2389 | 1051 | 1212 | 4652 | 4652 | 4640 | 4652 |
| Philippines | 2022 | 17416 | 4806 | 4970 | 7640 | 17416 | 17416 | 14068 | 17416 |
| Qatar | 2012 | 2777 | 819 | 833 | 1125 | 2777 | NA | 2731 | NA |
| St. Lucia | 2012 | 590 | 154 | 169 | 267 | 590 | 590 | 587 | 590 |
| Samoa | 2019 | 3994 | 2386 | 771 | 837 | 3994 | 3994 | 3973 | 3994 |
| Sao Tome and Principe | 2014 | 2177 | 890 | 609 | 678 | 2177 | 2177 | 2170 | 2177 |
| Sao Tome and Principe | 2019 | 3261 | 1680 | 748 | 833 | 3261 | 3261 | 3249 | 3261 |
| Serbia | 2014 | 2747 | 1660 | 597 | 490 | 2747 | 2747 | 2732 | 2747 |
| Serbia | 2005 | 3912 | 1742 | 1015 | 1155 | 3912 | 3912 | 3912 | 3912 |
| Serbia | 2010 | 3087 | 1754 | 742 | 591 | 3087 | 3087 | 3086 | 3087 |
| Serbia | 2019 | 2972 | 1790 | 617 | 565 | 2972 | 2972 | 2972 | 2972 |
| Sierra Leone | 2017 | 18571 | 10607 | 4295 | 3669 | 18571 | 18571 | 18568 | 18571 |
| Sierra Leone | 2005 | 6016 | 2110 | 1910 | 1996 | 6016 | 6016 | 6013 | 6016 |
| Sierra Leone | 2010 | 9185 | 2986 | 3063 | 3136 | 9184 | 9185 | 9184 | 9185 |
| Palestine | 2010 | 9511 | 3186 | 2714 | 3611 | 9511 | 9511 | 9356 | 9511 |
| Palestine | 2014 | 7042 | 2991 | 1722 | 2329 | 7042 | 7042 | 7042 | 7042 |
| Palestine | 2019 | 9251 | 5630 | 1792 | 1829 | 9251 | 9251 | 9250 | 9251 |
| Sudan | 2014 | 11289 | 4881 | 3015 | 3393 | 11288 | 11289 | 11280 | 11289 |
| Suriname | 2006 | 2970 | 1076 | 874 | 1020 | 2970 | 2970 | 2926 | 2970 |
| Suriname | 2018 | 6618 | 3930 | 1398 | 1290 | 6618 | 6618 | 6491 | 6618 |
| Suriname | 2010 | 3771 | 1210 | 1124 | 1437 | 3771 | 3771 | 3729 | 3771 |

| | | | | | | | | | |
|---|---|---|---|---|---|---|---|---|---|
| Syria | 2006 | 12766 | 3857 | 3588 | 5321 | 12766 | 12766 | 12762 | 12766 |
| Tajikistan | 2005 | 5178 | 1472 | 1483 | 2223 | 5178 | 5178 | 5177 | 5178 |
| Tajikistan | 2017 | 5874 | 2422 | 1612 | 1840 | 5874 | 5874 | 5564 | 5874 |
| Thailand | 2015 | 17305 | 8760 | 4113 | 4432 | 17305 | 17305 | 17258 | 17305 |
| Thailand | 2019 | 24900 | 13807 | 5380 | 5713 | 24900 | 24900 | 24888 | 24900 |
| Thailand | 2022 | 19826 | 11050 | 4248 | 4528 | 19826 | 19826 | 19807 | 19826 |
| Togo | 2006 | 4597 | 1342 | 1472 | 1783 | 4597 | 4597 | 4591 | 4597 |
| Togo | 2017 | 8040 | 4446 | 1782 | 1812 | 8040 | 8040 | 8037 | 8040 |
| Togo | 2013 | 6313 | 2494 | 1902 | 1917 | 6313 | 6313 | 4867 | 6313 |
| Togo | 2010 | 4499 | 1600 | 1408 | 1491 | 4499 | 4499 | 4497 | 4499 |
| Tonga | 2019 | 2336 | 1240 | 546 | 550 | 2336 | 2336 | 2329 | 2336 |
| Trinidad and Tobago | 2022 | 3972 | 1735 | 1218 | 1019 | 3972 | 3972 | 3925 | 3972 |
| Trinidad and Tobago | 2006 | 2060 | 581 | 590 | 889 | 2060 | NA | 2050 | 2060 |
| Trinidad and Tobago | 2011 | 1992 | 643 | 569 | 780 | 1992 | 1992 | 1990 | 1992 |
| Tunisia | 2023 | 4470 | 1984 | 1159 | 1327 | 4470 | 4470 | 4470 | 4470 |
| Tunisia | 2011 | 4084 | 1261 | 1148 | 1675 | 4084 | 4084 | 4084 | 4084 |
| Tunisia | 2018 | 6720 | 3378 | 1579 | 1763 | 6720 | 6720 | 6720 | 6720 |
| Turkmenistan | 2015 | 3449 | 1631 | 854 | 964 | 3449 | 3449 | 3448 | 3449 |
| Turkmenistan | 2019 | 6102 | 3502 | 1382 | 1218 | 6102 | 6102 | 6102 | 6102 |
| Turks and Caicos Islands | 2019 | 635 | 319 | 158 | 158 | 635 | 635 | 629 | 635 |
| Tuvalu | 2019 | 767 | 439 | 171 | 157 | 767 | 767 | 760 | 767 |
| Uganda | 2016 | 14380 | 6257 | 3757 | 4366 | 14380 | 14380 | 10373 | 14380 |
| Ukraine | 2005 | 2935 | 1632 | 593 | 710 | 2935 | 2935 | 2935 | 2935 |
| Ukraine | 2012 | 4380 | 2407 | 953 | 1020 | 4380 | 4380 | 4376 | 4380 |
| Uruguay | 2012 | 2037 | 1002 | 495 | 540 | 2037 | 2037 | 2033 | 2037 |
| Uzbekistan | 2021 | 3735 | 1988 | 853 | 894 | 3735 | 3735 | 3731 | 3735 |
| Vietnam | 2013 | 5223 | 1932 | 1528 | 1763 | 5223 | 5223 | 5221 | 5223 |
| Vietnam | 2006 | 2433 | 646 | 660 | 1127 | 2433 | 2433 | 2433 | 2433 |
| Vietnam | 2020 | 9206 | 4266 | 2533 | 2407 | 9206 | 9206 | 9204 | 9200 |
| Vietnam | 2010 | 6390 | 2002 | 1804 | 2584 | 6390 | 6390 | 6388 | 6390 |
| Yemen | 2006 | 2854 | 960 | 841 | 1053 | 2854 | 2854 | 2851 | 2854 |
| Yemen | 2022 | 27038 | 17019 | 4972 | 5047 | 27038 | 27038 | 27027 | 27005 |
| Zimbabwe | 2019 | 10642 | 5597 | 2602 | 2443 | 10642 | 10642 | 10617 | 10631 |
| Zimbabwe | 2014 | 11512 | 4781 | 2995 | 3736 | 11512 | 11512 | 11463 | 11512 |
| Total | | 1544183 | 719843 | 385664 | 438676 | 1544169 | 1517132 | 1458016 | 1517131 |

*Notes*: The table gives the number of observations for each country-wave, detailing how we compile the sample that we use in the main prevalence analysis and those for key sub-group analyses. Column (1) gives the survey year with child discipline. Column (2) reports the total number of observations in the year. Column (3) gives the number of observations for children aged 1-5y. Column (4) gives the number of observations for children aged 6-9y. Column (5) gives the number of observations for children aged 10-14y. Column (6) gives the number of observations with information on gender, while Column (7) gives the number of observations with information on rural residence. Columns (8) and (9) report the number of observations with information on mothers' education and wealth, respectively. "NA" implies information was not available in the survey wave. Data on (i) residence type (rural or urban) available in 172 surveys; (ii) wealth in 171 surveys; and (iii) age, gender and maternal education in all 176 surveys.

Table A3: Descriptive statistics

| Country | Age | Proportion female | Proportion aged 1-5 | Proportion aged 6-9 | Proportion with primary/lower education mothers | Proportion urban | Household size | Proportion in poorest household |
|---|---|---|---|---|---|---|---|---|
| Afghanistan | 5.586 | 0.482 | 0.596 | 0.202 | 0.911 | 0.221 | 9.428 | 0.224 |
| Albania | 8.749 | 0.463 | 0.240 | 0.283 | 0.023 | 0.386 | 4.997 | 0.212 |
| Algeria | 6.587 | 0.482 | 0.477 | 0.246 | 0.383 | 0.616 | 5.852 | 0.219 |
| Argentina | 7.901 | 0.488 | 0.327 | 0.295 | 0.272 | 1.000 | 4.575 | 0.223 |
| Bangladesh | 7.175 | 0.486 | 0.397 | 0.265 | 0.527 | 0.209 | 5.034 | 0.219 |
| Barbados | 8.113 | 0.487 | 0.305 | 0.269 | 0.042 | 0.643 | 4.459 | 0.165 |
| Belarus | 7.604 | 0.490 | 0.364 | 0.271 | 0.001 | 0.719 | 3.856 | 0.156 |
| Belize | 7.808 | 0.495 | 0.329 | 0.289 | 0.570 | 0.436 | 5.237 | 0.183 |
| Benin | 6.197 | 0.490 | 0.505 | 0.249 | 0.825 | 0.425 | 6.213 | 0.203 |
| Bosnia and Herzegovina | 8.557 | 0.481 | 0.259 | 0.289 | 0.344 | 0.336 | 4.620 | 0.165 |
| Burkina Faso | 7.703 | 0.508 | 0.344 | 0.302 | 0.958 | 0.259 | 7.548 | 0.213 |
| Burundi | 6.906 | 0.505 | 0.421 | 0.273 | 0.918 | 0.090 | 5.430 | 0.235 |
| Cambodia | 7.554 | 0.498 | 0.353 | 0.278 | 0.495 | 0.363 | 4.818 | 0.214 |
| Cameroon | 7.310 | 0.498 | 0.373 | 0.295 | 0.687 | 0.473 | 6.023 | 0.209 |
| Central Africa Rep. | 6.232 | 0.506 | 0.505 | 0.261 | 0.839 | 0.341 | 6.349 | 0.225 |
| Chad | 5.862 | 0.503 | 0.550 | 0.239 | 0.906 | 0.183 | 7.001 | 0.226 |
| Comoros | 5.766 | 0.492 | 0.577 | 0.201 | 0.534 | 0.311 | 6.431 | 0.222 |

| | | | | | | | | |
|---|---|---|---|---|---|---|---|---|
| Congo | 7.028 | 0.493 | 0.414 | 0.287 | 0.331 | 0.648 | 5.421 | 0.208 |
| Congo Dem. Rep. | 5.823 | 0.503 | 0.564 | 0.216 | 0.583 | 0.362 | 6.403 | 0.225 |
| Costa Rica | 6.846 | 0.480 | 0.442 | 0.246 | 0.295 | 0.660 | 4.450 | 0.262 |
| Cote d'Ivoire | 7.098 | 0.504 | 0.397 | 0.291 | 0.844 | 0.482 | 6.299 | 0.216 |
| Cuba | 7.560 | 0.497 | 0.359 | 0.272 | 0.037 | 0.678 | 4.096 | 0.217 |
| Djibouti | 8.213 | 0.484 | 0.279 | 0.304 | 0.851 | 0.955 | 6.834 | NA |
| Dominican Rep. | 6.874 | 0.483 | 0.438 | 0.255 | 0.335 | 0.749 | 4.392 | 0.194 |
| Egypt | 6.640 | 0.470 | 0.463 | 0.240 | 0.331 | 0.358 | 5.027 | 0.170 |
| El Salvador | 7.521 | 0.490 | 0.375 | 0.254 | 0.473 | 0.607 | 4.666 | 0.210 |
| Eswatini | 7.118 | 0.501 | 0.410 | 0.266 | 0.424 | 0.231 | 5.880 | 0.215 |
| Fiji | 6.407 | 0.469 | 0.493 | 0.235 | 0.120 | 0.547 | 5.921 | 0.243 |
| Gabon | 6.865 | 0.511 | 0.425 | 0.280 | 0.220 | 0.905 | 5.559 | 0.175 |
| Gambia | 6.291 | 0.519 | 0.507 | 0.244 | 0.775 | 0.567 | 10.944 | 0.219 |
| Georgia | 7.391 | 0.475 | 0.382 | 0.273 | 0.040 | 0.567 | 5.213 | 0.173 |
| Ghana | 6.798 | 0.502 | 0.451 | 0.249 | 0.539 | 0.454 | 5.805 | 0.200 |
| Guinea | 6.803 | 0.514 | 0.424 | 0.292 | 0.873 | 0.338 | 6.498 | 0.225 |
| Guinea Bissau | 6.273 | 0.498 | 0.509 | 0.236 | 0.883 | 0.350 | 8.395 | 0.226 |
| Guyana | 7.246 | 0.497 | 0.398 | 0.264 | 0.206 | 0.263 | 5.104 | 0.223 |
| Haiti | 7.758 | 0.484 | 0.333 | 0.292 | 0.572 | 0.384 | 5.441 | 0.205 |
| Honduras | 6.463 | 0.485 | 0.480 | 0.249 | 0.605 | 0.409 | 5.018 | 0.230 |
| Iraq | 6.843 | 0.481 | 0.451 | 0.251 | 0.650 | 0.692 | 7.379 | 0.211 |
| Jamaica | 7.764 | 0.491 | 0.334 | 0.285 | 0.071 | 0.543 | 4.763 | 0.212 |
| Jordan | 7.770 | 0.469 | 0.341 | 0.274 | 0.074 | 0.873 | 5.960 | 0.207 |
| Kazakhstan | 7.670 | 0.476 | 0.351 | 0.281 | 0.029 | 0.531 | 4.800 | 0.206 |
| Kiribati | 5.885 | 0.490 | 0.539 | 0.242 | 0.219 | 0.499 | 7.367 | 0.233 |
| Kosovo | 6.982 | 0.467 | 0.438 | 0.232 | 0.151 | 0.394 | 6.407 | 0.229 |
| Kyrgyzstan | 6.779 | 0.482 | 0.453 | 0.251 | 0.019 | 0.343 | 5.561 | 0.214 |
| Lao People's Dem. Rep. | 7.222 | 0.496 | 0.405 | 0.260 | 0.707 | 0.261 | 5.732 | 0.221 |
| Lesotho | 6.624 | 0.510 | 0.467 | 0.246 | 0.529 | 0.359 | 5.184 | 0.215 |
| Liberia | 7.281 | 0.495 | 0.378 | 0.299 | 0.729 | 0.467 | 5.654 | 0.216 |
| Macedonia | 7.643 | 0.449 | 0.379 | 0.249 | 0.449 | 0.553 | 5.587 | 0.230 |
| Madagascar | 5.812 | 0.494 | 0.564 | 0.208 | 0.749 | 0.210 | 5.671 | 0.240 |
| Malawi | 6.486 | 0.509 | 0.475 | 0.251 | 0.823 | 0.135 | 5.087 | 0.227 |
| Mauritania | 7.214 | 0.507 | 0.379 | 0.300 | 0.868 | 0.424 | 6.536 | 0.213 |
| Moldova | 7.798 | 0.480 | 0.350 | 0.280 | 0.008 | 0.354 | 3.939 | 0.152 |
| Mongolia | 6.785 | 0.484 | 0.447 | 0.258 | 0.113 | 0.633 | 4.483 | 0.197 |
| Montenegro | 7.191 | 0.478 | 0.418 | 0.246 | 0.177 | 0.645 | 5.023 | 0.188 |
| Mozambique | 7.204 | 0.510 | 0.381 | 0.288 | 0.777 | 0.325 | 5.438 | 0.223 |
| Myanmar | 8.181 | 0.496 | 0.298 | 0.295 | 0.618 | 0.230 | 5.122 | 0.234 |
| Nepal | 7.057 | 0.481 | 0.421 | 0.248 | 0.576 | 0.446 | 5.337 | 0.217 |
| Niger | 7.025 | 0.497 | 0.396 | 0.315 | 0.961 | 0.153 | 6.606 | 0.217 |
| Nigeria | 6.327 | 0.501 | 0.498 | 0.243 | 0.612 | 0.357 | 6.734 | 0.216 |
| Palestine | 6.768 | 0.479 | 0.460 | 0.242 | 0.419 | 0.748 | 6.513 | 0.197 |
| Panama | 7.636 | 0.467 | 0.343 | 0.282 | 0.275 | 0.665 | 4.844 | 0.199 |
| Paraguay | 7.274 | 0.480 | 0.383 | 0.276 | 0.387 | 0.614 | 4.733 | 0.191 |
| Philippines | 8.344 | 0.483 | 0.271 | 0.292 | 0.170 | 0.513 | 5.295 | 0.224 |
| Qatar | 7.928 | 0.460 | 0.319 | 0.310 | 0.088 | 1.000 | 5.758 | NA |
| St. Lucia | 8.604 | 0.480 | 0.258 | 0.285 | 0.409 | 0.203 | 4.501 | 0.215 |
| Samoa | 5.476 | 0.470 | 0.602 | 0.195 | 0.062 | 0.164 | 8.805 | 0.232 |
| Sao Tome and Principe | 6.581 | 0.493 | 0.468 | 0.250 | 0.615 | 0.667 | 5.025 | 0.203 |
| Serbia | 7.532 | 0.481 | 0.365 | 0.281 | 0.159 | 0.580 | 4.789 | 0.160 |
| Sierra Leone | 6.592 | 0.509 | 0.456 | 0.279 | 0.811 | 0.344 | 6.287 | 0.227 |
| Sudan | 6.838 | 0.501 | 0.436 | 0.265 | 0.782 | 0.290 | 6.465 | 0.210 |
| Suriname | 7.150 | 0.488 | 0.404 | 0.271 | 0.345 | 0.672 | 5.408 | 0.250 |
| Syria | 8.159 | 0.481 | 0.302 | 0.281 | 0.588 | 0.548 | 6.558 | 0.188 |
| Tajikistan | 7.539 | 0.483 | 0.361 | 0.278 | 0.047 | 0.277 | 6.847 | 0.204 |
| Thailand | 7.646 | 0.484 | 0.345 | 0.276 | 0.427 | 0.403 | 40.318 | 0.207 |
| Togo | 6.909 | 0.497 | 0.422 | 0.280 | 0.792 | 0.381 | 5.895 | 0.193 |
| Tonga | 6.102 | 0.465 | 0.525 | 0.239 | 0.027 | 0.200 | 6.960 | 0.224 |
| Trinidad and Tobago | 7.954 | 0.499 | 0.327 | 0.279 | 0.188 | 0.568 | 4.790 | 0.216 |
| Tunisia | 7.020 | 0.482 | 0.429 | 0.257 | 0.425 | 0.660 | 48.211 | 0.193 |
| Turkmenistan | 5.912 | 0.480 | 0.545 | 0.233 | 0.001 | 0.410 | 6.420 | 0.213 |
| Turks and Caicos Islands | 6.693 | 0.487 | 0.437 | 0.289 | 0.000 | 0.962 | 3.768 | 0.164 |
| Tuvalu | 5.567 | 0.474 | 0.576 | 0.221 | 0.185 | 0.608 | 8.254 | 0.227 |
| Uganda | 6.795 | 0.503 | 0.437 | 0.260 | 0.745 | 0.223 | 5.654 | 0.205 |
| Ukraine | 8.183 | 0.496 | 0.284 | 0.309 | 0.000 | 0.688 | 3.946 | 0.192 |
| Uruguay | 8.526 | 0.516 | 0.273 | 0.236 | 0.298 | 0.914 | 4.277 | 0.226 |
| Uzbekistan | 5.981 | 0.482 | 0.541 | 0.224 | 0.003 | 0.457 | 6.047 | 0.216 |
| Vietnam | 7.397 | 0.469 | 0.376 | 0.283 | 0.266 | 0.303 | 4.608 | 0.202 |
| Yemen | 5.537 | 0.484 | 0.597 | 0.197 | 0.714 | 0.292 | 7.926 | 0.225 |
| Zimbabwe | 6.563 | 0.495 | 0.470 | 0.253 | 0.424 | 0.284 | 5.074 | 0.216 |

*Notes*: The table shows country-level averages for key sociodemographic variables using household weights. Summary statistics are shown for 88 LMICs and HICs. Descriptive statistics are computed on the pooled analytical waves for each country. For countries and waves without information on a variable, estimates provided are averages across waves where the variable is included.

### Online supplementary appendix B. Additional tables and figures of child discipline

Table B1: Share of 1-14y-olds exposed different child disciplinary practices

| Country | Only non-violent | Physical punishment | Emotional violence | Severe physical violence | Observations |
|---|---|---|---|---|---|
| Afghanistan | 0.092 | 0.763 | 0.748 | 0.508 | 54200 |
| Albania | 0.450 | 0.472 | 0.118 | 0.085 | 2478 |
| Algeria | 0.105 | 0.673 | 0.779 | 0.179 | 42303 |
| Argentina | 0.232 | 0.444 | 0.644 | 0.088 | 22214 |
| Bangladesh | 0.093 | 0.651 | 0.803 | 0.275 | 87808 |
| Barbados | 0.152 | 0.536 | 0.601 | 0.056 | 891 |
| Belarus | 0.299 | 0.350 | 0.620 | 0.008 | 11741 |
| Belize | 0.270 | 0.502 | 0.503 | 0.052 | 6516 |
| Benin | 0.059 | 0.735 | 0.831 | 0.184 | 39976 |
| Bosnia and Herzegovina | 0.456 | 0.304 | 0.350 | 0.037 | 5472 |
| Burkina Faso | 0.065 | 0.607 | 0.833 | 0.203 | 4328 |
| Burundi | 0.051 | 0.651 | 0.852 | 0.066 | 11228 |
| Cambodia | 0.247 | 0.408 | 0.568 | 0.043 | 14196 |
| Cameroon | 0.059 | 0.710 | 0.830 | 0.224 | 11830 |
| Central Africa Rep. | 0.071 | 0.791 | 0.824 | 0.327 | 28347 |
| Chad | 0.121 | 0.715 | 0.701 | 0.278 | 51218 |
| Comoros | 0.241 | 0.434 | 0.522 | 0.064 | 6927 |
| Congo | 0.081 | 0.666 | 0.762 | 0.259 | 15381 |
| Congo Dem. Rep. | 0.063 | 0.774 | 0.769 | 0.350 | 44062 |
| Costa Rica | 0.445 | 0.347 | 0.340 | 0.026 | 9161 |
| Cote d'Ivoire | 0.097 | 0.613 | 0.774 | 0.154 | 24175 |
| Cuba | 0.356 | 0.310 | 0.267 | 0.019 | 13627 |
| Djibouti | 0.176 | 0.641 | 0.541 | 0.210 | 3119 |
| Dominican Rep. | 0.223 | 0.441 | 0.515 | 0.032 | 35779 |
| Egypt | 0.046 | 0.769 | 0.903 | 0.407 | 15664 |
| El Salvador | 0.358 | 0.398 | 0.314 | 0.029 | 8147 |
| Eswatini | 0.110 | 0.650 | 0.748 | 0.078 | 9646 |
| Fiji | 0.187 | 0.651 | 0.616 | 0.117 | 3947 |
| Gabon | 0.114 | 0.658 | 0.720 | 0.208 | 3842 |
| Gambia | 0.083 | 0.731 | 0.781 | 0.165 | 23822 |
| Georgia | 0.238 | 0.392 | 0.615 | 0.111 | 9336 |
| Ghana | 0.043 | 0.746 | 0.867 | 0.139 | 26519 |
| Guinea | 0.066 | 0.741 | 0.778 | 0.110 | 6138 |
| Guinea Bissau | 0.187 | 0.696 | 0.544 | 0.198 | 20765 |
| Guyana | 0.169 | 0.536 | 0.617 | 0.088 | 11026 |
| Haiti | 0.094 | 0.764 | 0.617 | 0.149 | 14524 |
| Honduras | 0.257 | 0.498 | 0.403 | 0.041 | 16307 |
| Iraq | 0.142 | 0.614 | 0.757 | 0.274 | 66664 |
| Jamaica | 0.118 | 0.643 | 0.698 | 0.052 | 8010 |
| Jordan | 0.153 | 0.573 | 0.770 | 0.135 | 18133 |
| Kazakhstan | 0.371 | 0.250 | 0.445 | 0.011 | 21118 |
| Kiribati | 0.080 | 0.850 | 0.778 | 0.210 | 3635 |
| Kosovo | 0.272 | 0.275 | 0.642 | 0.052 | 5118 |
| Kyrgyzstan | 0.322 | 0.420 | 0.538 | 0.036 | 13592 |
| Lao People’s Dem. Rep. | 0.227 | 0.392 | 0.656 | 0.056 | 41001 |
| Lesotho | 0.171 | 0.622 | 0.583 | 0.067 | 6653 |
| Liberia | 0.061 | 0.737 | 0.816 | 0.236 | 11382 |
| Macedonia | 0.253 | 0.505 | 0.600 | 0.097 | 7660 |
| Madagascar | 0.098 | 0.665 | 0.794 | 0.090 | 19842 |
| Malawi | 0.160 | 0.548 | 0.693 | 0.123 | 46275 |
| Mauritania | 0.101 | 0.706 | 0.749 | 0.254 | 16393 |
| Moldova | 0.227 | 0.468 | 0.684 | 0.021 | 3119 |
| Mongolia | 0.396 | 0.275 | 0.385 | 0.042 | 24677 |
| Montenegro | 0.221 | 0.336 | 0.614 | 0.036 | 4557 |
| Mozambique | 0.181 | 0.305 | 0.500 | 0.089 | 5299 |
| Myanmar | 0.153 | 0.428 | 0.739 | 0.118 | 7778 |
| Nepal | 0.142 | 0.579 | 0.760 | 0.164 | 19241 |
| Niger | 0.129 | 0.651 | 0.739 | 0.279 | 8964 |
| Nigeria | 0.063 | 0.758 | 0.801 | 0.286 | 81637 |
| Panama | 0.394 | 0.313 | 0.278 | 0.122 | 6316 |
| Paraguay | 0.409 | 0.370 | 0.300 | 0.039 | 4652 |
| Philippines | 0.370 | 0.367 | 0.470 | 0.031 | 17416 |
| Qatar | 0.424 | 0.313 | 0.389 | 0.057 | 2777 |

| | | | | | |
|---|---|---|---|---|---|
| St. Lucia | 0.193 | 0.430 | 0.591 | 0.052 | 590 |
| Samoa | 0.096 | 0.784 | 0.766 | 0.166 | 3994 |
| Sao Tome and Principe | 0.091 | 0.720 | 0.659 | 0.117 | 5438 |
| Serbia | 0.350 | 0.339 | 0.516 | 0.030 | 12718 |
| Sierra Leone | 0.070 | 0.700 | 0.771 | 0.213 | 33772 |
| Palestine | 0.072 | 0.715 | 0.870 | 0.212 | 25804 |
| Sudan | 0.232 | 0.476 | 0.530 | 0.127 | 11289 |
| Suriname | 0.100 | 0.588 | 0.808 | 0.083 | 13359 |
| Syria | 0.075 | 0.755 | 0.829 | 0.215 | 12766 |
| Tajikistan | 0.227 | 0.496 | 0.654 | 0.132 | 11052 |
| Thailand | 0.336 | 0.445 | 0.469 | 0.026 | 61950 |
| Togo | 0.074 | 0.699 | 0.814 | 0.168 | 23449 |
| Tonga | 0.100 | 0.788 | 0.715 | 0.215 | 2336 |
| Trinidad and Tobago | 0.206 | 0.463 | 0.635 | 0.032 | 8024 |
| Tunisia | 0.099 | 0.681 | 0.820 | 0.206 | 15140 |
| Turkmenistan | 0.421 | 0.366 | 0.455 | 0.010 | 9551 |
| Turks and Caicos Islands | 0.142 | 0.609 | 0.676 | 0.047 | 635 |
| Tuvalu | 0.183 | 0.689 | 0.600 | 0.050 | 767 |
| Uganda | 0.106 | 0.690 | 0.719 | 0.147 | 14380 |
| Ukraine | 0.319 | 0.317 | 0.596 | 0.015 | 7315 |
| Uruguay | 0.379 | 0.222 | 0.471 | 0.029 | 2037 |
| Uzbekistan | 0.322 | 0.294 | 0.573 | 0.043 | 3735 |
| Vietnam | 0.225 | 0.468 | 0.624 | 0.030 | 23252 |
| Yemen | 0.101 | 0.549 | 0.818 | 0.239 | 29892 |
| Zimbabwe | 0.225 | 0.400 | 0.530 | 0.054 | 22154 |
| Mean/total | 0.191 | 0.550 | 0.640 | 0.127 | 1543968 |

*Notes*: The table gives average share of children aged 1-14y exposed to child discipline, by type of violence and country using households weights. The analysis included 88 LMICs and HICs. "Mean" gives the unweighted average of the 88 country-estimates.

Table B2: Prevalence of child disciplinary practices by country and child age

| | **Only non-violent** | | | **Physical punishment** | | | **Emotional violence** | | | **Severe physical violence** | | |
|---|---|---|---|---|---|---|---|---|---|---|---|---|
| **Country** | **Age 1-5** | **Age 6-9** | **Age 10-14** | **Age 1-5** | **Age 6-9** | **Age 10-14** | **Age 1-5** | **Age 6-9** | **Age 10-14** | **Age 1-5** | **Age 6-9** | **Age 10-14** |
| Afghanistan | 0.090 | 0.075 | 0.114 | 0.752 | 0.823 | 0.737 | 0.728 | 0.803 | 0.753 | 0.499 | 0.565 | 0.481 |
| Albania | 0.446 | 0.401 | 0.481 | 0.453 | 0.541 | 0.440 | 0.087 | 0.126 | 0.129 | 0.067 | 0.101 | 0.085 |
| Algeria | 0.096 | 0.098 | 0.126 | 0.698 | 0.721 | 0.585 | 0.762 | 0.812 | 0.778 | 0.170 | 0.204 | 0.174 |
| Argentina | 0.208 | 0.214 | 0.266 | 0.542 | 0.457 | 0.349 | 0.634 | 0.669 | 0.634 | 0.069 | 0.099 | 0.096 |
| Bangladesh | 0.075 | 0.079 | 0.125 | 0.712 | 0.722 | 0.523 | 0.798 | 0.842 | 0.779 | 0.294 | 0.327 | 0.211 |
| Barbados | 0.137 | 0.142 | 0.169 | 0.646 | 0.602 | 0.416 | 0.571 | 0.588 | 0.631 | 0.040 | 0.056 | 0.066 |
| Belarus | 0.304 | 0.299 | 0.295 | 0.456 | 0.374 | 0.227 | 0.571 | 0.652 | 0.645 | 0.005 | 0.013 | 0.008 |
| Belize | 0.236 | 0.269 | 0.300 | 0.546 | 0.556 | 0.425 | 0.454 | 0.535 | 0.520 | 0.039 | 0.059 | 0.059 |
| Benin | 0.071 | 0.042 | 0.052 | 0.727 | 0.801 | 0.686 | 0.770 | 0.891 | 0.894 | 0.130 | 0.232 | 0.245 |
| Bosnia and Herzegovina | 0.471 | 0.447 | 0.453 | 0.354 | 0.356 | 0.242 | 0.273 | 0.363 | 0.387 | 0.026 | 0.053 | 0.033 |
| Burkina Faso | 0.067 | 0.044 | 0.081 | 0.639 | 0.649 | 0.540 | 0.802 | 0.872 | 0.830 | 0.199 | 0.207 | 0.202 |
| Burundi | 0.034 | 0.047 | 0.079 | 0.737 | 0.708 | 0.481 | 0.830 | 0.889 | 0.850 | 0.056 | 0.088 | 0.060 |
| Cambodia | 0.265 | 0.206 | 0.261 | 0.448 | 0.472 | 0.322 | 0.478 | 0.635 | 0.602 | 0.033 | 0.056 | 0.043 |
| Cameroon | 0.047 | 0.053 | 0.077 | 0.757 | 0.762 | 0.610 | 0.797 | 0.858 | 0.842 | 0.203 | 0.259 | 0.218 |
| Central Africa Rep. | 0.072 | 0.060 | 0.078 | 0.793 | 0.837 | 0.736 | 0.794 | 0.861 | 0.847 | 0.285 | 0.385 | 0.352 |
| Chad | 0.125 | 0.100 | 0.135 | 0.702 | 0.775 | 0.681 | 0.662 | 0.761 | 0.737 | 0.237 | 0.337 | 0.317 |
| Comoros | 0.260 | 0.192 | 0.238 | 0.397 | 0.552 | 0.425 | 0.449 | 0.635 | 0.610 | 0.048 | 0.081 | 0.090 |
| Congo | 0.057 | 0.071 | 0.123 | 0.736 | 0.711 | 0.526 | 0.768 | 0.786 | 0.730 | 0.277 | 0.285 | 0.207 |
| Congo Dem. Rep. | 0.049 | 0.058 | 0.104 | 0.780 | 0.840 | 0.696 | 0.750 | 0.816 | 0.771 | 0.318 | 0.435 | 0.351 |
| Costa Rica | 0.419 | 0.447 | 0.480 | 0.431 | 0.375 | 0.207 | 0.287 | 0.386 | 0.378 | 0.019 | 0.039 | 0.025 |
| Cote d'Ivoire | 0.086 | 0.090 | 0.119 | 0.636 | 0.673 | 0.526 | 0.734 | 0.825 | 0.778 | 0.126 | 0.185 | 0.161 |
| Cuba | 0.319 | 0.342 | 0.402 | 0.321 | 0.350 | 0.270 | 0.217 | 0.321 | 0.274 | 0.008 | 0.024 | 0.026 |
| Djibouti | 0.207 | 0.168 | 0.162 | 0.579 | 0.670 | 0.663 | 0.446 | 0.581 | 0.575 | 0.182 | 0.210 | 0.228 |
| Dominican Rep. | 0.187 | 0.243 | 0.256 | 0.490 | 0.468 | 0.348 | 0.489 | 0.543 | 0.528 | 0.023 | 0.041 | 0.038 |
| Egypt | 0.037 | 0.036 | 0.069 | 0.817 | 0.822 | 0.649 | 0.899 | 0.934 | 0.883 | 0.400 | 0.461 | 0.373 |
| El Salvador | 0.301 | 0.380 | 0.400 | 0.507 | 0.409 | 0.282 | 0.260 | 0.320 | 0.364 | 0.018 | 0.044 | 0.030 |
| Eswatini | 0.088 | 0.103 | 0.142 | 0.745 | 0.665 | 0.517 | 0.716 | 0.784 | 0.760 | 0.069 | 0.087 | 0.081 |
| Fiji | 0.193 | 0.148 | 0.210 | 0.650 | 0.720 | 0.591 | 0.575 | 0.658 | 0.655 | 0.109 | 0.105 | 0.142 |
| Gabon | 0.094 | 0.100 | 0.157 | 0.679 | 0.747 | 0.541 | 0.677 | 0.766 | 0.737 | 0.169 | 0.274 | 0.203 |
| Gambia | 0.074 | 0.076 | 0.110 | 0.762 | 0.754 | 0.643 | 0.757 | 0.818 | 0.792 | 0.136 | 0.195 | 0.193 |
| Georgia | 0.257 | 0.193 | 0.253 | 0.426 | 0.454 | 0.304 | 0.592 | 0.686 | 0.586 | 0.098 | 0.139 | 0.102 |
| Ghana | 0.036 | 0.034 | 0.062 | 0.804 | 0.800 | 0.616 | 0.843 | 0.902 | 0.875 | 0.114 | 0.170 | 0.149 |
| Guinea | 0.081 | 0.044 | 0.066 | 0.686 | 0.824 | 0.739 | 0.682 | 0.849 | 0.849 | 0.077 | 0.123 | 0.147 |
| Guinea Bissau | 0.219 | 0.138 | 0.170 | 0.668 | 0.770 | 0.682 | 0.464 | 0.618 | 0.637 | 0.158 | 0.244 | 0.237 |
| Guyana | 0.161 | 0.161 | 0.186 | 0.575 | 0.574 | 0.458 | 0.584 | 0.661 | 0.621 | 0.070 | 0.102 | 0.099 |
| Haiti | 0.071 | 0.080 | 0.127 | 0.810 | 0.813 | 0.687 | 0.565 | 0.659 | 0.632 | 0.108 | 0.171 | 0.169 |

| | | | | | | | | | | | | |
|---|---|---|---|---|---|---|---|---|---|---|---|---|
| Honduras | 0.217 | 0.265 | 0.319 | 0.553 | 0.519 | 0.382 | 0.357 | 0.448 | 0.442 | 0.036 | 0.043 | 0.049 |
| Iraq | 0.120 | 0.129 | 0.185 | 0.642 | 0.666 | 0.530 | 0.753 | 0.796 | 0.729 | 0.290 | 0.313 | 0.218 |
| Jamaica | 0.106 | 0.112 | 0.134 | 0.723 | 0.690 | 0.537 | 0.652 | 0.728 | 0.718 | 0.029 | 0.059 | 0.068 |
| Jordan | 0.151 | 0.129 | 0.171 | 0.617 | 0.639 | 0.488 | 0.743 | 0.821 | 0.758 | 0.133 | 0.158 | 0.121 |
| Kazakhstan | 0.368 | 0.361 | 0.382 | 0.291 | 0.285 | 0.185 | 0.386 | 0.490 | 0.467 | 0.008 | 0.014 | 0.011 |
| Kiribati | 0.112 | 0.029 | 0.058 | 0.842 | 0.928 | 0.783 | 0.682 | 0.874 | 0.909 | 0.157 | 0.268 | 0.277 |
| Kosovo | 0.252 | 0.276 | 0.296 | 0.344 | 0.307 | 0.163 | 0.660 | 0.658 | 0.608 | 0.059 | 0.063 | 0.037 |
| Kyrgyzstan | 0.307 | 0.314 | 0.354 | 0.444 | 0.452 | 0.356 | 0.508 | 0.578 | 0.550 | 0.030 | 0.045 | 0.037 |
| Lao People’s Dem. Rep. | 0.230 | 0.193 | 0.251 | 0.462 | 0.434 | 0.273 | 0.608 | 0.716 | 0.667 | 0.057 | 0.067 | 0.048 |
| Lesotho | 0.103 | 0.189 | 0.267 | 0.750 | 0.618 | 0.417 | 0.561 | 0.614 | 0.590 | 0.056 | 0.078 | 0.075 |
| Liberia | 0.063 | 0.055 | 0.064 | 0.712 | 0.782 | 0.725 | 0.769 | 0.855 | 0.835 | 0.202 | 0.252 | 0.260 |
| Macedonia | 0.237 | 0.226 | 0.287 | 0.564 | 0.535 | 0.425 | 0.589 | 0.634 | 0.589 | 0.091 | 0.121 | 0.087 |
| Madagascar | 0.076 | 0.094 | 0.156 | 0.751 | 0.667 | 0.450 | 0.796 | 0.813 | 0.772 | 0.080 | 0.104 | 0.101 |
| Malawi | 0.132 | 0.160 | 0.209 | 0.596 | 0.585 | 0.430 | 0.662 | 0.733 | 0.711 | 0.127 | 0.139 | 0.099 |
| Mauritania | 0.108 | 0.079 | 0.112 | 0.693 | 0.769 | 0.661 | 0.697 | 0.806 | 0.755 | 0.243 | 0.277 | 0.245 |
| Moldova | 0.208 | 0.221 | 0.249 | 0.582 | 0.474 | 0.356 | 0.675 | 0.713 | 0.671 | 0.022 | 0.018 | 0.023 |
| Mongolia | 0.373 | 0.398 | 0.429 | 0.346 | 0.273 | 0.171 | 0.354 | 0.436 | 0.386 | 0.042 | 0.052 | 0.034 |
| Montenegro | 0.202 | 0.208 | 0.254 | 0.382 | 0.394 | 0.236 | 0.600 | 0.662 | 0.597 | 0.032 | 0.045 | 0.036 |
| Mozambique | 0.174 | 0.176 | 0.194 | 0.282 | 0.345 | 0.297 | 0.448 | 0.546 | 0.521 | 0.068 | 0.108 | 0.095 |
| Myanmar | 0.121 | 0.137 | 0.188 | 0.541 | 0.463 | 0.321 | 0.767 | 0.771 | 0.695 | 0.138 | 0.139 | 0.087 |
| Nepal | 0.155 | 0.104 | 0.154 | 0.604 | 0.669 | 0.481 | 0.701 | 0.829 | 0.784 | 0.167 | 0.195 | 0.137 |
| Niger | 0.114 | 0.119 | 0.160 | 0.661 | 0.701 | 0.583 | 0.726 | 0.771 | 0.723 | 0.246 | 0.318 | 0.281 |
| Nigeria | 0.064 | 0.054 | 0.071 | 0.741 | 0.805 | 0.748 | 0.764 | 0.843 | 0.834 | 0.237 | 0.336 | 0.331 |
| Panama | 0.391 | 0.434 | 0.368 | 0.316 | 0.311 | 0.311 | 0.270 | 0.272 | 0.290 | 0.041 | 0.137 | 0.184 |
| Paraguay | 0.343 | 0.426 | 0.469 | 0.484 | 0.363 | 0.248 | 0.290 | 0.287 | 0.321 | 0.030 | 0.047 | 0.043 |
| Philippines | 0.362 | 0.344 | 0.394 | 0.427 | 0.421 | 0.295 | 0.393 | 0.512 | 0.491 | 0.021 | 0.031 | 0.037 |
| Qatar | 0.395 | 0.466 | 0.412 | 0.321 | 0.288 | 0.326 | 0.336 | 0.394 | 0.430 | 0.072 | 0.040 | 0.059 |
| St. Lucia | 0.164 | 0.201 | 0.204 | 0.565 | 0.474 | 0.326 | 0.568 | 0.540 | 0.635 | 0.033 | 0.055 | 0.061 |
| Samoa | 0.126 | 0.067 | 0.035 | 0.747 | 0.842 | 0.835 | 0.684 | 0.865 | 0.912 | 0.122 | 0.222 | 0.241 |
| Sao Tome and Principe | 0.078 | 0.076 | 0.124 | 0.749 | 0.784 | 0.616 | 0.591 | 0.726 | 0.713 | 0.097 | 0.140 | 0.128 |
| Serbia | 0.344 | 0.348 | 0.358 | 0.427 | 0.369 | 0.223 | 0.495 | 0.527 | 0.530 | 0.024 | 0.035 | 0.033 |
| Sierra Leone | 0.075 | 0.062 | 0.068 | 0.654 | 0.756 | 0.721 | 0.708 | 0.817 | 0.833 | 0.160 | 0.244 | 0.271 |
| Palestine | 0.070 | 0.052 | 0.092 | 0.757 | 0.769 | 0.607 | 0.848 | 0.912 | 0.868 | 0.211 | 0.256 | 0.179 |
| Sudan | 0.205 | 0.232 | 0.269 | 0.501 | 0.518 | 0.401 | 0.495 | 0.570 | 0.545 | 0.120 | 0.150 | 0.117 |
| Suriname | 0.083 | 0.096 | 0.124 | 0.679 | 0.615 | 0.451 | 0.807 | 0.822 | 0.800 | 0.072 | 0.091 | 0.090 |
| Syria | 0.067 | 0.057 | 0.092 | 0.771 | 0.817 | 0.702 | 0.791 | 0.865 | 0.832 | 0.220 | 0.248 | 0.190 |
| Tajikistan | 0.265 | 0.185 | 0.221 | 0.485 | 0.548 | 0.466 | 0.582 | 0.716 | 0.678 | 0.122 | 0.153 | 0.124 |
| Thailand | 0.311 | 0.305 | 0.382 | 0.551 | 0.494 | 0.312 | 0.426 | 0.518 | 0.472 | 0.024 | 0.028 | 0.026 |
| Togo | 0.074 | 0.059 | 0.087 | 0.718 | 0.750 | 0.625 | 0.763 | 0.866 | 0.837 | 0.141 | 0.204 | 0.172 |
| Tonga | 0.096 | 0.071 | 0.139 | 0.791 | 0.853 | 0.713 | 0.667 | 0.798 | 0.738 | 0.192 | 0.240 | 0.241 |
| Trinidad and Tobago | 0.179 | 0.204 | 0.230 | 0.589 | 0.500 | 0.333 | 0.612 | 0.669 | 0.631 | 0.027 | 0.030 | 0.037 |
| Tunisia | 0.096 | 0.081 | 0.119 | 0.729 | 0.743 | 0.565 | 0.788 | 0.867 | 0.824 | 0.181 | 0.245 | 0.208 |
| Turkmenistan | 0.433 | 0.366 | 0.449 | 0.396 | 0.407 | 0.251 | 0.400 | 0.545 | 0.496 | 0.010 | 0.010 | 0.011 |
| Turks and Caicos Islands | 0.147 | 0.146 | 0.129 | 0.624 | 0.668 | 0.522 | 0.653 | 0.656 | 0.735 | 0.013 | 0.078 | 0.067 |
| Tuvalu | 0.187 | 0.163 | 0.194 | 0.700 | 0.744 | 0.594 | 0.551 | 0.700 | 0.628 | 0.045 | 0.057 | 0.056 |
| Uganda | 0.095 | 0.095 | 0.132 | 0.712 | 0.740 | 0.615 | 0.696 | 0.752 | 0.723 | 0.130 | 0.177 | 0.145 |
| Ukraine | 0.367 | 0.282 | 0.315 | 0.365 | 0.350 | 0.260 | 0.514 | 0.648 | 0.614 | 0.011 | 0.016 | 0.017 |
| Uruguay | 0.366 | 0.495 | 0.331 | 0.390 | 0.199 | 0.140 | 0.485 | 0.393 | 0.501 | 0.024 | 0.020 | 0.035 |
| Uzbekistan | 0.327 | 0.287 | 0.345 | 0.309 | 0.345 | 0.211 | 0.546 | 0.638 | 0.572 | 0.045 | 0.054 | 0.028 |
| Vietnam | 0.212 | 0.206 | 0.255 | 0.537 | 0.509 | 0.359 | 0.563 | 0.673 | 0.651 | 0.026 | 0.028 | 0.035 |
| Yemen | 0.122 | 0.058 | 0.082 | 0.511 | 0.650 | 0.560 | 0.766 | 0.910 | 0.881 | 0.185 | 0.315 | 0.284 |
| Zimbabwe | 0.194 | 0.223 | 0.280 | 0.470 | 0.415 | 0.267 | 0.498 | 0.579 | 0.542 | 0.055 | 0.068 | 0.041 |
| Mean | 0.183 | 0.180 | 0.211 | 0.589 | 0.595 | 0.468 | 0.602 | 0.680 | 0.658 | 0.112 | 0.149 | 0.132 |

*Notes*: The table shows the average share of children aged 1-14y exposed to child discipline, by country, type of violence, and age group using household weights. The analysis included 88 LMICs and HICs. “Mean” gives the unweighted average of the 88 country-estimates.

Table B3: Aggregate-level gap by gender (Male [M] – Female [F]) and wealth (Richest [T] – Poorest [B])

| By gender | Only non-violent | | | | Physical punishment | | | | Emotional violence | | | | Severe physical violence | | | |
|---|---|---|---|---|---|---|---|---|---|---|---|---|---|---|---|---|
| Region | M | F | M - F | p-value | M | F | M - F | p-value | M | F | M - F | p-value | M | F | M - F | p-value |
| Asia and Pacific | 0.201 | 0.229 | -0.028 | 0.000 | 0.540 | 0.489 | 0.051 | 0.000 | 0.655 | 0.626 | 0.029 | 0.000 | 0.176 | 0.154 | 0.021 | 0.000 |
| Eastern Europe | 0.293 | 0.337 | -0.044 | 0.000 | 0.393 | 0.332 | 0.061 | 0.000 | 0.572 | 0.521 | 0.051 | 0.000 | 0.047 | 0.030 | 0.017 | 0.000 |
| Latin America and the Caribbean | 0.272 | 0.281 | -0.008 | 0.42 | 0.438 | 0.400 | 0.039 | 0.001 | 0.576 | 0.552 | 0.025 | 0.036 | 0.093 | 0.071 | 0.022 | 0.001 |
| Middle East and North Africa | 0.109 | 0.131 | -0.022 | 0.000 | 0.658 | 0.613 | 0.045 | 0.000 | 0.795 | 0.765 | 0.029 | 0.000 | 0.245 | 0.207 | 0.038 | 0.000 |
| Sub-Saharan Africa | 0.093 | 0.100 | -0.007 | 0.000 | 0.697 | 0.677 | 0.020 | 0.000 | 0.757 | 0.748 | 0.009 | 0.000 | 0.215 | 0.199 | 0.015 | 0.000 |
| Mean | 0.194 | 0.215 | -0.022 | | 0.545 | 0.502 | 0.043 | | 0.671 | 0.643 | 0.029 | | 0.155 | 0.132 | 0.023 | |
| **By wealth** | | | | | | | | | | | | | | | | |
| | T | B | T - B | p-value | T | B | T - B | p-value | T | B | T - B | p-value | T | B | T - B | p-value |
| Asia and Pacific | 0.259 | 0.191 | 0.068 | 0.000 | 0.463 | 0.549 | -0.086 | 0.000 | 0.591 | 0.663 | -0.072 | 0.000 | 0.135 | 0.189 | -0.054 | 0.000 |
| Eastern Europe | 0.356 | 0.281 | 0.075 | 0.000 | 0.311 | 0.408 | -0.097 | 0.000 | 0.528 | 0.569 | -0.042 | 0.000 | 0.024 | 0.066 | -0.043 | 0.000 |
| Latin America and the Caribbean | 0.324 | 0.231 | 0.093 | 0.000 | 0.332 | 0.487 | -0.155 | 0.000 | 0.553 | 0.579 | -0.026 | 0.160 | 0.064 | 0.107 | -0.043 | 0.000 |
| Middle East and North Africa | 0.151 | 0.101 | 0.050 | 0.000 | 0.566 | 0.671 | -0.105 | 0.000 | 0.759 | 0.785 | -0.026 | 0.000 | 0.148 | 0.279 | -0.131 | 0.000 |
| Sub-Saharan Africa | 0.108 | 0.093 | 0.015 | 0.000 | 0.682 | 0.677 | 0.005 | 0.058 | 0.745 | 0.748 | -0.004 | 0.102 | 0.192 | 0.209 | -0.017 | 0.000 |
| Mean | 0.240 | 0.180 | 0.060 | | 0.471 | 0.558 | -0.088 | | 0.635 | 0.669 | -0.034 | | 0.113 | 0.170 | -0.058 | |

*Notes*: Gaps are computed as the difference between male and female averages or richest and poorest quintile averages. A positive value implies an (dis)advantage while a negative value implies (dis)disadvantage. The gender gap analysis included 88 LMICs and HICs. The wealth gap analysis included 86 LMICs and HICs – it excludes Cuba (MICS6), Djibouti, Iraq (MICS3), Jamaica (MICS3) and Qatar due to lack of information on wealth. Household wealth quintiles are included in both MICS and DHS surveys as composite index and are based on the characteristics of the household's dwelling and assets. All averages were estimated using household weights. "Mean" gives the unweighted average of the 5 regional-estimates.

Table B4: Aggregate-level gap by maternal education (Secondary/Higher [S] – Primary/Lower [P]) and residential type (Urban [U] – Rural [R])

| By maternal education | Only non-violent | | | | Physical punishment | | | | Emotional violence | | | | Severe physical violence | | | |
|---|---|---|---|---|---|---|---|---|---|---|---|---|---|---|---|---|
| | S | P | S - P | p-value | S | P | S - P | p-value | S | P | S - P | p-value | S | P | S - P | p-value |
| Asia and Pacific | 0.261 | 0.148 | 0.114 | 0.000 | 0.463 | 0.602 | -0.139 | 0.000 | 0.585 | 0.721 | -0.136 | 0.000 | 0.102 | 0.264 | -0.162 | 0.000 |
| Eastern Europe | 0.321 | 0.275 | 0.046 | 0.000 | 0.352 | 0.432 | -0.080 | 0.000 | 0.544 | 0.568 | -0.024 | 0.009 | 0.032 | 0.082 | -0.050 | 0.000 |
| Latin America and the Caribbean | 0.281 | 0.268 | 0.013 | 0.262 | 0.415 | 0.429 | -0.014 | 0.295 | 0.575 | 0.535 | 0.040 | 0.003 | 0.073 | 0.104 | -0.031 | 0.000 |
| Middle East and North Africa | 0.119 | 0.116 | 0.002 | 0.246 | 0.638 | 0.642 | -0.004 | 0.155 | 0.785 | 0.776 | 0.009 | 0.000 | 0.199 | 0.263 | -0.063 | 0.000 |
| Sub-Saharan Africa | 0.097 | 0.093 | 0.003 | 0.002 | 0.698 | 0.695 | 0.003 | 0.087 | 0.747 | 0.760 | -0.013 | 0.000 | 0.196 | 0.216 | -0.020 | 0.000 |
| Mean | 0.216 | 0.180 | 0.036 | | 0.513 | 0.560 | -0.047 | | 0.647 | 0.672 | -0.025 | | 0.120 | 0.186 | -0.065 | |
| **By residence** | | | | | | | | | | | | | | | | |
| | U | R | U - R | p-value | U | R | U - R | p-value | U | R | U - R | p-value | U | R | U - R | p-value |
| Asia and Pacific | 0.263 | 0.190 | 0.073 | 0.000 | 0.463 | 0.542 | -0.080 | 0.000 | 0.580 | 0.672 | -0.093 | 0.000 | 0.121 | 0.188 | -0.067 | 0.000 |
| Eastern Europe | 0.317 | 0.310 | 0.007 | 0.181 | 0.349 | 0.381 | -0.032 | 0.000 | 0.559 | 0.534 | 0.025 | 0.000 | 0.033 | 0.047 | -0.014 | 0.000 |
| Latin America and the Caribbean | 0.412 | 0.394 | 0.018 | 0.189 | 0.348 | 0.350 | -0.002 | 0.859 | 0.336 | 0.327 | 0.009 | 0.455 | 0.063 | 0.069 | -0.006 | 0.426 |
| Middle East and North Africa | 0.124 | 0.103 | 0.021 | 0.000 | 0.632 | 0.652 | -0.021 | 0.000 | 0.780 | 0.793 | -0.014 | 0.000 | 0.213 | 0.256 | -0.043 | 0.000 |
| Sub-Saharan Africa | 0.092 | 0.099 | -0.007 | 0.000 | 0.705 | 0.678 | 0.027 | 0.000 | 0.759 | 0.749 | 0.010 | 0.000 | 0.209 | 0.206 | 0.004 | 0.018 |
| Mean | 0.242 | 0.219 | 0.022 | | 0.499 | 0.521 | -0.021 | | 0.603 | 0.615 | -0.012 | | 0.128 | 0.153 | -0.025 | |

*Notes*: Gaps are computed as the difference between secondary/higher and primary/lower education or urban and rural averages. A positive value implies an (dis)advantage while a negative value implies (dis)advantage. The maternal education gap analysis included 88 LMICs and HICs. The Urban/rural gap analysis included 86 LMICs and HICs – it excludes Argentina (all survey-waves), Qatar (all survey-waves), and Trinidad and Tobago (MICS3) due to lack of information on urban/rural. All averages were estimated using household weights. "Mean" gives the unweighted average of the 5 regional-estimates.

Table B5: Country-level gap by gender (Male [M] – Female [F])

| | Only non-violent | | | | Physical punishment | | | | Emotional violence | | | | Severe physical violence | | | |
|---|---|---|---|---|---|---|---|---|---|---|---|---|---|---|---|---|
| **Country** | **M** | **F** | **M - F** | **p-value** | **M** | **F** | **M - F** | **p-value** | **M** | **F** | **M - F** | **p-value** | **M** | **F** | **M - F** | **p-value** |
| Afghanistan | 0.089 | 0.095 | -0.006 | 0.063 | 0.771 | 0.754 | 0.017 | 0.000 | 0.752 | 0.745 | 0.007 | 0.154 | 0.522 | 0.493 | 0.029 | 0.000 |
| Albania | 0.417 | 0.489 | -0.072 | 0.001 | 0.507 | 0.431 | 0.076 | 0.000 | 0.117 | 0.119 | -0.001 | 0.934 | 0.095 | 0.075 | 0.020 | 0.090 |
| Algeria | 0.096 | 0.115 | -0.019 | 0.000 | 0.692 | 0.652 | 0.040 | 0.000 | 0.792 | 0.765 | 0.027 | 0.000 | 0.194 | 0.164 | 0.030 | 0.000 |
| Argentina | 0.224 | 0.240 | -0.016 | 0.212 | 0.465 | 0.422 | 0.043 | 0.005 | 0.659 | 0.629 | 0.030 | 0.041 | 0.102 | 0.074 | 0.028 | 0.001 |
| Bangladesh | 0.088 | 0.098 | -0.010 | 0.000 | 0.673 | 0.628 | 0.045 | 0.000 | 0.809 | 0.798 | 0.010 | 0.002 | 0.293 | 0.255 | 0.037 | 0.000 |
| Barbados | 0.132 | 0.173 | -0.041 | 0.099 | 0.580 | 0.490 | 0.090 | 0.011 | 0.609 | 0.593 | 0.016 | 0.651 | 0.061 | 0.050 | 0.010 | 0.544 |
| Belarus | 0.278 | 0.322 | -0.044 | 0.000 | 0.380 | 0.320 | 0.060 | 0.000 | 0.650 | 0.589 | 0.061 | 0.000 | 0.010 | 0.007 | 0.003 | 0.259 |
| Belize | 0.263 | 0.278 | -0.015 | 0.231 | 0.524 | 0.481 | 0.043 | 0.002 | 0.508 | 0.497 | 0.010 | 0.454 | 0.055 | 0.049 | 0.006 | 0.285 |
| Benin | 0.059 | 0.059 | -0.001 | 0.805 | 0.739 | 0.731 | 0.007 | 0.141 | 0.830 | 0.831 | -0.001 | 0.862 | 0.192 | 0.175 | 0.018 | 0.000 |
| Bosnia and Herzegovina | 0.425 | 0.489 | -0.064 | 0.001 | 0.339 | 0.266 | 0.073 | 0.000 | 0.395 | 0.303 | 0.092 | 0.000 | 0.038 | 0.035 | 0.003 | 0.655 |
| Burkina Faso | 0.066 | 0.063 | 0.003 | 0.752 | 0.619 | 0.595 | 0.024 | 0.226 | 0.836 | 0.830 | 0.006 | 0.692 | 0.211 | 0.195 | 0.016 | 0.336 |
| Burundi | 0.044 | 0.058 | -0.014 | 0.003 | 0.677 | 0.625 | 0.053 | 0.000 | 0.864 | 0.840 | 0.024 | 0.001 | 0.073 | 0.059 | 0.014 | 0.006 |
| Cambodia | 0.237 | 0.258 | -0.021 | 0.034 | 0.435 | 0.381 | 0.054 | 0.000 | 0.585 | 0.551 | 0.034 | 0.002 | 0.045 | 0.041 | 0.003 | 0.441 |
| Cameroon | 0.054 | 0.063 | -0.009 | 0.108 | 0.723 | 0.696 | 0.027 | 0.006 | 0.839 | 0.821 | 0.017 | 0.035 | 0.237 | 0.212 | 0.025 | 0.004 |
| Central Africa Rep. | 0.071 | 0.071 | 0.000 | 0.994 | 0.798 | 0.785 | 0.013 | 0.032 | 0.831 | 0.816 | 0.015 | 0.007 | 0.340 | 0.314 | 0.026 | 0.000 |
| Chad | 0.121 | 0.122 | -0.001 | 0.704 | 0.717 | 0.712 | 0.005 | 0.343 | 0.703 | 0.700 | 0.002 | 0.645 | 0.282 | 0.273 | 0.008 | 0.090 |
| Comoros | 0.228 | 0.255 | -0.027 | 0.022 | 0.458 | 0.409 | 0.049 | 0.000 | 0.534 | 0.511 | 0.023 | 0.092 | 0.068 | 0.060 | 0.008 | 0.221 |
| Congo | 0.070 | 0.092 | -0.022 | 0.001 | 0.679 | 0.653 | 0.026 | 0.027 | 0.771 | 0.752 | 0.020 | 0.068 | 0.276 | 0.241 | 0.035 | 0.001 |
| Congo Dem. Rep. | 0.057 | 0.068 | -0.011 | 0.002 | 0.787 | 0.762 | 0.025 | 0.000 | 0.777 | 0.762 | 0.015 | 0.016 | 0.360 | 0.341 | 0.019 | 0.008 |
| Costa Rica | 0.447 | 0.443 | 0.004 | 0.832 | 0.364 | 0.329 | 0.035 | 0.066 | 0.344 | 0.334 | 0.010 | 0.591 | 0.031 | 0.020 | 0.011 | 0.065 |
| Cote d'Ivoire | 0.093 | 0.101 | -0.008 | 0.117 | 0.627 | 0.598 | 0.029 | 0.000 | 0.780 | 0.769 | 0.011 | 0.126 | 0.158 | 0.150 | 0.009 | 0.164 |
| Cuba | 0.355 | 0.356 | -0.001 | 0.958 | 0.319 | 0.301 | 0.018 | 0.266 | 0.285 | 0.248 | 0.036 | 0.019 | 0.024 | 0.015 | 0.009 | 0.040 |
| Djibouti | 0.164 | 0.189 | -0.024 | 0.156 | 0.650 | 0.632 | 0.018 | 0.396 | 0.544 | 0.537 | 0.006 | 0.774 | 0.226 | 0.192 | 0.033 | 0.073 |
| Dominican Rep. | 0.220 | 0.225 | -0.005 | 0.492 | 0.465 | 0.415 | 0.050 | 0.000 | 0.519 | 0.510 | 0.009 | 0.261 | 0.038 | 0.026 | 0.012 | 0.000 |
| Egypt | 0.043 | 0.049 | -0.006 | 0.095 | 0.782 | 0.754 | 0.028 | 0.000 | 0.909 | 0.896 | 0.013 | 0.023 | 0.430 | 0.381 | 0.049 | 0.000 |
| El Salvador | 0.343 | 0.373 | -0.030 | 0.038 | 0.431 | 0.365 | 0.066 | 0.000 | 0.327 | 0.300 | 0.027 | 0.051 | 0.037 | 0.021 | 0.017 | 0.000 |
| Eswatini | 0.100 | 0.119 | -0.018 | 0.008 | 0.672 | 0.628 | 0.044 | 0.000 | 0.762 | 0.735 | 0.027 | 0.005 | 0.082 | 0.074 | 0.008 | 0.165 |
| Fiji | 0.172 | 0.204 | -0.032 | 0.017 | 0.671 | 0.627 | 0.044 | 0.007 | 0.639 | 0.590 | 0.048 | 0.004 | 0.122 | 0.112 | 0.009 | 0.393 |
| Gabon | 0.102 | 0.126 | -0.023 | 0.175 | 0.670 | 0.646 | 0.024 | 0.339 | 0.734 | 0.706 | 0.028 | 0.246 | 0.243 | 0.176 | 0.067 | 0.001 |
| Gambia | 0.081 | 0.085 | -0.003 | 0.478 | 0.743 | 0.719 | 0.024 | 0.001 | 0.782 | 0.779 | 0.004 | 0.577 | 0.171 | 0.158 | 0.013 | 0.024 |
| Georgia | 0.217 | 0.262 | -0.045 | 0.000 | 0.415 | 0.367 | 0.048 | 0.000 | 0.635 | 0.593 | 0.042 | 0.001 | 0.125 | 0.095 | 0.031 | 0.000 |
| Ghana | 0.040 | 0.046 | -0.006 | 0.081 | 0.758 | 0.735 | 0.023 | 0.002 | 0.870 | 0.864 | 0.005 | 0.361 | 0.144 | 0.133 | 0.011 | 0.062 |
| Guinea | 0.068 | 0.064 | 0.003 | 0.643 | 0.750 | 0.733 | 0.016 | 0.178 | 0.774 | 0.782 | -0.009 | 0.452 | 0.109 | 0.112 | -0.003 | 0.712 |
| Guinea Bissau | 0.183 | 0.192 | -0.009 | 0.148 | 0.700 | 0.692 | 0.009 | 0.251 | 0.545 | 0.543 | 0.002 | 0.831 | 0.205 | 0.192 | 0.013 | 0.045 |
| Guyana | 0.154 | 0.185 | -0.031 | 0.002 | 0.578 | 0.493 | 0.084 | 0.000 | 0.638 | 0.595 | 0.042 | 0.001 | 0.101 | 0.075 | 0.027 | 0.000 |
| Haiti | 0.089 | 0.100 | -0.011 | 0.064 | 0.777 | 0.751 | 0.026 | 0.002 | 0.640 | 0.593 | 0.047 | 0.000 | 0.168 | 0.129 | 0.038 | 0.000 |
| Honduras | 0.250 | 0.265 | -0.015 | 0.060 | 0.525 | 0.470 | 0.054 | 0.000 | 0.413 | 0.392 | 0.022 | 0.016 | 0.045 | 0.037 | 0.009 | 0.012 |
| Iraq | 0.127 | 0.157 | -0.030 | 0.000 | 0.641 | 0.586 | 0.054 | 0.000 | 0.776 | 0.736 | 0.039 | 0.000 | 0.296 | 0.251 | 0.046 | 0.000 |
| Jamaica | 0.111 | 0.126 | -0.015 | 0.072 | 0.666 | 0.619 | 0.046 | 0.000 | 0.711 | 0.685 | 0.026 | 0.029 | 0.061 | 0.044 | 0.017 | 0.002 |
| Jordan | 0.137 | 0.171 | -0.034 | 0.000 | 0.595 | 0.548 | 0.047 | 0.000 | 0.788 | 0.750 | 0.038 | 0.000 | 0.147 | 0.122 | 0.025 | 0.002 |
| Kazakhstan | 0.340 | 0.405 | -0.065 | 0.000 | 0.285 | 0.212 | 0.073 | 0.000 | 0.479 | 0.408 | 0.070 | 0.000 | 0.014 | 0.007 | 0.007 | 0.000 |
| Kiribati | 0.081 | 0.078 | 0.003 | 0.740 | 0.847 | 0.854 | -0.007 | 0.589 | 0.771 | 0.786 | -0.015 | 0.300 | 0.210 | 0.210 | -0.000 | 0.992 |
| Kosovo | 0.268 | 0.278 | -0.010 | 0.448 | 0.294 | 0.253 | 0.041 | 0.002 | 0.657 | 0.626 | 0.030 | 0.029 | 0.059 | 0.045 | 0.013 | 0.040 |
| Kyrgyzstan | 0.300 | 0.346 | -0.046 | 0.000 | 0.443 | 0.395 | 0.048 | 0.000 | 0.565 | 0.509 | 0.055 | 0.000 | 0.041 | 0.031 | 0.010 | 0.015 |

| | | | | | | | | | | | | | | | | |
|---|---|---|---|---|---|---|---|---|---|---|---|---|---|---|---|---|
| Lao People's Dem. Rep. | 0.215 | 0.240 | -0.024 | 0.000 | 0.417 | 0.366 | 0.051 | 0.000 | 0.670 | 0.641 | 0.029 | 0.000 | 0.065 | 0.047 | 0.018 | 0.000 |
| Lesotho | 0.162 | 0.180 | -0.018 | 0.114 | 0.638 | 0.607 | 0.031 | 0.031 | 0.598 | 0.567 | 0.031 | 0.037 | 0.069 | 0.064 | 0.005 | 0.510 |
| Liberia | 0.059 | 0.063 | -0.003 | 0.631 | 0.744 | 0.730 | 0.013 | 0.248 | 0.821 | 0.811 | 0.010 | 0.330 | 0.241 | 0.230 | 0.011 | 0.318 |
| Macedonia | 0.230 | 0.281 | -0.051 | 0.016 | 0.526 | 0.479 | 0.047 | 0.040 | 0.619 | 0.576 | 0.043 | 0.063 | 0.124 | 0.063 | 0.061 | 0.000 |
| Madagascar | 0.092 | 0.104 | -0.012 | 0.026 | 0.679 | 0.650 | 0.029 | 0.000 | 0.800 | 0.788 | 0.013 | 0.078 | 0.097 | 0.082 | 0.015 | 0.002 |
| Malawi | 0.155 | 0.165 | -0.011 | 0.027 | 0.562 | 0.535 | 0.027 | 0.000 | 0.698 | 0.689 | 0.010 | 0.110 | 0.131 | 0.115 | 0.015 | 0.001 |
| Mauritania | 0.100 | 0.102 | -0.002 | 0.669 | 0.707 | 0.704 | 0.003 | 0.757 | 0.751 | 0.746 | 0.004 | 0.580 | 0.261 | 0.246 | 0.015 | 0.059 |
| Moldova | 0.209 | 0.246 | -0.038 | 0.022 | 0.501 | 0.433 | 0.068 | 0.001 | 0.703 | 0.665 | 0.038 | 0.037 | 0.029 | 0.013 | 0.016 | 0.005 |
| Mongolia | 0.374 | 0.420 | -0.046 | 0.000 | 0.311 | 0.238 | 0.072 | 0.000 | 0.406 | 0.362 | 0.044 | 0.000 | 0.051 | 0.033 | 0.018 | 0.000 |
| Montenegro | 0.228 | 0.213 | 0.015 | 0.369 | 0.359 | 0.310 | 0.049 | 0.006 | 0.627 | 0.601 | 0.027 | 0.164 | 0.039 | 0.033 | 0.006 | 0.399 |
| Mozambique | 0.169 | 0.192 | -0.023 | 0.076 | 0.326 | 0.285 | 0.041 | 0.011 | 0.516 | 0.485 | 0.031 | 0.075 | 0.100 | 0.078 | 0.022 | 0.020 |
| Myanmar | 0.139 | 0.167 | -0.028 | 0.003 | 0.470 | 0.386 | 0.083 | 0.000 | 0.767 | 0.710 | 0.057 | 0.000 | 0.135 | 0.100 | 0.035 | 0.000 |
| Nepal | 0.135 | 0.151 | -0.016 | 0.010 | 0.597 | 0.560 | 0.037 | 0.000 | 0.768 | 0.752 | 0.017 | 0.031 | 0.175 | 0.153 | 0.022 | 0.001 |
| Niger | 0.128 | 0.130 | -0.002 | 0.767 | 0.655 | 0.647 | 0.008 | 0.516 | 0.743 | 0.735 | 0.008 | 0.472 | 0.277 | 0.280 | -0.003 | 0.778 |
| Nigeria | 0.059 | 0.067 | -0.008 | 0.001 | 0.768 | 0.748 | 0.020 | 0.000 | 0.804 | 0.798 | 0.006 | 0.144 | 0.295 | 0.276 | 0.019 | 0.000 |
| Panama | 0.402 | 0.385 | 0.017 | 0.498 | 0.323 | 0.301 | 0.022 | 0.351 | 0.295 | 0.258 | 0.037 | 0.100 | 0.118 | 0.126 | -0.008 | 0.658 |
| Paraguay | 0.392 | 0.428 | -0.036 | 0.070 | 0.399 | 0.338 | 0.061 | 0.001 | 0.299 | 0.301 | -0.001 | 0.941 | 0.046 | 0.031 | 0.015 | 0.055 |
| Philippines | 0.345 | 0.397 | -0.052 | 0.000 | 0.405 | 0.328 | 0.077 | 0.000 | 0.487 | 0.452 | 0.036 | 0.002 | 0.037 | 0.025 | 0.012 | 0.003 |
| Qatar | 0.411 | 0.438 | -0.027 | 0.280 | 0.340 | 0.281 | 0.059 | 0.006 | 0.414 | 0.359 | 0.055 | 0.018 | 0.058 | 0.056 | 0.001 | 0.893 |
| St. Lucia | 0.168 | 0.219 | -0.050 | 0.157 | 0.475 | 0.381 | 0.094 | 0.032 | 0.606 | 0.574 | 0.032 | 0.475 | 0.062 | 0.041 | 0.021 | 0.261 |
| Samoa | 0.088 | 0.105 | -0.018 | 0.077 | 0.810 | 0.754 | 0.056 | 0.000 | 0.787 | 0.742 | 0.045 | 0.002 | 0.178 | 0.151 | 0.027 | 0.037 |
| Sao Tome and Principe | 0.088 | 0.094 | -0.006 | 0.521 | 0.736 | 0.703 | 0.033 | 0.017 | 0.674 | 0.644 | 0.030 | 0.040 | 0.132 | 0.101 | 0.031 | 0.002 |
| Serbia | 0.339 | 0.362 | -0.023 | 0.057 | 0.354 | 0.322 | 0.032 | 0.006 | 0.533 | 0.499 | 0.034 | 0.007 | 0.036 | 0.023 | 0.013 | 0.001 |
| Sierra Leone | 0.066 | 0.073 | -0.006 | 0.050 | 0.709 | 0.692 | 0.017 | 0.002 | 0.774 | 0.769 | 0.005 | 0.377 | 0.219 | 0.207 | 0.012 | 0.016 |
| Palestine | 0.060 | 0.085 | -0.025 | 0.000 | 0.745 | 0.682 | 0.063 | 0.000 | 0.887 | 0.851 | 0.036 | 0.000 | 0.235 | 0.187 | 0.049 | 0.000 |
| Sudan | 0.218 | 0.245 | -0.026 | 0.010 | 0.483 | 0.468 | 0.016 | 0.199 | 0.539 | 0.521 | 0.018 | 0.149 | 0.130 | 0.124 | 0.006 | 0.469 |
| Suriname | 0.090 | 0.110 | -0.019 | 0.003 | 0.609 | 0.566 | 0.043 | 0.000 | 0.823 | 0.793 | 0.030 | 0.000 | 0.090 | 0.075 | 0.015 | 0.007 |
| Syria | 0.070 | 0.080 | -0.010 | 0.029 | 0.778 | 0.730 | 0.048 | 0.000 | 0.843 | 0.814 | 0.028 | 0.000 | 0.232 | 0.197 | 0.035 | 0.000 |
| Tajikistan | 0.208 | 0.248 | -0.040 | 0.000 | 0.528 | 0.461 | 0.067 | 0.000 | 0.676 | 0.630 | 0.046 | 0.000 | 0.147 | 0.115 | 0.032 | 0.000 |
| Thailand | 0.315 | 0.359 | -0.045 | 0.000 | 0.473 | 0.415 | 0.057 | 0.000 | 0.490 | 0.446 | 0.045 | 0.000 | 0.028 | 0.023 | 0.005 | 0.059 |
| Togo | 0.072 | 0.076 | -0.004 | 0.362 | 0.709 | 0.689 | 0.020 | 0.004 | 0.816 | 0.812 | 0.004 | 0.476 | 0.180 | 0.155 | 0.025 | 0.000 |
| Tonga | 0.084 | 0.118 | -0.035 | 0.035 | 0.814 | 0.757 | 0.057 | 0.009 | 0.737 | 0.690 | 0.048 | 0.050 | 0.236 | 0.191 | 0.045 | 0.035 |
| Trinidad and Tobago | 0.196 | 0.216 | -0.020 | 0.086 | 0.493 | 0.433 | 0.060 | 0.000 | 0.660 | 0.611 | 0.049 | 0.000 | 0.037 | 0.026 | 0.011 | 0.018 |
| Tunisia | 0.087 | 0.113 | -0.026 | 0.000 | 0.702 | 0.659 | 0.044 | 0.000 | 0.833 | 0.805 | 0.027 | 0.000 | 0.228 | 0.183 | 0.046 | 0.000 |
| Turkmenistan | 0.408 | 0.435 | -0.028 | 0.012 | 0.386 | 0.344 | 0.042 | 0.000 | 0.467 | 0.443 | 0.024 | 0.031 | 0.011 | 0.008 | 0.003 | 0.208 |
| Turks and Caicos Islands | 0.131 | 0.153 | -0.022 | 0.610 | 0.621 | 0.596 | 0.024 | 0.719 | 0.751 | 0.598 | 0.154 | 0.020 | 0.062 | 0.031 | 0.031 | 0.223 |
| Tuvalu | 0.168 | 0.200 | -0.032 | 0.262 | 0.713 | 0.662 | 0.051 | 0.134 | 0.615 | 0.583 | 0.032 | 0.364 | 0.070 | 0.028 | 0.042 | 0.007 |
| Uganda | 0.099 | 0.113 | -0.014 | 0.017 | 0.692 | 0.687 | 0.005 | 0.566 | 0.725 | 0.713 | 0.012 | 0.155 | 0.146 | 0.147 | -0.001 | 0.880 |
| Ukraine | 0.261 | 0.378 | -0.117 | 0.000 | 0.373 | 0.261 | 0.112 | 0.000 | 0.660 | 0.531 | 0.129 | 0.000 | 0.019 | 0.011 | 0.008 | 0.059 |
| Uruguay | 0.382 | 0.376 | 0.006 | 0.939 | 0.288 | 0.160 | 0.128 | 0.004 | 0.490 | 0.453 | 0.037 | 0.633 | 0.034 | 0.024 | 0.010 | 0.504 |
| Uzbekistan | 0.311 | 0.335 | -0.024 | 0.165 | 0.317 | 0.269 | 0.048 | 0.005 | 0.588 | 0.556 | 0.031 | 0.093 | 0.048 | 0.038 | 0.010 | 0.195 |
| Vietnam | 0.200 | 0.254 | -0.055 | 0.000 | 0.511 | 0.421 | 0.090 | 0.000 | 0.645 | 0.601 | 0.044 | 0.000 | 0.037 | 0.022 | 0.015 | 0.000 |
| Yemen | 0.099 | 0.104 | -0.005 | 0.262 | 0.564 | 0.533 | 0.031 | 0.000 | 0.821 | 0.815 | 0.007 | 0.233 | 0.258 | 0.220 | 0.038 | 0.000 |
| Zimbabwe | 0.223 | 0.227 | -0.004 | 0.480 | 0.412 | 0.387 | 0.025 | 0.000 | 0.533 | 0.528 | 0.005 | 0.497 | 0.058 | 0.051 | 0.007 | 0.023 |
| Mean | 0.181 | 0.202 | -0.021 | | 0.571 | 0.528 | 0.043 | | 0.653 | 0.625 | 0.028 | | 0.136 | 0.118 | 0.019 | |

*Notes*: Gaps are computed as the difference between male and female averages. A positive value implies male (dis)advantage while a negative value implies female (dis)advantage. The analysis included 88 LMICs and HICs. All averages were estimated using household weights. "Mean" gives the unweighted average of the 88 country-estimates.

Table B6: Country-level gap by residential area (Urban [U] – Rural [R])

| | Only non-violent | | | | Physical punishment | | | | Emotional violence | | | | Severe physical violence | | | |
|---|---|---|---|---|---|---|---|---|---|---|---|---|---|---|---|---|
| **Country** | **U** | **R** | **U - R** | **p-value** | **U** | **R** | **U - R** | **p-value** | **U** | **R** | **U – R** | **p-value** | **U** | **R** | **U - R** | **p-value** |
| Afghanistan | 0.109 | 0.087 | 0.022 | 0.000 | 0.750 | 0.767 | -0.017 | 0.009 | 0.731 | 0.753 | -0.023 | 0.001 | 0.469 | 0.520 | -0.051 | 0.000 |
| Albania | 0.466 | 0.440 | 0.027 | 0.197 | 0.435 | 0.495 | -0.060 | 0.004 | 0.110 | 0.123 | -0.013 | 0.342 | 0.065 | 0.098 | -0.033 | 0.004 |
| Algeria | 0.112 | 0.093 | 0.019 | 0.000 | 0.664 | 0.687 | -0.023 | 0.000 | 0.775 | 0.784 | -0.009 | 0.103 | 0.173 | 0.190 | -0.017 | 0.002 |
| Bangladesh | 0.104 | 0.090 | 0.013 | 0.000 | 0.630 | 0.656 | -0.026 | 0.000 | 0.776 | 0.811 | -0.034 | 0.000 | 0.282 | 0.272 | 0.010 | 0.041 |
| Barbados | 0.149 | 0.157 | -0.008 | 0.770 | 0.532 | 0.544 | -0.012 | 0.743 | 0.615 | 0.576 | 0.039 | 0.273 | 0.055 | 0.057 | -0.003 | 0.879 |
| Belarus | 0.301 | 0.294 | 0.007 | 0.568 | 0.352 | 0.346 | 0.006 | 0.639 | 0.620 | 0.621 | -0.001 | 0.949 | 0.009 | 0.008 | 0.001 | 0.747 |
| Belize | 0.273 | 0.268 | 0.005 | 0.711 | 0.497 | 0.507 | -0.010 | 0.454 | 0.517 | 0.491 | 0.026 | 0.064 | 0.050 | 0.054 | -0.004 | 0.502 |
| Benin | 0.055 | 0.062 | -0.006 | 0.012 | 0.754 | 0.721 | 0.033 | 0.000 | 0.846 | 0.820 | 0.026 | 0.000 | 0.182 | 0.185 | -0.003 | 0.546 |
| Bosnia and Herzegovina | 0.481 | 0.444 | 0.037 | 0.070 | 0.268 | 0.322 | -0.054 | 0.004 | 0.351 | 0.350 | 0.000 | 0.992 | 0.040 | 0.035 | 0.005 | 0.585 |
| Burkina Faso | 0.071 | 0.063 | 0.008 | 0.634 | 0.669 | 0.585 | 0.084 | 0.004 | 0.806 | 0.843 | -0.036 | 0.157 | 0.180 | 0.211 | -0.030 | 0.237 |
| Burundi | 0.056 | 0.051 | 0.005 | 0.552 | 0.688 | 0.647 | 0.041 | 0.013 | 0.857 | 0.852 | 0.005 | 0.675 | 0.087 | 0.064 | 0.024 | 0.012 |
| Cambodia | 0.305 | 0.214 | 0.091 | 0.000 | 0.365 | 0.433 | -0.068 | 0.000 | 0.476 | 0.620 | -0.143 | 0.000 | 0.041 | 0.045 | -0.004 | 0.413 |
| Cameroon | 0.056 | 0.061 | -0.005 | 0.338 | 0.720 | 0.701 | 0.019 | 0.052 | 0.829 | 0.831 | -0.002 | 0.783 | 0.220 | 0.228 | -0.008 | 0.356 |
| Central Africa Rep. | 0.071 | 0.070 | 0.000 | 0.941 | 0.789 | 0.792 | -0.003 | 0.649 | 0.820 | 0.825 | -0.005 | 0.377 | 0.322 | 0.329 | -0.007 | 0.304 |
| Chad | 0.139 | 0.117 | 0.022 | 0.000 | 0.703 | 0.717 | -0.015 | 0.017 | 0.675 | 0.707 | -0.033 | 0.000 | 0.286 | 0.276 | 0.011 | 0.083 |
| Comoros | 0.229 | 0.247 | -0.017 | 0.143 | 0.459 | 0.423 | 0.036 | 0.008 | 0.531 | 0.518 | 0.013 | 0.347 | 0.065 | 0.064 | 0.001 | 0.854 |
| Congo | 0.088 | 0.068 | 0.021 | 0.001 | 0.643 | 0.709 | -0.067 | 0.000 | 0.722 | 0.836 | -0.114 | 0.000 | 0.231 | 0.309 | -0.078 | 0.000 |
| Congo Dem. Rep. | 0.062 | 0.063 | -0.001 | 0.802 | 0.803 | 0.758 | 0.044 | 0.000 | 0.778 | 0.764 | 0.014 | 0.038 | 0.387 | 0.330 | 0.057 | 0.000 |
| Costa Rica | 0.448 | 0.439 | 0.009 | 0.656 | 0.343 | 0.357 | -0.014 | 0.448 | 0.340 | 0.338 | 0.002 | 0.917 | 0.021 | 0.035 | -0.015 | 0.028 |
| Cote d'Ivoire | 0.110 | 0.086 | 0.024 | 0.000 | 0.600 | 0.625 | -0.025 | 0.002 | 0.747 | 0.800 | -0.053 | 0.000 | 0.154 | 0.154 | 0.000 | 0.973 |
| Cuba | 0.372 | 0.322 | 0.050 | 0.009 | 0.311 | 0.309 | 0.001 | 0.938 | 0.267 | 0.266 | 0.001 | 0.948 | 0.022 | 0.013 | 0.010 | 0.017 |
| Djibouti | 0.176 | 0.178 | -0.002 | 0.914 | 0.643 | 0.610 | 0.033 | 0.248 | 0.544 | 0.475 | 0.069 | 0.017 | 0.210 | 0.211 | -0.002 | 0.944 |
| Dominican Rep. | 0.226 | 0.211 | 0.015 | 0.031 | 0.435 | 0.460 | -0.025 | 0.003 | 0.513 | 0.520 | -0.007 | 0.435 | 0.030 | 0.039 | -0.009 | 0.007 |
| Egypt | 0.054 | 0.042 | 0.012 | 0.004 | 0.734 | 0.788 | -0.054 | 0.000 | 0.902 | 0.903 | -0.002 | 0.757 | 0.345 | 0.441 | -0.096 | 0.000 |
| El Salvador | 0.364 | 0.349 | 0.015 | 0.286 | 0.403 | 0.392 | 0.011 | 0.434 | 0.324 | 0.298 | 0.027 | 0.049 | 0.026 | 0.034 | -0.008 | 0.105 |
| Eswatini | 0.125 | 0.105 | 0.021 | 0.030 | 0.644 | 0.652 | -0.008 | 0.604 | 0.740 | 0.751 | -0.011 | 0.413 | 0.064 | 0.082 | -0.018 | 0.017 |
| Fiji | 0.202 | 0.169 | 0.033 | 0.013 | 0.615 | 0.693 | -0.078 | 0.000 | 0.595 | 0.642 | -0.047 | 0.004 | 0.111 | 0.126 | -0.015 | 0.173 |
| Gabon | 0.116 | 0.095 | 0.021 | 0.122 | 0.656 | 0.670 | -0.014 | 0.523 | 0.717 | 0.742 | -0.025 | 0.213 | 0.207 | 0.225 | -0.018 | 0.337 |
| Gambia | 0.085 | 0.081 | 0.004 | 0.321 | 0.731 | 0.730 | 0.001 | 0.885 | 0.780 | 0.782 | -0.002 | 0.758 | 0.141 | 0.195 | -0.053 | 0.000 |
| Georgia | 0.250 | 0.223 | 0.027 | 0.016 | 0.370 | 0.420 | -0.050 | 0.000 | 0.624 | 0.604 | 0.020 | 0.117 | 0.093 | 0.133 | -0.040 | 0.000 |
| Ghana | 0.049 | 0.039 | 0.010 | 0.008 | 0.754 | 0.741 | 0.013 | 0.084 | 0.862 | 0.871 | -0.009 | 0.151 | 0.139 | 0.139 | 0.000 | 0.964 |
| Guinea | 0.099 | 0.049 | 0.050 | 0.000 | 0.696 | 0.764 | -0.068 | 0.000 | 0.728 | 0.804 | -0.076 | 0.000 | 0.092 | 0.119 | -0.027 | 0.002 |
| Guinea Bissau | 0.172 | 0.196 | -0.024 | 0.000 | 0.711 | 0.688 | 0.023 | 0.005 | 0.570 | 0.530 | 0.039 | 0.000 | 0.201 | 0.197 | 0.005 | 0.525 |
| Guyana | 0.159 | 0.173 | -0.014 | 0.192 | 0.571 | 0.523 | 0.048 | 0.001 | 0.658 | 0.602 | 0.057 | 0.000 | 0.091 | 0.088 | 0.003 | 0.716 |
| Haiti | 0.094 | 0.094 | 0.000 | 0.993 | 0.767 | 0.762 | 0.005 | 0.577 | 0.621 | 0.615 | 0.006 | 0.533 | 0.162 | 0.141 | 0.021 | 0.004 |
| Honduras | 0.263 | 0.252 | 0.011 | 0.194 | 0.508 | 0.492 | 0.016 | 0.086 | 0.424 | 0.388 | 0.036 | 0.000 | 0.040 | 0.042 | -0.002 | 0.523 |
| Iraq | 0.150 | 0.122 | 0.028 | 0.000 | 0.597 | 0.654 | -0.058 | 0.000 | 0.747 | 0.780 | -0.033 | 0.000 | 0.265 | 0.296 | -0.031 | 0.000 |
| Jamaica | 0.120 | 0.116 | 0.004 | 0.667 | 0.643 | 0.643 | -0.001 | 0.938 | 0.703 | 0.693 | 0.010 | 0.390 | 0.053 | 0.051 | 0.002 | 0.767 |
| Jordan | 0.154 | 0.145 | 0.009 | 0.347 | 0.572 | 0.582 | -0.010 | 0.403 | 0.768 | 0.784 | -0.016 | 0.134 | 0.135 | 0.137 | -0.002 | 0.817 |
| Kazakhstan | 0.381 | 0.360 | 0.021 | 0.010 | 0.261 | 0.238 | 0.023 | 0.002 | 0.449 | 0.441 | 0.009 | 0.317 | 0.011 | 0.011 | 0.001 | 0.670 |
| Kiribati | 0.089 | 0.071 | 0.018 | 0.061 | 0.851 | 0.849 | 0.002 | 0.858 | 0.744 | 0.812 | -0.068 | 0.000 | 0.209 | 0.212 | -0.003 | 0.831 |
| Kosovo | 0.261 | 0.280 | -0.018 | 0.157 | 0.253 | 0.290 | -0.036 | 0.006 | 0.672 | 0.623 | 0.049 | 0.001 | 0.046 | 0.057 | -0.011 | 0.093 |

| | | | | | | | | | | | | | | | | |
|---|---|---|---|---|---|---|---|---|---|---|---|---|---|---|---|---|
| Kyrgyzstan | 0.310 | 0.329 | -0.019 | 0.068 | 0.442 | 0.408 | 0.034 | 0.002 | 0.546 | 0.534 | 0.012 | 0.265 | 0.031 | 0.039 | -0.008 | 0.046 |
| Lao People's Dem. Rep. | 0.256 | 0.217 | 0.039 | 0.000 | 0.391 | 0.392 | -0.001 | 0.886 | 0.626 | 0.666 | -0.040 | 0.000 | 0.049 | 0.059 | -0.009 | 0.001 |
| Lesotho | 0.167 | 0.174 | -0.007 | 0.571 | 0.637 | 0.614 | 0.023 | 0.156 | 0.605 | 0.570 | 0.035 | 0.034 | 0.070 | 0.065 | 0.006 | 0.488 |
| Liberia | 0.060 | 0.062 | -0.001 | 0.871 | 0.747 | 0.728 | 0.019 | 0.109 | 0.823 | 0.810 | 0.013 | 0.210 | 0.248 | 0.224 | 0.024 | 0.028 |
| Macedonia | 0.267 | 0.236 | 0.031 | 0.132 | 0.491 | 0.523 | -0.032 | 0.164 | 0.582 | 0.623 | -0.041 | 0.076 | 0.089 | 0.107 | -0.018 | 0.260 |
| Madagascar | 0.103 | 0.096 | 0.007 | 0.249 | 0.668 | 0.664 | 0.004 | 0.633 | 0.781 | 0.797 | -0.017 | 0.042 | 0.097 | 0.088 | 0.009 | 0.100 |
| Malawi | 0.161 | 0.160 | 0.001 | 0.915 | 0.559 | 0.546 | 0.012 | 0.236 | 0.696 | 0.693 | 0.003 | 0.797 | 0.108 | 0.125 | -0.017 | 0.013 |
| Mauritania | 0.120 | 0.087 | 0.034 | 0.000 | 0.662 | 0.738 | -0.076 | 0.000 | 0.707 | 0.779 | -0.072 | 0.000 | 0.234 | 0.268 | -0.034 | 0.000 |
| Moldova | 0.250 | 0.214 | 0.036 | 0.023 | 0.459 | 0.473 | -0.015 | 0.436 | 0.659 | 0.699 | -0.040 | 0.022 | 0.021 | 0.021 | 0.000 | 0.947 |
| Mongolia | 0.394 | 0.400 | -0.006 | 0.403 | 0.284 | 0.261 | 0.022 | 0.002 | 0.399 | 0.360 | 0.039 | 0.000 | 0.046 | 0.036 | 0.010 | 0.002 |
| Montenegro | 0.225 | 0.214 | 0.011 | 0.511 | 0.322 | 0.361 | -0.039 | 0.027 | 0.634 | 0.579 | 0.055 | 0.004 | 0.038 | 0.034 | 0.003 | 0.631 |
| Mozambique | 0.185 | 0.179 | 0.006 | 0.625 | 0.300 | 0.307 | -0.008 | 0.635 | 0.528 | 0.487 | 0.041 | 0.017 | 0.091 | 0.087 | 0.004 | 0.697 |
| Myanmar | 0.179 | 0.145 | 0.034 | 0.005 | 0.420 | 0.431 | -0.011 | 0.507 | 0.734 | 0.740 | -0.006 | 0.653 | 0.120 | 0.117 | 0.003 | 0.767 |
| Nepal | 0.155 | 0.132 | 0.023 | 0.000 | 0.586 | 0.574 | 0.012 | 0.204 | 0.741 | 0.776 | -0.035 | 0.000 | 0.168 | 0.161 | 0.007 | 0.313 |
| Niger | 0.142 | 0.126 | 0.016 | 0.088 | 0.659 | 0.650 | 0.010 | 0.453 | 0.739 | 0.739 | -0.000 | 0.993 | 0.287 | 0.277 | 0.010 | 0.418 |
| Nigeria | 0.063 | 0.063 | -0.000 | 0.980 | 0.769 | 0.752 | 0.017 | 0.000 | 0.811 | 0.796 | 0.015 | 0.001 | 0.274 | 0.292 | -0.018 | 0.000 |
| Panama | 0.398 | 0.387 | 0.010 | 0.644 | 0.327 | 0.284 | 0.043 | 0.041 | 0.292 | 0.249 | 0.043 | 0.031 | 0.125 | 0.116 | 0.008 | 0.604 |
| Paraguay | 0.399 | 0.424 | -0.025 | 0.187 | 0.379 | 0.355 | 0.024 | 0.186 | 0.310 | 0.284 | 0.026 | 0.148 | 0.040 | 0.038 | 0.001 | 0.879 |
| Philippines | 0.358 | 0.383 | -0.025 | 0.019 | 0.380 | 0.355 | 0.025 | 0.021 | 0.477 | 0.462 | 0.015 | 0.182 | 0.031 | 0.031 | -0.000 | 0.986 |
| St. Lucia | 0.172 | 0.198 | -0.026 | 0.430 | 0.478 | 0.417 | 0.061 | 0.148 | 0.676 | 0.569 | 0.107 | 0.008 | 0.070 | 0.047 | 0.023 | 0.254 |
| Samoa | 0.097 | 0.096 | 0.001 | 0.951 | 0.787 | 0.783 | 0.003 | 0.826 | 0.763 | 0.767 | -0.004 | 0.808 | 0.153 | 0.168 | -0.015 | 0.282 |
| Sao Tome and Principe | 0.090 | 0.092 | -0.001 | 0.887 | 0.725 | 0.710 | 0.016 | 0.261 | 0.669 | 0.639 | 0.030 | 0.041 | 0.122 | 0.106 | 0.016 | 0.094 |
| Serbia | 0.359 | 0.338 | 0.021 | 0.082 | 0.324 | 0.358 | -0.034 | 0.004 | 0.511 | 0.524 | -0.013 | 0.295 | 0.025 | 0.037 | -0.012 | 0.007 |
| Sierra Leone | 0.074 | 0.068 | 0.006 | 0.083 | 0.714 | 0.693 | 0.021 | 0.000 | 0.771 | 0.771 | -0.000 | 0.989 | 0.215 | 0.212 | 0.003 | 0.561 |
| Palestine | 0.073 | 0.068 | 0.005 | 0.156 | 0.711 | 0.726 | -0.015 | 0.019 | 0.868 | 0.875 | -0.007 | 0.108 | 0.210 | 0.217 | -0.007 | 0.240 |
| Sudan | 0.222 | 0.235 | -0.013 | 0.223 | 0.498 | 0.466 | 0.031 | 0.018 | 0.568 | 0.514 | 0.054 | 0.000 | 0.135 | 0.124 | 0.011 | 0.210 |
| Suriname | 0.112 | 0.075 | 0.037 | 0.000 | 0.564 | 0.637 | -0.073 | 0.000 | 0.789 | 0.848 | -0.059 | 0.000 | 0.072 | 0.106 | -0.034 | 0.000 |
| Syria | 0.081 | 0.067 | 0.015 | 0.002 | 0.753 | 0.758 | -0.005 | 0.522 | 0.822 | 0.837 | -0.015 | 0.026 | 0.202 | 0.231 | -0.029 | 0.000 |
| Tajikistan | 0.238 | 0.223 | 0.015 | 0.116 | 0.480 | 0.502 | -0.022 | 0.056 | 0.640 | 0.659 | -0.019 | 0.080 | 0.133 | 0.131 | 0.001 | 0.863 |
| Thailand | 0.359 | 0.321 | 0.038 | 0.000 | 0.431 | 0.454 | -0.024 | 0.009 | 0.430 | 0.495 | -0.064 | 0.000 | 0.024 | 0.027 | -0.004 | 0.171 |
| Togo | 0.070 | 0.076 | -0.006 | 0.124 | 0.717 | 0.688 | 0.029 | 0.000 | 0.811 | 0.816 | -0.005 | 0.429 | 0.161 | 0.171 | -0.010 | 0.100 |
| Tonga | 0.109 | 0.098 | 0.011 | 0.494 | 0.749 | 0.797 | -0.049 | 0.041 | 0.677 | 0.725 | -0.047 | 0.066 | 0.140 | 0.234 | -0.094 | 0.000 |
| Trinidad and Tobago | 0.213 | 0.227 | -0.014 | 0.354 | 0.443 | 0.432 | 0.011 | 0.515 | 0.627 | 0.623 | 0.004 | 0.840 | 0.029 | 0.021 | 0.008 | 0.145 |
| Tunisia | 0.106 | 0.086 | 0.021 | 0.000 | 0.661 | 0.721 | -0.060 | 0.000 | 0.814 | 0.831 | -0.017 | 0.014 | 0.176 | 0.264 | -0.088 | 0.000 |
| Turkmenistan | 0.410 | 0.429 | -0.019 | 0.079 | 0.346 | 0.380 | -0.034 | 0.002 | 0.485 | 0.435 | 0.050 | 0.000 | 0.012 | 0.008 | 0.004 | 0.120 |
| Turks and Caicos Islands | 0.142 | 0.140 | 0.002 | 0.968 | 0.609 | 0.606 | 0.003 | 0.965 | 0.679 | 0.614 | 0.065 | 0.370 | 0.047 | 0.045 | 0.002 | 0.952 |
| Tuvalu | 0.179 | 0.190 | -0.011 | 0.691 | 0.680 | 0.702 | -0.022 | 0.517 | 0.609 | 0.585 | 0.025 | 0.491 | 0.052 | 0.047 | 0.005 | 0.751 |
| Uganda | 0.115 | 0.104 | 0.011 | 0.155 | 0.651 | 0.701 | -0.050 | 0.000 | 0.692 | 0.726 | -0.034 | 0.002 | 0.125 | 0.153 | -0.028 | 0.000 |
| Ukraine | 0.337 | 0.282 | 0.055 | 0.001 | 0.310 | 0.334 | -0.024 | 0.162 | 0.584 | 0.623 | -0.040 | 0.025 | 0.013 | 0.018 | -0.005 | 0.310 |
| Uruguay | 0.370 | 0.481 | -0.111 | 0.114 | 0.222 | 0.220 | 0.002 | 0.960 | 0.483 | 0.348 | 0.135 | 0.040 | 0.030 | 0.015 | 0.015 | 0.131 |
| Uzbekistan | 0.340 | 0.307 | 0.034 | 0.053 | 0.281 | 0.305 | -0.024 | 0.170 | 0.550 | 0.592 | -0.042 | 0.022 | 0.040 | 0.045 | -0.005 | 0.509 |
| Vietnam | 0.248 | 0.216 | 0.032 | 0.000 | 0.460 | 0.472 | -0.012 | 0.131 | 0.590 | 0.639 | -0.049 | 0.000 | 0.025 | 0.032 | -0.007 | 0.010 |
| Yemen | 0.128 | 0.090 | 0.039 | 0.000 | 0.498 | 0.570 | -0.072 | 0.000 | 0.798 | 0.826 | -0.028 | 0.000 | 0.183 | 0.268 | -0.085 | 0.000 |
| Zimbabwe | 0.214 | 0.229 | -0.015 | 0.024 | 0.485 | 0.366 | 0.119 | 0.000 | 0.549 | 0.523 | 0.026 | 0.001 | 0.064 | 0.050 | 0.014 | 0.000 |
| Mean | 0.194 | 0.184 | 0.010 | | 0.552 | 0.558 | -0.006 | | 0.640 | 0.642 | -0.003 | | 0.124 | 0.133 | -0.009 | |

*Notes*: Gaps are computed as the difference between urban and rural averages. A positive value implies an urban (dis)advantage while a negative value implies rural (dis)advantage. The analysis included 86 LMICs and HICs – it excludes Argentina (all survey-waves), Qatar (all survey-waves), and Trinidad and Tobago (MICS3) due to lack of information on urban/rural. All averages were estimated using household weights. "Mean" gives the unweighted average of the 86 country-estimates.

Table B7: Country-level gap by mothers' education (Secondary/Higher [S] – Primary/Lower [P])

| | **Only non-violent** | | | | **Physical punishment** | | | | **Emotional violence** | | | | **Severe physical violence** | | | |
|---|---|---|---|---|---|---|---|---|---|---|---|---|---|---|---|---|
| **Country** | **S** | **P** | **S - P** | **p-value** | **S** | **P** | **S - P** | **p-value** | **S** | **P** | **S - P** | **p-value** | **S** | **P** | **S - P** | **p-value** |
| Afghanistan | 0.130 | 0.088 | 0.042 | 0.000 | 0.708 | 0.768 | -0.060 | 0.000 | 0.687 | 0.754 | -0.067 | 0.000 | 0.396 | 0.519 | -0.124 | 0.000 |
| Albania | 0.452 | 0.353 | 0.099 | 0.143 | 0.471 | 0.511 | -0.041 | 0.573 | 0.116 | 0.195 | -0.079 | 0.168 | 0.084 | 0.142 | -0.058 | 0.258 |
| Algeria | 0.102 | 0.111 | -0.009 | 0.023 | 0.682 | 0.657 | 0.025 | 0.000 | 0.785 | 0.768 | 0.017 | 0.001 | 0.169 | 0.197 | -0.028 | 0.000 |
| Argentina | 0.239 | 0.214 | 0.026 | 0.075 | 0.435 | 0.470 | -0.035 | 0.043 | 0.653 | 0.619 | 0.034 | 0.043 | 0.076 | 0.118 | -0.042 | 0.000 |
| Bangladesh | 0.091 | 0.095 | -0.003 | 0.187 | 0.648 | 0.653 | -0.005 | 0.211 | 0.802 | 0.804 | -0.002 | 0.577 | 0.269 | 0.280 | -0.011 | 0.003 |
| Barbados | 0.151 | 0.155 | -0.004 | 0.953 | 0.539 | 0.526 | 0.013 | 0.880 | 0.603 | 0.594 | 0.009 | 0.910 | 0.056 | 0.055 | 0.001 | 0.985 |
| Belarus | 0.299 | 0.471 | -0.172 | 0.507 | 0.350 | 0.107 | 0.244 | 0.008 | 0.620 | 0.529 | 0.091 | 0.726 | 0.008 | 0.000 | 0.008 | 0.000 |
| Belize | 0.260 | 0.279 | -0.019 | 0.141 | 0.500 | 0.504 | -0.004 | 0.769 | 0.516 | 0.492 | 0.024 | 0.093 | 0.044 | 0.058 | -0.014 | 0.013 |
| Benin | 0.062 | 0.058 | 0.003 | 0.361 | 0.749 | 0.732 | 0.018 | 0.007 | 0.820 | 0.829 | -0.008 | 0.154 | 0.153 | 0.189 | -0.036 | 0.000 |
| Bosnia and Herzegovina | 0.468 | 0.433 | 0.035 | 0.090 | 0.295 | 0.319 | -0.024 | 0.210 | 0.339 | 0.372 | -0.033 | 0.098 | 0.033 | 0.044 | -0.011 | 0.191 |
| Burkina Faso | 0.049 | 0.056 | -0.007 | 0.753 | 0.646 | 0.629 | 0.016 | 0.770 | 0.709 | 0.848 | -0.139 | 0.052 | 0.163 | 0.222 | -0.059 | 0.219 |
| Burundi | 0.047 | 0.038 | 0.009 | 0.356 | 0.720 | 0.683 | 0.037 | 0.046 | 0.839 | 0.866 | -0.028 | 0.068 | 0.053 | 0.071 | -0.018 | 0.059 |
| Cambodia | 0.289 | 0.233 | 0.056 | 0.035 | 0.423 | 0.445 | -0.023 | 0.447 | 0.484 | 0.505 | -0.021 | 0.483 | 0.017 | 0.040 | -0.023 | 0.009 |
| Cameroon | 0.055 | 0.060 | -0.005 | 0.327 | 0.711 | 0.709 | 0.002 | 0.834 | 0.821 | 0.834 | -0.013 | 0.151 | 0.190 | 0.240 | -0.050 | 0.000 |
| Central Africa Rep. | 0.078 | 0.069 | 0.009 | 0.098 | 0.793 | 0.790 | 0.003 | 0.713 | 0.807 | 0.827 | -0.020 | 0.015 | 0.314 | 0.329 | -0.015 | 0.102 |
| Chad | 0.125 | 0.118 | 0.008 | 0.236 | 0.723 | 0.720 | 0.003 | 0.743 | 0.698 | 0.706 | -0.008 | 0.370 | 0.269 | 0.284 | -0.014 | 0.098 |
| Comoros | 0.275 | 0.213 | 0.063 | 0.000 | 0.402 | 0.462 | -0.060 | 0.000 | 0.478 | 0.560 | -0.083 | 0.000 | 0.054 | 0.073 | -0.019 | 0.004 |
| Congo | 0.078 | 0.063 | 0.015 | 0.026 | 0.674 | 0.692 | -0.018 | 0.137 | 0.752 | 0.804 | -0.052 | 0.000 | 0.251 | 0.296 | -0.045 | 0.000 |
| Congo Dem. Rep. | 0.059 | 0.060 | -0.001 | 0.752 | 0.794 | 0.770 | 0.024 | 0.000 | 0.777 | 0.771 | 0.007 | 0.288 | 0.362 | 0.349 | 0.013 | 0.098 |
| Costa Rica | 0.435 | 0.469 | -0.035 | 0.100 | 0.364 | 0.308 | 0.056 | 0.004 | 0.352 | 0.310 | 0.042 | 0.035 | 0.026 | 0.026 | -0.000 | 0.962 |
| Cote d'Ivoire | 0.098 | 0.079 | 0.019 | 0.025 | 0.607 | 0.676 | -0.069 | 0.000 | 0.787 | 0.825 | -0.037 | 0.003 | 0.128 | 0.177 | -0.049 | 0.000 |
| Cuba | 0.357 | 0.320 | 0.038 | 0.409 | 0.312 | 0.253 | 0.059 | 0.150 | 0.265 | 0.321 | -0.056 | 0.240 | 0.019 | 0.017 | 0.003 | 0.711 |
| Djibouti | 0.246 | 0.164 | 0.081 | 0.004 | 0.567 | 0.654 | -0.088 | 0.007 | 0.472 | 0.553 | -0.081 | 0.014 | 0.162 | 0.218 | -0.056 | 0.032 |
| Dominican Rep. | 0.231 | 0.206 | 0.025 | 0.000 | 0.435 | 0.454 | -0.019 | 0.025 | 0.512 | 0.522 | -0.011 | 0.197 | 0.027 | 0.043 | -0.015 | 0.000 |
| Egypt | 0.044 | 0.046 | -0.002 | 0.591 | 0.769 | 0.778 | -0.009 | 0.294 | 0.906 | 0.902 | 0.004 | 0.458 | 0.381 | 0.465 | -0.084 | 0.000 |
| El Salvador | 0.366 | 0.349 | 0.016 | 0.257 | 0.419 | 0.376 | 0.043 | 0.003 | 0.317 | 0.310 | 0.006 | 0.644 | 0.027 | 0.031 | -0.004 | 0.418 |
| Eswatini | 0.117 | 0.100 | 0.017 | 0.013 | 0.653 | 0.645 | 0.008 | 0.459 | 0.735 | 0.767 | -0.032 | 0.001 | 0.072 | 0.085 | -0.012 | 0.044 |
| Fiji | 0.184 | 0.210 | -0.025 | 0.236 | 0.658 | 0.598 | 0.060 | 0.018 | 0.614 | 0.632 | -0.018 | 0.456 | 0.116 | 0.132 | -0.016 | 0.363 |
| Gabon | 0.085 | 0.122 | -0.037 | 0.358 | 0.688 | 0.586 | 0.102 | 0.073 | 0.693 | 0.611 | 0.083 | 0.144 | 0.187 | 0.159 | 0.028 | 0.494 |
| Gambia | 0.096 | 0.079 | 0.017 | 0.005 | 0.714 | 0.736 | -0.022 | 0.019 | 0.764 | 0.786 | -0.022 | 0.011 | 0.127 | 0.175 | -0.048 | 0.000 |
| Georgia | 0.241 | 0.167 | 0.073 | 0.001 | 0.392 | 0.397 | -0.005 | 0.866 | 0.609 | 0.771 | -0.162 | 0.000 | 0.112 | 0.089 | 0.023 | 0.185 |
| Ghana | 0.052 | 0.036 | 0.015 | 0.000 | 0.755 | 0.739 | 0.016 | 0.033 | 0.856 | 0.876 | -0.020 | 0.001 | 0.128 | 0.148 | -0.019 | 0.001 |
| Guinea | 0.104 | 0.060 | 0.044 | 0.000 | 0.651 | 0.755 | -0.104 | 0.000 | 0.711 | 0.788 | -0.077 | 0.000 | 0.062 | 0.117 | -0.055 | 0.000 |
| Guinea Bissau | 0.178 | 0.189 | -0.011 | 0.307 | 0.712 | 0.694 | 0.018 | 0.139 | 0.560 | 0.542 | 0.018 | 0.193 | 0.181 | 0.201 | -0.020 | 0.066 |
| Guyana | 0.171 | 0.162 | 0.009 | 0.434 | 0.537 | 0.531 | 0.006 | 0.681 | 0.619 | 0.612 | 0.007 | 0.649 | 0.082 | 0.108 | -0.026 | 0.003 |
| Haiti | 0.074 | 0.062 | 0.012 | 0.329 | 0.798 | 0.806 | -0.008 | 0.685 | 0.538 | 0.558 | -0.020 | 0.426 | 0.100 | 0.101 | -0.001 | 0.935 |
| Honduras | 0.258 | 0.256 | 0.002 | 0.829 | 0.523 | 0.482 | 0.040 | 0.000 | 0.420 | 0.392 | 0.028 | 0.003 | 0.036 | 0.044 | -0.008 | 0.021 |
| Iraq | 0.152 | 0.136 | 0.016 | 0.001 | 0.578 | 0.634 | -0.056 | 0.000 | 0.746 | 0.762 | -0.016 | 0.006 | 0.233 | 0.297 | -0.064 | 0.000 |
| Jamaica | 0.119 | 0.113 | 0.006 | 0.696 | 0.643 | 0.643 | 0.000 | 0.994 | 0.698 | 0.699 | -0.001 | 0.953 | 0.052 | 0.055 | -0.003 | 0.779 |

| | | | | | | | | | | | | | | | | |
|---|---|---|---|---|---|---|---|---|---|---|---|---|---|---|---|---|
| Jordan | 0.160 | 0.087 | 0.072 | 0.000 | 0.605 | 0.702 | -0.098 | 0.006 | 0.731 | 0.809 | -0.079 | 0.009 | 0.118 | 0.176 | -0.058 | 0.062 |
| Kazakhstan | 0.374 | 0.270 | 0.105 | 0.000 | 0.249 | 0.283 | -0.034 | 0.090 | 0.443 | 0.506 | -0.063 | 0.005 | 0.011 | 0.011 | -0.000 | 0.949 |
| Kiribati | 0.083 | 0.069 | 0.014 | 0.202 | 0.844 | 0.870 | -0.026 | 0.062 | 0.763 | 0.830 | -0.067 | 0.000 | 0.193 | 0.272 | -0.079 | 0.000 |
| Kosovo | 0.273 | 0.268 | 0.005 | 0.784 | 0.264 | 0.340 | -0.077 | 0.000 | 0.643 | 0.641 | 0.001 | 0.939 | 0.046 | 0.091 | -0.045 | 0.000 |
| Kyrgyzstan | 0.323 | 0.276 | 0.047 | 0.235 | 0.419 | 0.483 | -0.064 | 0.183 | 0.539 | 0.501 | 0.038 | 0.433 | 0.036 | 0.043 | -0.007 | 0.655 |
| Lao People’s Dem. Rep. | 0.252 | 0.217 | 0.035 | 0.000 | 0.386 | 0.394 | -0.007 | 0.229 | 0.629 | 0.667 | -0.037 | 0.000 | 0.041 | 0.063 | -0.022 | 0.000 |
| Lesotho | 0.145 | 0.194 | -0.049 | 0.000 | 0.655 | 0.594 | 0.061 | 0.000 | 0.592 | 0.575 | 0.018 | 0.237 | 0.067 | 0.067 | 0.001 | 0.912 |
| Liberia | 0.056 | 0.053 | 0.003 | 0.794 | 0.729 | 0.757 | -0.028 | 0.109 | 0.806 | 0.823 | -0.017 | 0.277 | 0.215 | 0.253 | -0.038 | 0.015 |
| Macedonia | 0.288 | 0.209 | 0.079 | 0.000 | 0.476 | 0.541 | -0.065 | 0.005 | 0.567 | 0.640 | -0.073 | 0.002 | 0.075 | 0.123 | -0.048 | 0.003 |
| Madagascar | 0.106 | 0.095 | 0.012 | 0.066 | 0.677 | 0.661 | 0.016 | 0.103 | 0.774 | 0.801 | -0.027 | 0.003 | 0.084 | 0.092 | -0.008 | 0.207 |
| Malawi | 0.169 | 0.158 | 0.011 | 0.104 | 0.556 | 0.547 | 0.009 | 0.297 | 0.679 | 0.697 | -0.018 | 0.028 | 0.109 | 0.126 | -0.017 | 0.002 |
| Mauritania | 0.139 | 0.095 | 0.044 | 0.000 | 0.622 | 0.720 | -0.098 | 0.000 | 0.667 | 0.762 | -0.095 | 0.000 | 0.192 | 0.263 | -0.071 | 0.000 |
| Moldova | 0.229 | 0.097 | 0.131 | 0.020 | 0.467 | 0.632 | -0.165 | 0.112 | 0.683 | 0.739 | -0.056 | 0.540 | 0.020 | 0.144 | -0.124 | 0.107 |
| Mongolia | 0.397 | 0.386 | 0.011 | 0.302 | 0.276 | 0.273 | 0.003 | 0.795 | 0.388 | 0.360 | 0.028 | 0.010 | 0.042 | 0.041 | 0.002 | 0.729 |
| Montenegro | 0.232 | 0.172 | 0.060 | 0.002 | 0.318 | 0.419 | -0.101 | 0.000 | 0.609 | 0.640 | -0.031 | 0.208 | 0.032 | 0.056 | -0.024 | 0.039 |
| Mozambique | 0.173 | 0.139 | 0.034 | 0.338 | 0.265 | 0.241 | 0.024 | 0.588 | 0.389 | 0.438 | -0.049 | 0.343 | 0.068 | 0.079 | -0.011 | 0.641 |
| Myanmar | 0.159 | 0.084 | 0.075 | 0.000 | 0.490 | 0.587 | -0.097 | 0.002 | 0.743 | 0.792 | -0.049 | 0.062 | 0.111 | 0.148 | -0.037 | 0.088 |
| Nepal | 0.169 | 0.122 | 0.047 | 0.000 | 0.564 | 0.593 | -0.029 | 0.002 | 0.720 | 0.791 | -0.071 | 0.000 | 0.134 | 0.187 | -0.052 | 0.000 |
| Niger | 0.136 | 0.124 | 0.012 | 0.547 | 0.673 | 0.668 | 0.005 | 0.853 | 0.719 | 0.747 | -0.029 | 0.268 | 0.299 | 0.283 | 0.015 | 0.577 |
| Nigeria | 0.056 | 0.067 | -0.011 | 0.000 | 0.796 | 0.735 | 0.060 | 0.000 | 0.821 | 0.789 | 0.033 | 0.000 | 0.281 | 0.288 | -0.007 | 0.110 |
| Panama | 0.398 | 0.386 | 0.013 | 0.619 | 0.321 | 0.287 | 0.035 | 0.127 | 0.289 | 0.249 | 0.040 | 0.060 | 0.119 | 0.127 | -0.008 | 0.650 |
| Paraguay | 0.417 | 0.395 | 0.023 | 0.263 | 0.382 | 0.353 | 0.029 | 0.141 | 0.293 | 0.313 | -0.021 | 0.268 | 0.032 | 0.050 | -0.018 | 0.036 |
| Philippines | 0.362 | 0.368 | -0.006 | 0.702 | 0.377 | 0.382 | -0.005 | 0.730 | 0.474 | 0.461 | 0.013 | 0.388 | 0.028 | 0.045 | -0.017 | 0.007 |
| Qatar | 0.432 | 0.348 | 0.084 | 0.019 | 0.302 | 0.414 | -0.111 | 0.001 | 0.376 | 0.501 | -0.125 | 0.001 | 0.053 | 0.099 | -0.046 | 0.052 |
| St. Lucia | 0.200 | 0.183 | 0.017 | 0.642 | 0.434 | 0.421 | 0.013 | 0.775 | 0.573 | 0.615 | -0.042 | 0.345 | 0.055 | 0.046 | 0.009 | 0.642 |
| Samoa | 0.097 | 0.077 | 0.021 | 0.311 | 0.782 | 0.806 | -0.023 | 0.433 | 0.762 | 0.829 | -0.067 | 0.014 | 0.161 | 0.238 | -0.076 | 0.013 |
| Sao Tome and Principe | 0.108 | 0.079 | 0.029 | 0.003 | 0.708 | 0.730 | -0.022 | 0.125 | 0.652 | 0.665 | -0.012 | 0.428 | 0.088 | 0.135 | -0.046 | 0.000 |
| Serbia | 0.364 | 0.272 | 0.092 | 0.000 | 0.324 | 0.419 | -0.095 | 0.000 | 0.504 | 0.589 | -0.085 | 0.000 | 0.026 | 0.052 | -0.025 | 0.000 |
| Sierra Leone | 0.071 | 0.069 | 0.002 | 0.677 | 0.708 | 0.698 | 0.010 | 0.197 | 0.759 | 0.774 | -0.015 | 0.033 | 0.194 | 0.217 | -0.023 | 0.001 |
| Palestine | 0.074 | 0.070 | 0.005 | 0.190 | 0.706 | 0.727 | -0.022 | 0.000 | 0.863 | 0.878 | -0.015 | 0.001 | 0.188 | 0.243 | -0.055 | 0.000 |
| Sudan | 0.230 | 0.232 | -0.002 | 0.903 | 0.474 | 0.476 | -0.002 | 0.909 | 0.567 | 0.520 | 0.047 | 0.002 | 0.087 | 0.138 | -0.051 | 0.000 |
| Suriname | 0.110 | 0.080 | 0.030 | 0.000 | 0.562 | 0.636 | -0.074 | 0.000 | 0.799 | 0.829 | -0.030 | 0.000 | 0.066 | 0.116 | -0.050 | 0.000 |
| Syria | 0.085 | 0.067 | 0.018 | 0.000 | 0.738 | 0.767 | -0.029 | 0.000 | 0.822 | 0.834 | -0.012 | 0.075 | 0.169 | 0.248 | -0.079 | 0.000 |
| Tajikistan | 0.225 | 0.203 | 0.022 | 0.311 | 0.498 | 0.561 | -0.063 | 0.016 | 0.656 | 0.687 | -0.031 | 0.200 | 0.133 | 0.141 | -0.008 | 0.652 |
| Thailand | 0.367 | 0.295 | 0.073 | 0.000 | 0.436 | 0.457 | -0.021 | 0.016 | 0.421 | 0.533 | -0.113 | 0.000 | 0.021 | 0.033 | -0.012 | 0.000 |
| Togo | 0.082 | 0.066 | 0.015 | 0.005 | 0.708 | 0.714 | -0.006 | 0.517 | 0.782 | 0.828 | -0.046 | 0.000 | 0.140 | 0.183 | -0.043 | 0.000 |
| Tonga | 0.099 | 0.133 | -0.034 | 0.545 | 0.789 | 0.735 | 0.054 | 0.427 | 0.716 | 0.685 | 0.032 | 0.662 | 0.215 | 0.245 | -0.030 | 0.650 |
| Trinidad and Tobago | 0.207 | 0.190 | 0.017 | 0.226 | 0.458 | 0.489 | -0.031 | 0.073 | 0.633 | 0.655 | -0.022 | 0.196 | 0.028 | 0.048 | -0.020 | 0.004 |
| Tunisia | 0.103 | 0.094 | 0.009 | 0.084 | 0.676 | 0.688 | -0.012 | 0.165 | 0.811 | 0.831 | -0.019 | 0.005 | 0.163 | 0.264 | -0.101 | 0.000 |
| Turkmenistan | 0.421 | 0.510 | -0.089 | 0.728 | 0.366 | 0.490 | -0.124 | 0.627 | 0.455 | 0.188 | 0.267 | 0.132 | 0.010 | 0.000 | 0.010 | 0.000 |
| Tuvalu | 0.179 | 0.213 | -0.034 | 0.363 | 0.706 | 0.624 | 0.081 | 0.069 | 0.602 | 0.593 | 0.009 | 0.851 | 0.047 | 0.056 | -0.009 | 0.678 |
| Uganda | 0.112 | 0.081 | 0.030 | 0.000 | 0.662 | 0.751 | -0.088 | 0.000 | 0.684 | 0.749 | -0.065 | 0.000 | 0.120 | 0.177 | -0.057 | 0.000 |
| Ukraine | 0.319 | 1.000 | -0.681 | 0.000 | 0.318 | 0.000 | 0.318 | 0.000 | 0.596 | 0.000 | 0.596 | 0.000 | 0.015 | 0.000 | 0.015 | 0.000 |
| Uruguay | 0.389 | 0.359 | 0.030 | 0.777 | 0.213 | 0.244 | -0.032 | 0.591 | 0.457 | 0.501 | -0.044 | 0.656 | 0.031 | 0.022 | 0.009 | 0.568 |
| Uzbekistan | 0.322 | 0.292 | 0.029 | 0.853 | 0.294 | 0.368 | -0.074 | 0.635 | 0.573 | 0.708 | -0.135 | 0.393 | 0.043 | 0.193 | -0.151 | 0.230 |
| Vietnam | 0.246 | 0.169 | 0.077 | 0.000 | 0.455 | 0.507 | -0.052 | 0.000 | 0.589 | 0.720 | -0.131 | 0.000 | 0.021 | 0.054 | -0.034 | 0.000 |
| Yemen | 0.132 | 0.089 | 0.044 | 0.000 | 0.460 | 0.584 | -0.125 | 0.000 | 0.784 | 0.832 | -0.048 | 0.000 | 0.117 | 0.290 | -0.174 | 0.000 |
| Zimbabwe | 0.218 | 0.234 | -0.016 | 0.011 | 0.438 | 0.349 | 0.089 | 0.000 | 0.534 | 0.526 | 0.007 | 0.307 | 0.057 | 0.051 | 0.006 | 0.075 |

| | | | | | | | | | | | | | | | |
|---|---|---|---|---|---|---|---|---|---|---|---|---|---|---|---|
| Mean | 0.197 | 0.186 | 0.011 | | 0.549 | 0.559 | -0.011 | | 0.626 | 0.642 | -0.017 | | 0.112 | 0.144 | -0.031 |

*Notes*: Gaps are computed as the difference between secondary/higher and primary/lower education averages. A positive value implies secondary/higher education (dis)advantage while a negative value implies primary/lower education (dis)advantage. The analysis included 87 LMICs and HICs – it excludes Turks and Caicos Island (all survey-waves) due to lack of variation in both education categories. All averages were estimated using household weights. "Mean" gives the unweighted average of the 87 country-estimates.

Table B8: Aggregate-level gap by wealth (Richest [T] – Poorest [B])

| | Only non-violent | | | | Physical punishment | | | | Emotional violence | | | | Severe physical violence | | | |
|---|---|---|---|---|---|---|---|---|---|---|---|---|---|---|---|---|
| **Country** | **T** | **B** | **T - B** | **p-value** | **T** | **B** | **T - B** | **p-value** | **T** | **B** | **T - B** | **p-value** | **T** | **B** | **T – B** | **p-value** |
| Afghanistan | 0.122 | 0.072 | 0.050 | 0.000 | 0.725 | 0.773 | -0.048 | 0.000 | 0.711 | 0.742 | -0.032 | 0.000 | 0.440 | 0.542 | -0.102 | 0.000 |
| Albania | 0.509 | 0.361 | 0.147 | 0.000 | 0.407 | 0.552 | -0.145 | 0.000 | 0.094 | 0.151 | -0.057 | 0.010 | 0.051 | 0.106 | -0.055 | 0.002 |
| Algeria | 0.140 | 0.099 | 0.041 | 0.000 | 0.605 | 0.688 | -0.084 | 0.000 | 0.763 | 0.760 | 0.003 | 0.756 | 0.132 | 0.219 | -0.087 | 0.000 |
| Argentina | 0.295 | 0.172 | 0.122 | 0.000 | 0.340 | 0.536 | -0.196 | 0.000 | 0.613 | 0.672 | -0.059 | 0.008 | 0.065 | 0.125 | -0.060 | 0.000 |
| Bangladesh | 0.120 | 0.079 | 0.040 | 0.000 | 0.578 | 0.698 | -0.120 | 0.000 | 0.751 | 0.828 | -0.077 | 0.000 | 0.235 | 0.303 | -0.068 | 0.000 |
| Barbados | 0.205 | 0.146 | 0.059 | 0.181 | 0.492 | 0.564 | -0.072 | 0.221 | 0.605 | 0.636 | -0.031 | 0.589 | 0.054 | 0.103 | -0.048 | 0.153 |
| Belarus | 0.321 | 0.291 | 0.031 | 0.127 | 0.335 | 0.353 | -0.018 | 0.341 | 0.604 | 0.631 | -0.027 | 0.185 | 0.010 | 0.005 | 0.005 | 0.180 |
| Belize | 0.321 | 0.222 | 0.099 | 0.000 | 0.418 | 0.581 | -0.163 | 0.000 | 0.468 | 0.528 | -0.061 | 0.006 | 0.033 | 0.082 | -0.049 | 0.000 |
| Benin | 0.050 | 0.070 | -0.020 | 0.000 | 0.771 | 0.676 | 0.096 | 0.000 | 0.857 | 0.801 | 0.055 | 0.000 | 0.163 | 0.175 | -0.013 | 0.057 |
| Bosnia and Herzegovina | 0.522 | 0.478 | 0.045 | 0.156 | 0.234 | 0.324 | -0.090 | 0.001 | 0.335 | 0.327 | 0.008 | 0.788 | 0.017 | 0.048 | -0.030 | 0.012 |
| Burkina Faso | 0.071 | 0.062 | 0.009 | 0.639 | 0.684 | 0.584 | 0.099 | 0.003 | 0.807 | 0.847 | -0.039 | 0.185 | 0.189 | 0.237 | -0.048 | 0.126 |
| Burundi | 0.061 | 0.049 | 0.012 | 0.137 | 0.655 | 0.650 | 0.005 | 0.758 | 0.840 | 0.844 | -0.004 | 0.752 | 0.072 | 0.059 | 0.013 | 0.118 |
| Cambodia | 0.368 | 0.198 | 0.170 | 0.000 | 0.333 | 0.469 | -0.137 | 0.000 | 0.436 | 0.650 | -0.214 | 0.000 | 0.028 | 0.054 | -0.026 | 0.000 |
| Cameroon | 0.063 | 0.078 | -0.015 | 0.105 | 0.687 | 0.676 | 0.011 | 0.487 | 0.815 | 0.790 | 0.025 | 0.083 | 0.188 | 0.242 | -0.054 | 0.000 |
| Central Africa Rep. | 0.073 | 0.068 | 0.005 | 0.407 | 0.782 | 0.798 | -0.017 | 0.094 | 0.807 | 0.832 | -0.025 | 0.010 | 0.303 | 0.339 | -0.036 | 0.002 |
| Chad | 0.140 | 0.114 | 0.026 | 0.000 | 0.708 | 0.704 | 0.004 | 0.619 | 0.677 | 0.699 | -0.022 | 0.006 | 0.291 | 0.270 | 0.021 | 0.008 |
| Comoros | 0.306 | 0.202 | 0.104 | 0.000 | 0.368 | 0.446 | -0.078 | 0.000 | 0.420 | 0.562 | -0.142 | 0.000 | 0.049 | 0.060 | -0.011 | 0.253 |
| Congo | 0.109 | 0.061 | 0.048 | 0.000 | 0.604 | 0.723 | -0.119 | 0.000 | 0.702 | 0.845 | -0.143 | 0.000 | 0.205 | 0.322 | -0.117 | 0.000 |
| Congo Dem. Rep. | 0.061 | 0.061 | 0.000 | 0.944 | 0.808 | 0.748 | 0.060 | 0.000 | 0.781 | 0.760 | 0.021 | 0.041 | 0.386 | 0.328 | 0.058 | 0.000 |
| Costa Rica | 0.451 | 0.440 | 0.011 | 0.773 | 0.312 | 0.360 | -0.048 | 0.165 | 0.359 | 0.310 | 0.049 | 0.164 | 0.023 | 0.038 | -0.015 | 0.180 |
| Cote d'Ivoire | 0.122 | 0.089 | 0.034 | 0.000 | 0.588 | 0.624 | -0.036 | 0.007 | 0.733 | 0.796 | -0.063 | 0.000 | 0.140 | 0.161 | -0.021 | 0.036 |
| Cuba | 0.413 | 0.301 | 0.112 | 0.001 | 0.249 | 0.408 | -0.159 | 0.000 | 0.249 | 0.350 | -0.102 | 0.001 | 0.014 | 0.025 | -0.011 | 0.165 |
| Dominican Rep. | 0.250 | 0.189 | 0.060 | 0.000 | 0.381 | 0.491 | -0.110 | 0.000 | 0.516 | 0.495 | 0.021 | 0.100 | 0.023 | 0.041 | -0.018 | 0.000 |
| Egypt | 0.060 | 0.047 | 0.013 | 0.048 | 0.698 | 0.782 | -0.084 | 0.000 | 0.888 | 0.897 | -0.009 | 0.312 | 0.287 | 0.470 | -0.184 | 0.000 |
| El Salvador | 0.387 | 0.356 | 0.031 | 0.197 | 0.380 | 0.383 | -0.003 | 0.898 | 0.326 | 0.268 | 0.058 | 0.009 | 0.027 | 0.031 | -0.005 | 0.555 |
| Eswatini | 0.152 | 0.094 | 0.058 | 0.000 | 0.589 | 0.663 | -0.073 | 0.000 | 0.712 | 0.764 | -0.052 | 0.001 | 0.053 | 0.086 | -0.033 | 0.000 |
| Fiji | 0.295 | 0.128 | 0.167 | 0.000 | 0.439 | 0.761 | -0.323 | 0.000 | 0.542 | 0.673 | -0.132 | 0.000 | 0.072 | 0.158 | -0.086 | 0.000 |
| Gabon | 0.166 | 0.089 | 0.077 | 0.004 | 0.584 | 0.701 | -0.117 | 0.001 | 0.618 | 0.778 | -0.159 | 0.000 | 0.113 | 0.252 | -0.139 | 0.000 |
| Gambia | 0.116 | 0.069 | 0.047 | 0.000 | 0.685 | 0.738 | -0.053 | 0.000 | 0.744 | 0.798 | -0.054 | 0.000 | 0.136 | 0.183 | -0.047 | 0.000 |
| Georgia | 0.276 | 0.218 | 0.058 | 0.003 | 0.345 | 0.443 | -0.098 | 0.000 | 0.606 | 0.619 | -0.013 | 0.537 | 0.075 | 0.137 | -0.062 | 0.000 |
| Ghana | 0.064 | 0.037 | 0.026 | 0.000 | 0.725 | 0.736 | -0.011 | 0.352 | 0.853 | 0.866 | -0.013 | 0.146 | 0.113 | 0.159 | -0.046 | 0.000 |
| Guinea | 0.099 | 0.036 | 0.063 | 0.000 | 0.674 | 0.812 | -0.139 | 0.000 | 0.728 | 0.836 | -0.108 | 0.000 | 0.078 | 0.149 | -0.071 | 0.000 |
| Guinea Bissau | 0.181 | 0.161 | 0.020 | 0.050 | 0.708 | 0.720 | -0.012 | 0.304 | 0.577 | 0.583 | -0.005 | 0.685 | 0.194 | 0.192 | 0.001 | 0.894 |
| Guyana | 0.186 | 0.144 | 0.042 | 0.004 | 0.499 | 0.583 | -0.083 | 0.000 | 0.609 | 0.640 | -0.031 | 0.102 | 0.060 | 0.112 | -0.052 | 0.000 |
| Haiti | 0.118 | 0.103 | 0.015 | 0.132 | 0.742 | 0.748 | -0.006 | 0.673 | 0.604 | 0.609 | -0.005 | 0.759 | 0.143 | 0.137 | 0.006 | 0.568 |
| Honduras | 0.314 | 0.236 | 0.078 | 0.000 | 0.463 | 0.493 | -0.030 | 0.055 | 0.403 | 0.372 | 0.031 | 0.039 | 0.040 | 0.052 | -0.013 | 0.041 |
| Iraq | 0.207 | 0.113 | 0.094 | 0.000 | 0.516 | 0.642 | -0.127 | 0.000 | 0.691 | 0.774 | -0.082 | 0.000 | 0.171 | 0.340 | -0.170 | 0.000 |

| | | | | | | | | | | | | | | | | |
|---|---|---|---|---|---|---|---|---|---|---|---|---|---|---|---|---|
| Jamaica | 0.199 | 0.081 | 0.119 | 0.000 | 0.482 | 0.693 | -0.211 | 0.000 | 0.617 | 0.713 | -0.097 | 0.000 | 0.024 | 0.072 | -0.047 | 0.000 |
| Jordan | 0.235 | 0.132 | 0.103 | 0.000 | 0.450 | 0.626 | -0.176 | 0.000 | 0.674 | 0.782 | -0.109 | 0.000 | 0.088 | 0.165 | -0.077 | 0.000 |
| Kazakhstan | 0.407 | 0.339 | 0.069 | 0.000 | 0.263 | 0.253 | 0.010 | 0.386 | 0.449 | 0.469 | -0.020 | 0.117 | 0.009 | 0.013 | -0.004 | 0.122 |
| Kiribati | 0.136 | 0.064 | 0.072 | 0.000 | 0.767 | 0.857 | -0.090 | 0.000 | 0.680 | 0.825 | -0.145 | 0.000 | 0.145 | 0.252 | -0.107 | 0.000 |
| Kosovo | 0.287 | 0.266 | 0.021 | 0.310 | 0.200 | 0.324 | -0.124 | 0.000 | 0.634 | 0.630 | 0.004 | 0.837 | 0.027 | 0.083 | -0.056 | 0.000 |
| Kyrgyzstan | 0.298 | 0.354 | -0.056 | 0.000 | 0.441 | 0.398 | 0.042 | 0.012 | 0.563 | 0.494 | 0.069 | 0.000 | 0.030 | 0.026 | 0.004 | 0.456 |
| Lao People's Dem. Rep. | 0.280 | 0.208 | 0.073 | 0.000 | 0.362 | 0.411 | -0.049 | 0.000 | 0.596 | 0.665 | -0.069 | 0.000 | 0.034 | 0.082 | -0.048 | 0.000 |
| Lesotho | 0.164 | 0.180 | -0.017 | 0.355 | 0.634 | 0.612 | 0.021 | 0.362 | 0.590 | 0.557 | 0.032 | 0.171 | 0.073 | 0.067 | 0.006 | 0.627 |
| Liberia | 0.058 | 0.060 | -0.002 | 0.809 | 0.744 | 0.709 | 0.035 | 0.060 | 0.816 | 0.791 | 0.025 | 0.144 | 0.237 | 0.210 | 0.027 | 0.113 |
| Macedonia | 0.341 | 0.216 | 0.125 | 0.000 | 0.396 | 0.536 | -0.140 | 0.000 | 0.523 | 0.650 | -0.127 | 0.000 | 0.044 | 0.162 | -0.118 | 0.000 |
| Madagascar | 0.101 | 0.096 | 0.004 | 0.629 | 0.667 | 0.665 | 0.003 | 0.851 | 0.768 | 0.793 | -0.026 | 0.045 | 0.089 | 0.086 | 0.003 | 0.745 |
| Malawi | 0.176 | 0.146 | 0.030 | 0.000 | 0.532 | 0.568 | -0.036 | 0.001 | 0.696 | 0.695 | 0.001 | 0.942 | 0.109 | 0.131 | -0.022 | 0.002 |
| Mauritania | 0.138 | 0.078 | 0.060 | 0.000 | 0.629 | 0.797 | -0.168 | 0.000 | 0.678 | 0.819 | -0.141 | 0.000 | 0.191 | 0.290 | -0.100 | 0.000 |
| Moldova | 0.255 | 0.164 | 0.091 | 0.000 | 0.459 | 0.566 | -0.107 | 0.001 | 0.661 | 0.769 | -0.108 | 0.000 | 0.016 | 0.037 | -0.021 | 0.046 |
| Mongolia | 0.406 | 0.401 | 0.005 | 0.681 | 0.293 | 0.267 | 0.026 | 0.024 | 0.384 | 0.336 | 0.048 | 0.000 | 0.043 | 0.035 | 0.008 | 0.138 |
| Montenegro | 0.253 | 0.187 | 0.066 | 0.026 | 0.272 | 0.426 | -0.154 | 0.000 | 0.627 | 0.617 | 0.010 | 0.752 | 0.034 | 0.048 | -0.014 | 0.255 |
| Mozambique | 0.222 | 0.157 | 0.064 | 0.002 | 0.274 | 0.278 | -0.004 | 0.858 | 0.542 | 0.446 | 0.096 | 0.000 | 0.069 | 0.079 | -0.009 | 0.476 |
| Myanmar | 0.195 | 0.113 | 0.081 | 0.000 | 0.350 | 0.507 | -0.156 | 0.000 | 0.710 | 0.762 | -0.051 | 0.008 | 0.099 | 0.151 | -0.051 | 0.000 |
| Nepal | 0.202 | 0.119 | 0.083 | 0.000 | 0.498 | 0.607 | -0.109 | 0.000 | 0.689 | 0.773 | -0.084 | 0.000 | 0.109 | 0.193 | -0.084 | 0.000 |
| Niger | 0.136 | 0.138 | -0.001 | 0.909 | 0.666 | 0.635 | 0.032 | 0.065 | 0.753 | 0.728 | 0.025 | 0.109 | 0.285 | 0.289 | -0.003 | 0.833 |
| Nigeria | 0.068 | 0.071 | -0.003 | 0.436 | 0.765 | 0.710 | 0.055 | 0.000 | 0.813 | 0.759 | 0.053 | 0.000 | 0.263 | 0.267 | -0.004 | 0.572 |
| Panama | 0.453 | 0.364 | 0.088 | 0.034 | 0.262 | 0.288 | -0.026 | 0.447 | 0.265 | 0.289 | -0.024 | 0.477 | 0.113 | 0.095 | 0.018 | 0.443 |
| Paraguay | 0.436 | 0.418 | 0.018 | 0.581 | 0.309 | 0.392 | -0.083 | 0.006 | 0.311 | 0.273 | 0.038 | 0.200 | 0.033 | 0.050 | -0.017 | 0.173 |
| Philippines | 0.415 | 0.387 | 0.028 | 0.112 | 0.299 | 0.376 | -0.077 | 0.000 | 0.428 | 0.444 | -0.016 | 0.358 | 0.021 | 0.039 | -0.019 | 0.003 |
| St. Lucia | 0.193 | 0.167 | 0.027 | 0.614 | 0.335 | 0.517 | -0.182 | 0.008 | 0.470 | 0.683 | -0.212 | 0.002 | 0.036 | 0.055 | -0.019 | 0.511 |
| Samoa | 0.136 | 0.076 | 0.060 | 0.000 | 0.720 | 0.819 | -0.099 | 0.000 | 0.713 | 0.783 | -0.070 | 0.002 | 0.130 | 0.217 | -0.087 | 0.000 |
| Sao Tome and Principe | 0.140 | 0.064 | 0.076 | 0.000 | 0.626 | 0.762 | -0.135 | 0.000 | 0.648 | 0.648 | 0.001 | 0.974 | 0.070 | 0.146 | -0.076 | 0.000 |
| Serbia | 0.398 | 0.293 | 0.105 | 0.000 | 0.281 | 0.386 | -0.106 | 0.000 | 0.483 | 0.559 | -0.076 | 0.000 | 0.020 | 0.066 | -0.045 | 0.000 |
| Sierra Leone | 0.067 | 0.069 | -0.002 | 0.679 | 0.731 | 0.681 | 0.050 | 0.000 | 0.782 | 0.767 | 0.015 | 0.064 | 0.208 | 0.212 | -0.003 | 0.700 |
| Palestine | 0.091 | 0.059 | 0.032 | 0.000 | 0.633 | 0.793 | -0.160 | 0.000 | 0.851 | 0.881 | -0.030 | 0.000 | 0.127 | 0.306 | -0.179 | 0.000 |
| Sudan | 0.204 | 0.259 | -0.055 | 0.001 | 0.470 | 0.427 | 0.043 | 0.028 | 0.628 | 0.413 | 0.215 | 0.000 | 0.095 | 0.125 | -0.030 | 0.012 |
| Suriname | 0.162 | 0.049 | 0.113 | 0.000 | 0.479 | 0.708 | -0.230 | 0.000 | 0.743 | 0.875 | -0.132 | 0.000 | 0.052 | 0.122 | -0.071 | 0.000 |
| Syria | 0.104 | 0.067 | 0.037 | 0.000 | 0.719 | 0.774 | -0.056 | 0.000 | 0.799 | 0.824 | -0.025 | 0.019 | 0.154 | 0.258 | -0.104 | 0.000 |
| Tajikistan | 0.238 | 0.204 | 0.033 | 0.014 | 0.468 | 0.553 | -0.085 | 0.000 | 0.640 | 0.692 | -0.051 | 0.001 | 0.120 | 0.162 | -0.042 | 0.000 |
| Thailand | 0.463 | 0.260 | 0.203 | 0.000 | 0.348 | 0.505 | -0.157 | 0.000 | 0.335 | 0.575 | -0.240 | 0.000 | 0.019 | 0.037 | -0.019 | 0.000 |
| Togo | 0.075 | 0.074 | 0.001 | 0.830 | 0.694 | 0.685 | 0.009 | 0.418 | 0.785 | 0.808 | -0.023 | 0.019 | 0.153 | 0.194 | -0.042 | 0.000 |
| Tonga | 0.183 | 0.079 | 0.104 | 0.000 | 0.675 | 0.840 | -0.166 | 0.000 | 0.606 | 0.779 | -0.173 | 0.000 | 0.163 | 0.288 | -0.125 | 0.000 |
| Trinidad and Tobago | 0.237 | 0.168 | 0.069 | 0.000 | 0.370 | 0.531 | -0.161 | 0.000 | 0.607 | 0.682 | -0.075 | 0.001 | 0.021 | 0.036 | -0.015 | 0.026 |
| Tunisia | 0.119 | 0.089 | 0.030 | 0.001 | 0.604 | 0.725 | -0.120 | 0.000 | 0.805 | 0.820 | -0.015 | 0.181 | 0.122 | 0.277 | -0.155 | 0.000 |
| Turkmenistan | 0.392 | 0.458 | -0.066 | 0.000 | 0.337 | 0.340 | -0.003 | 0.849 | 0.522 | 0.436 | 0.086 | 0.000 | 0.011 | 0.012 | -0.001 | 0.861 |
| Turks and Caicos Islands | 0.211 | 0.128 | 0.083 | 0.250 | 0.542 | 0.660 | -0.118 | 0.294 | 0.715 | 0.608 | 0.107 | 0.346 | 0.040 | 0.047 | -0.007 | 0.870 |
| Tuvalu | 0.221 | 0.148 | 0.073 | 0.113 | 0.623 | 0.720 | -0.097 | 0.082 | 0.542 | 0.655 | -0.114 | 0.048 | 0.025 | 0.073 | -0.048 | 0.048 |
| Uganda | 0.138 | 0.106 | 0.032 | 0.002 | 0.605 | 0.704 | -0.099 | 0.000 | 0.674 | 0.702 | -0.028 | 0.049 | 0.097 | 0.155 | -0.058 | 0.000 |
| Ukraine | 0.361 | 0.295 | 0.066 | 0.010 | 0.299 | 0.327 | -0.028 | 0.266 | 0.577 | 0.615 | -0.038 | 0.162 | 0.019 | 0.016 | 0.003 | 0.691 |
| Uruguay | 0.446 | 0.359 | 0.087 | 0.326 | 0.147 | 0.330 | -0.183 | 0.002 | 0.378 | 0.494 | -0.116 | 0.151 | 0.013 | 0.036 | -0.023 | 0.224 |
| Uzbekistan | 0.343 | 0.291 | 0.053 | 0.052 | 0.266 | 0.364 | -0.097 | 0.000 | 0.538 | 0.643 | -0.105 | 0.000 | 0.034 | 0.048 | -0.014 | 0.238 |
| Vietnam | 0.283 | 0.198 | 0.085 | 0.000 | 0.425 | 0.486 | -0.061 | 0.000 | 0.551 | 0.644 | -0.093 | 0.000 | 0.020 | 0.036 | -0.015 | 0.000 |
| Yemen | 0.145 | 0.081 | 0.063 | 0.000 | 0.473 | 0.613 | -0.140 | 0.000 | 0.765 | 0.826 | -0.061 | 0.000 | 0.134 | 0.340 | -0.206 | 0.000 |

| | | | | | | | | | | | | | | | | |
|---|---|---|---|---|---|---|---|---|---|---|---|---|---|---|---|---|
| Zimbabwe | 0.226 | 0.214 | 0.011 | 0.234 | 0.476 | 0.347 | 0.129 | 0.000 | 0.554 | 0.507 | 0.048 | 0.000 | 0.057 | 0.055 | 0.002 | 0.649 |
| Mean | 0.223 | 0.171 | 0.051 | | 0.505 | 0.578 | -0.073 | | 0.617 | 0.655 | -0.038 | | 0.102 | 0.146 | -0.044 | |

*Notes*: Gaps are computed as the difference between richest and poorest quintile averages. A positive value implies richest (dis)advantage while a negative value implies poorer (dis)advantage. The analysis included 86 LMICs and HICs – it excludes Cuba (MICS6), Djibouti, Iraq (MICS3), Jamaica (MICS3) and Qatar due to lack of information on wealth. All averages were estimated using household weights. "Mean" gives the unweighted average of the 86 country-estimates.

Table B9: Share of 1-14y-olds exposed different child disciplinary practices

| Country | Only physical punishment | Only emotional violence | Physical and emotional violence | Observations |
|---|---|---|---|---|
| Afghanistan | 0.018 | 0.012 | 0.061 | 54200 |
| Albania | 0.024 | 0.000 | 0.007 | 2478 |
| Algeria | 0.006 | 0.012 | 0.025 | 42303 |
| Argentina | 0.003 | 0.010 | 0.008 | 22214 |
| Bangladesh | 0.008 | 0.026 | 0.045 | 87808 |
| Barbados | 0.015 | 0.033 | 0.027 | 891 |
| Belarus | 0.001 | 0.008 | 0.003 | 11741 |
| Belize | 0.011 | 0.008 | 0.010 | 6516 |
| Benin | 0.011 | 0.025 | 0.058 | 39976 |
| Bosnia and Herzegovina | 0.006 | 0.006 | 0.005 | 5472 |
| Burkina Faso | 0.007 | 0.045 | 0.055 | 4328 |
| Burundi | 0.011 | 0.037 | 0.076 | 11228 |
| Cambodia | 0.010 | 0.014 | 0.015 | 14196 |
| Cameroon | 0.011 | 0.029 | 0.082 | 11830 |
| Central Africa Rep. | 0.009 | 0.014 | 0.054 | 28347 |
| Chad | 0.016 | 0.015 | 0.034 | 51218 |
| Comoros | 0.012 | 0.022 | 0.029 | 6927 |
| Congo | 0.013 | 0.033 | 0.083 | 15381 |
| Congo Dem. Rep. | 0.023 | 0.026 | 0.088 | 44062 |
| Costa Rica | 0.009 | 0.006 | 0.005 | 9161 |
| Cote d'Ivoire | 0.009 | 0.029 | 0.050 | 24175 |
| Cuba | 0.021 | 0.010 | 0.010 | 13627 |
| Djibouti | 0.022 | 0.012 | 0.026 | 3119 |
| Dominican Rep. | 0.024 | 0.030 | 0.037 | 35779 |
| Egypt | 0.002 | 0.012 | 0.047 | 15664 |
| El Salvador | 0.020 | 0.010 | 0.012 | 8147 |
| Eswatini | 0.016 | 0.030 | 0.064 | 9646 |
| Fiji | 0.006 | 0.004 | 0.006 | 3947 |
| Gabon | 0.010 | 0.015 | 0.033 | 3842 |
| Gambia | 0.010 | 0.016 | 0.030 | 23822 |
| Georgia | 0.006 | 0.020 | 0.020 | 9336 |
| Ghana | 0.010 | 0.032 | 0.074 | 26519 |
| Guinea | 0.012 | 0.028 | 0.030 | 6138 |
| Guinea Bissau | 0.021 | 0.008 | 0.035 | 20765 |
| Guyana | 0.012 | 0.026 | 0.027 | 11026 |
| Haiti | 0.054 | 0.016 | 0.115 | 14524 |
| Honduras | 0.034 | 0.019 | 0.028 | 16307 |
| Iraq | 0.003 | 0.021 | 0.031 | 66664 |
| Jamaica | 0.018 | 0.023 | 0.039 | 8010 |
| Jordan | 0.002 | 0.010 | 0.015 | 18133 |
| Kazakhstan | 0.003 | 0.012 | 0.006 | 21118 |
| Kiribati | 0.009 | 0.002 | 0.015 | 3635 |
| Kosovo | 0.001 | 0.016 | 0.005 | 5118 |
| Kyrgyzstan | 0.006 | 0.008 | 0.010 | 13592 |
| Lao People's Dem. Rep. | 0.007 | 0.032 | 0.029 | 41001 |
| Lesotho | 0.037 | 0.013 | 0.044 | 6653 |
| Liberia | 0.008 | 0.018 | 0.049 | 11382 |
| Macedonia | 0.007 | 0.008 | 0.012 | 7660 |
| Madagascar | 0.010 | 0.018 | 0.050 | 19842 |
| Malawi | 0.013 | 0.020 | 0.030 | 46275 |
| Mauritania | 0.010 | 0.014 | 0.021 | 16393 |
| Moldova | 0.002 | 0.011 | 0.002 | 3119 |
| Mongolia | 0.010 | 0.017 | 0.010 | 24677 |
| Montenegro | 0.005 | 0.025 | 0.018 | 4557 |
| Mozambique | 0.010 | 0.072 | 0.034 | 5299 |
| Myanmar | 0.007 | 0.054 | 0.045 | 7778 |
| Nepal | 0.005 | 0.016 | 0.023 | 19241 |
| Niger | 0.008 | 0.018 | 0.036 | 8964 |
| Nigeria | 0.012 | 0.021 | 0.053 | 81637 |
| Panama | 0.010 | 0.009 | 0.012 | 6316 |
| Paraguay | 0.013 | 0.010 | 0.006 | 4652 |
| Philippines | 0.005 | 0.005 | 0.003 | 17416 |
| Qatar | 0.007 | 0.004 | 0.003 | 2777 |
| St. Lucia | 0.004 | 0.039 | 0.021 | 590 |
| Samoa | 0.013 | 0.003 | 0.015 | 3994 |
| Sao Tome and Principe | 0.043 | 0.023 | 0.096 | 5438 |
| Serbia | 0.008 | 0.008 | 0.006 | 12718 |
| Sierra Leone | 0.015 | 0.030 | 0.069 | 33772 |
| Palestine | 0.002 | 0.009 | 0.013 | 25804 |
| Sudan | 0.015 | 0.025 | 0.036 | 11289 |

| | | | | |
|---|---|---|---|---|
| Suriname | 0.005 | 0.028 | 0.029 | 13359 |
| Syria | 0.006 | 0.010 | 0.037 | 12766 |
| Tajikistan | 0.004 | 0.007 | 0.016 | 11052 |
| Thailand | 0.005 | 0.005 | 0.004 | 61950 |
| Togo | 0.011 | 0.025 | 0.061 | 23449 |
| Tonga | 0.011 | 0.005 | 0.010 | 2336 |
| Trinidad and Tobago | 0.008 | 0.021 | 0.012 | 8024 |
| Tunisia | 0.004 | 0.006 | 0.014 | 15140 |
| Turkmenistan | 0.002 | 0.001 | 0.001 | 9551 |
| Turks and Caicos Islands | 0.005 | 0.020 | 0.003 | 635 |
| Tuvalu | 0.006 | 0.000 | 0.008 | 767 |
| Uganda | 0.021 | 0.024 | 0.055 | 14380 |
| Ukraine | 0.001 | 0.002 | 0.002 | 7315 |
| Uruguay | 0.001 | 0.012 | 0.006 | 2037 |
| Uzbekistan | 0.002 | 0.013 | 0.005 | 3735 |
| Vietnam | 0.008 | 0.016 | 0.019 | 23252 |
| Yemen | 0.004 | 0.023 | 0.030 | 29892 |
| Zimbabwe | 0.018 | 0.039 | 0.027 | 22154 |
| Mean/total | 0.011 | 0.018 | 0.030 | 1543968 |

*Notes*: The table gives average share of children aged 1-14y exposed to child discipline, by type of violence and country using household weights. The analysis included 88 LMICs and HICs. "Mean" gives the unweighted average of the 88 country-estimates.

**Figure B1: Share of 1-14y-olds exposed to severe physical violence by country. Online supplementary appendix B Table B1 presents the details of the estimates by country.**

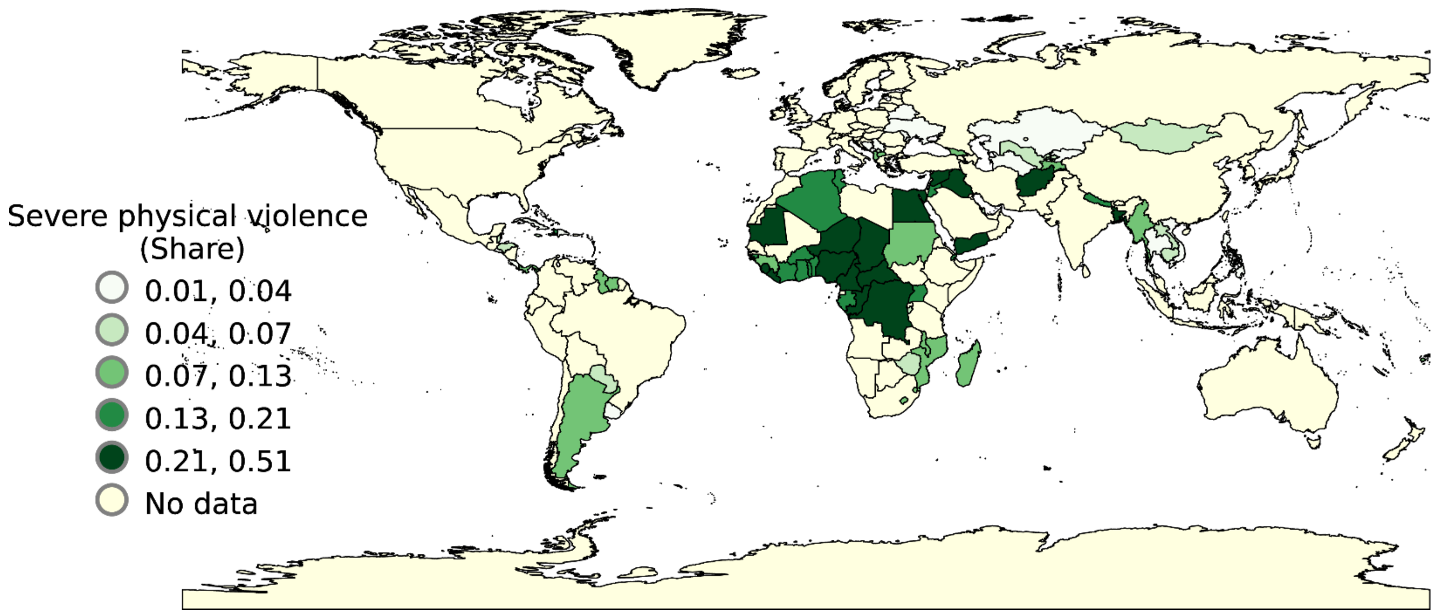

**Figure B2: Trends in severe physical voilence by country and survey wave. The x-axis represents cumulative share of each country's sample population within a given wave, ordered from highest to lowest values of severe physical violence. Countries with at least three survey rounds are labelled. See online supplementary appendix B Figure B3 for estimates for the 54 countries.**

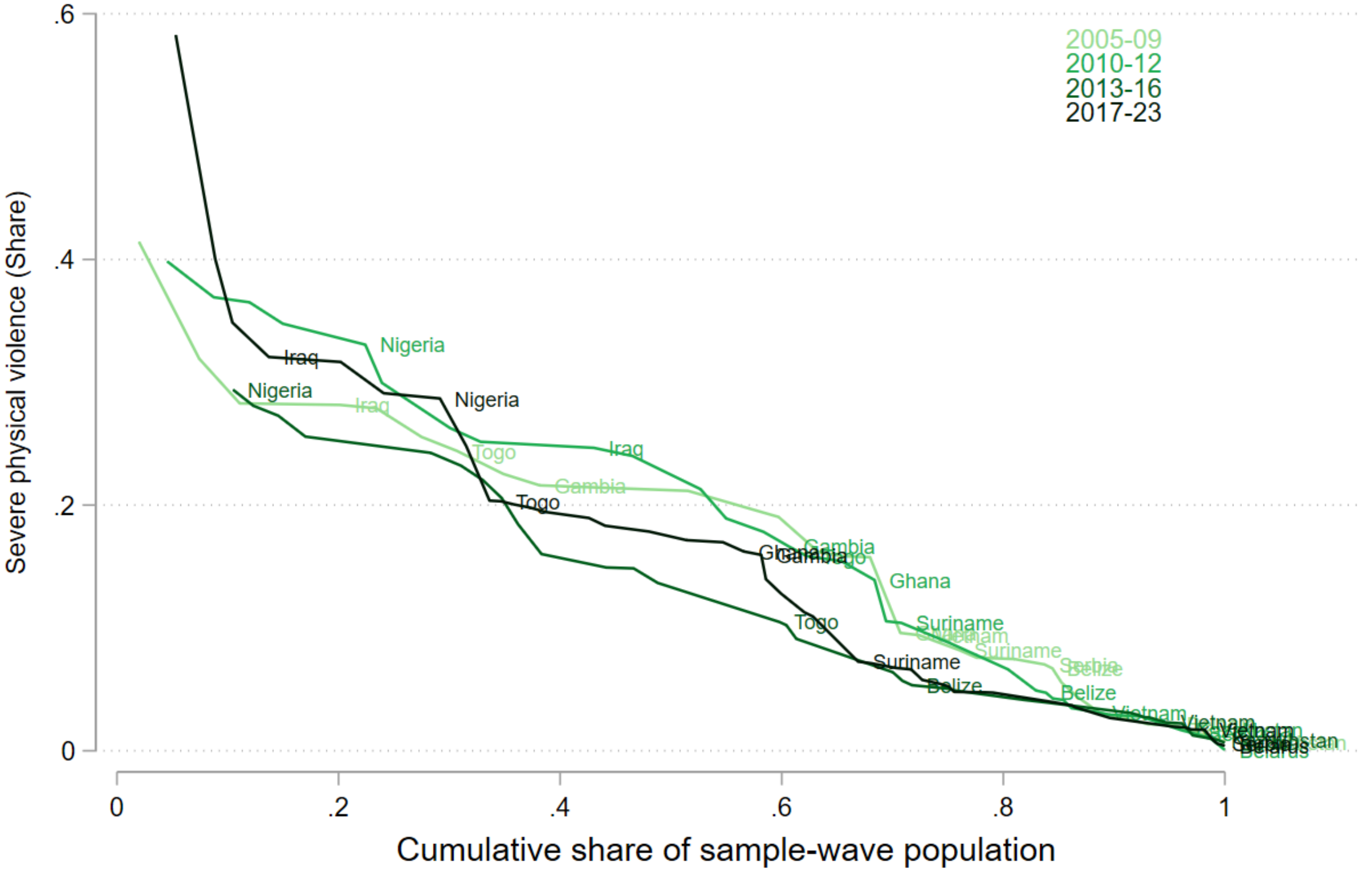

**Figure B3: Evolution of types of child discipline by country. Estimates are shown for all 54 countries with at least two survey waves.**

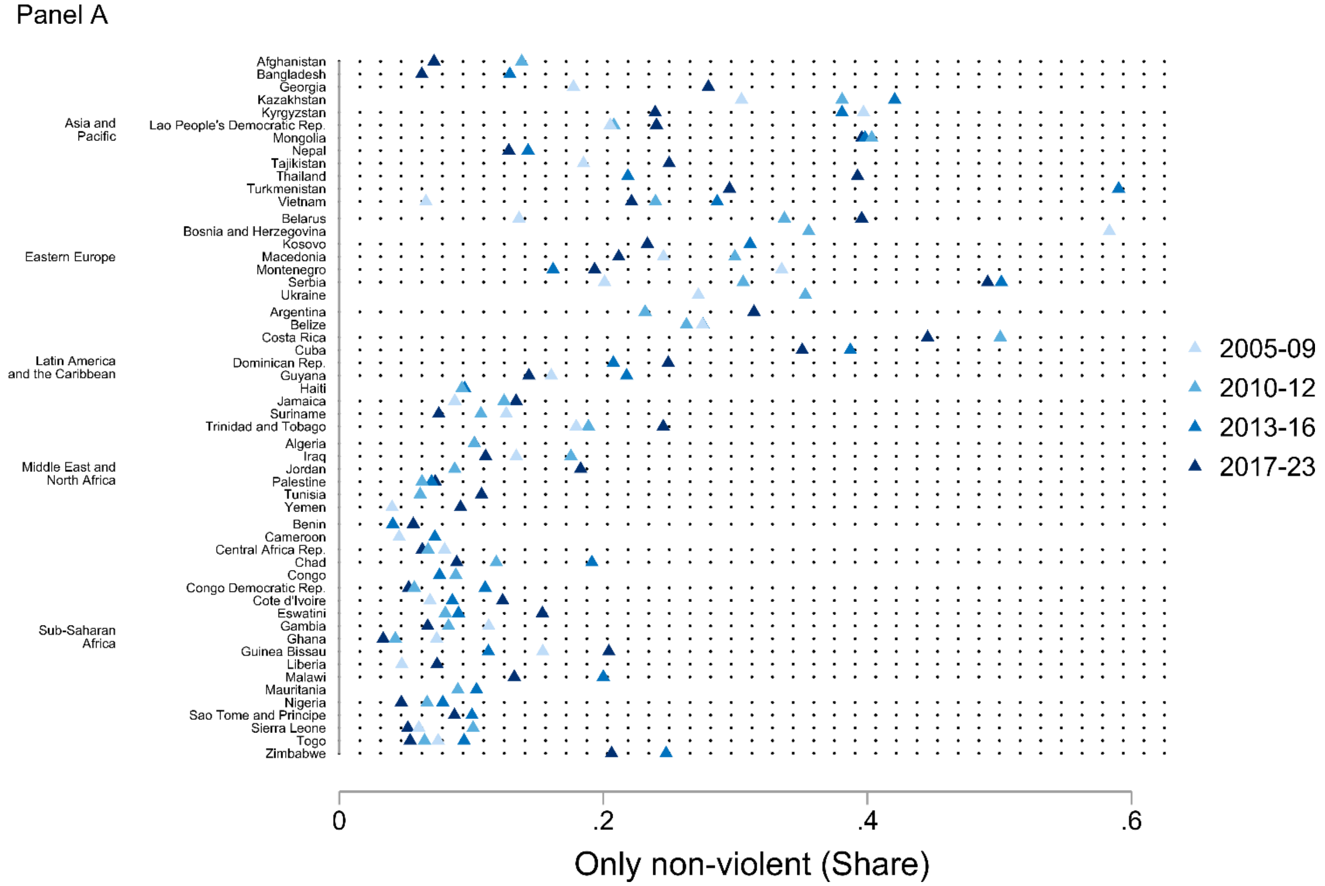

Panel B

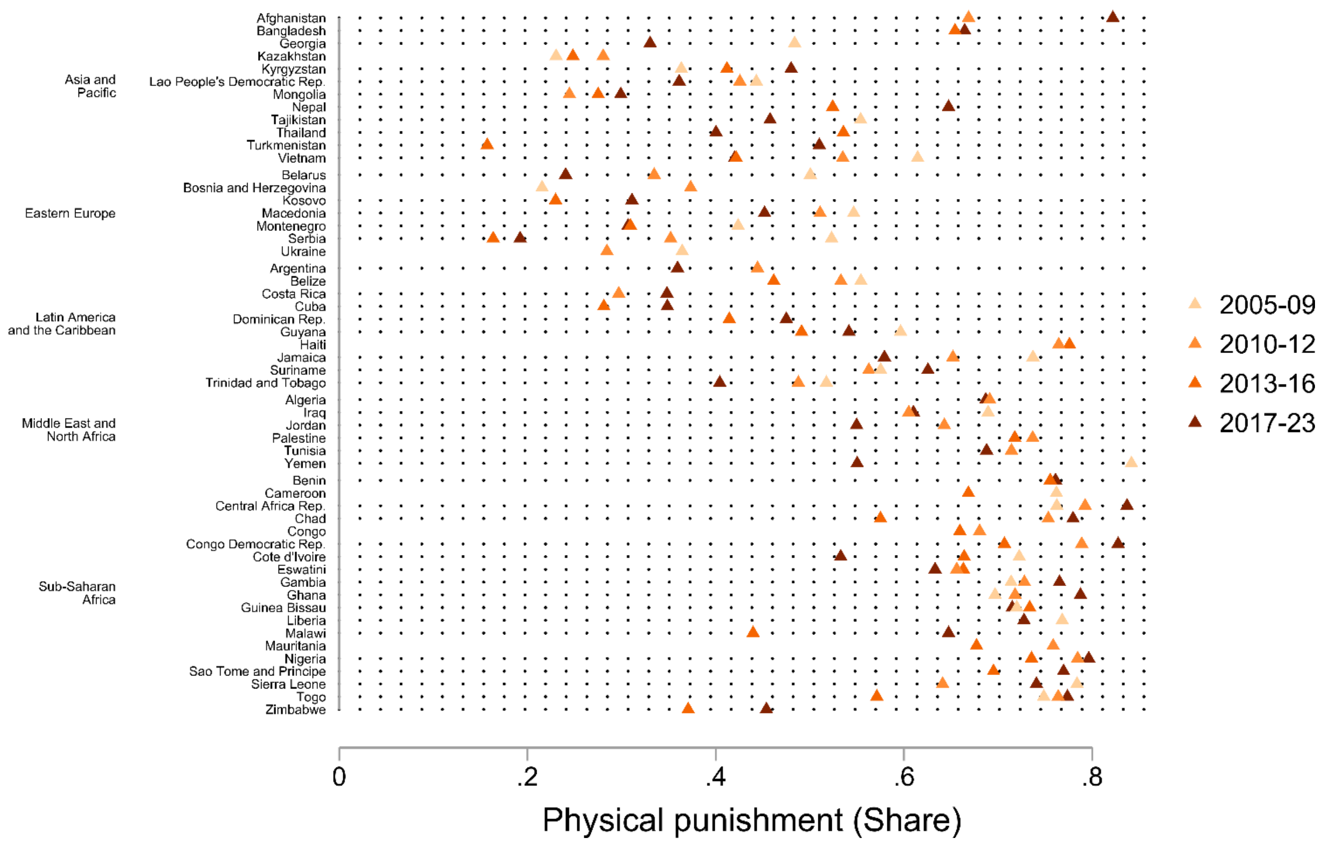

Panel C

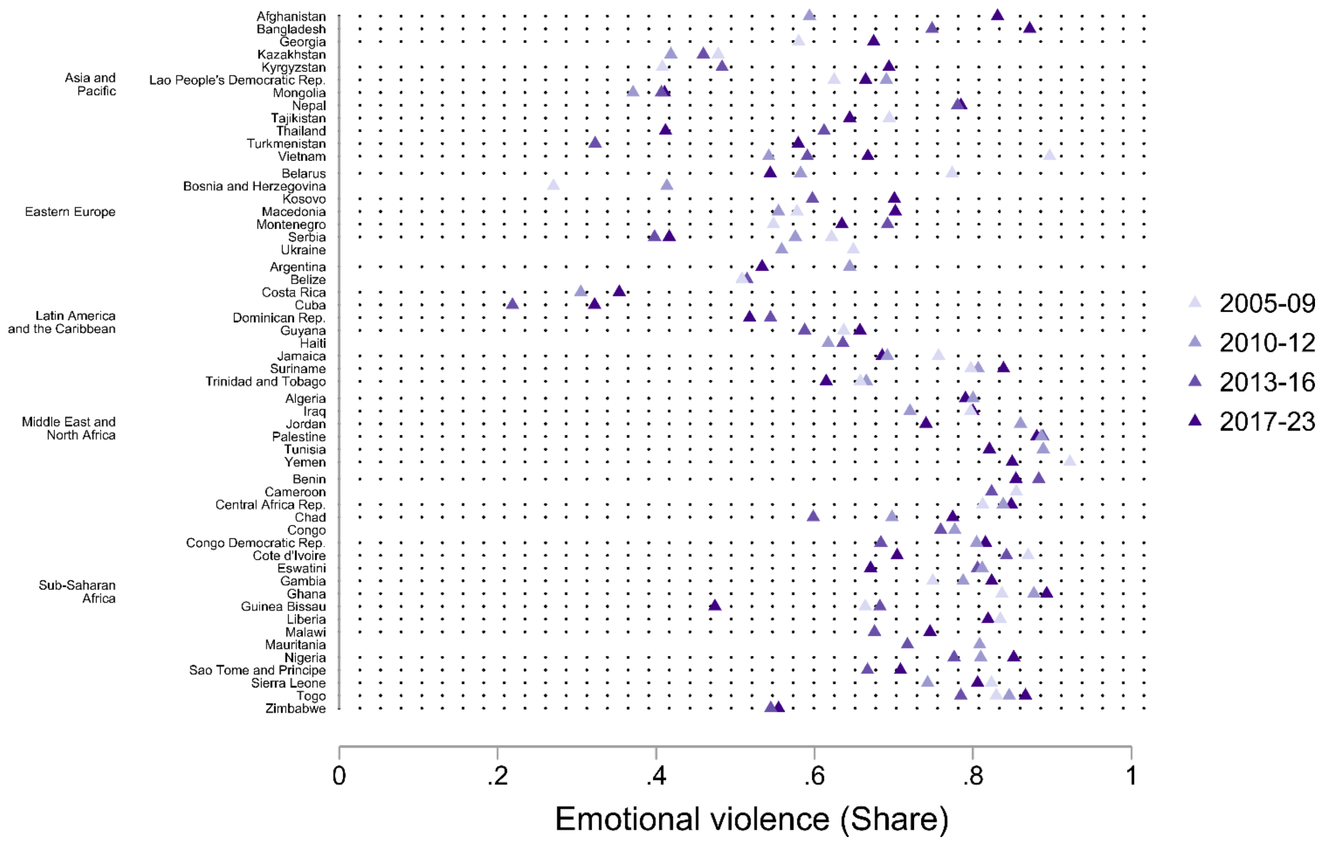

Panel D

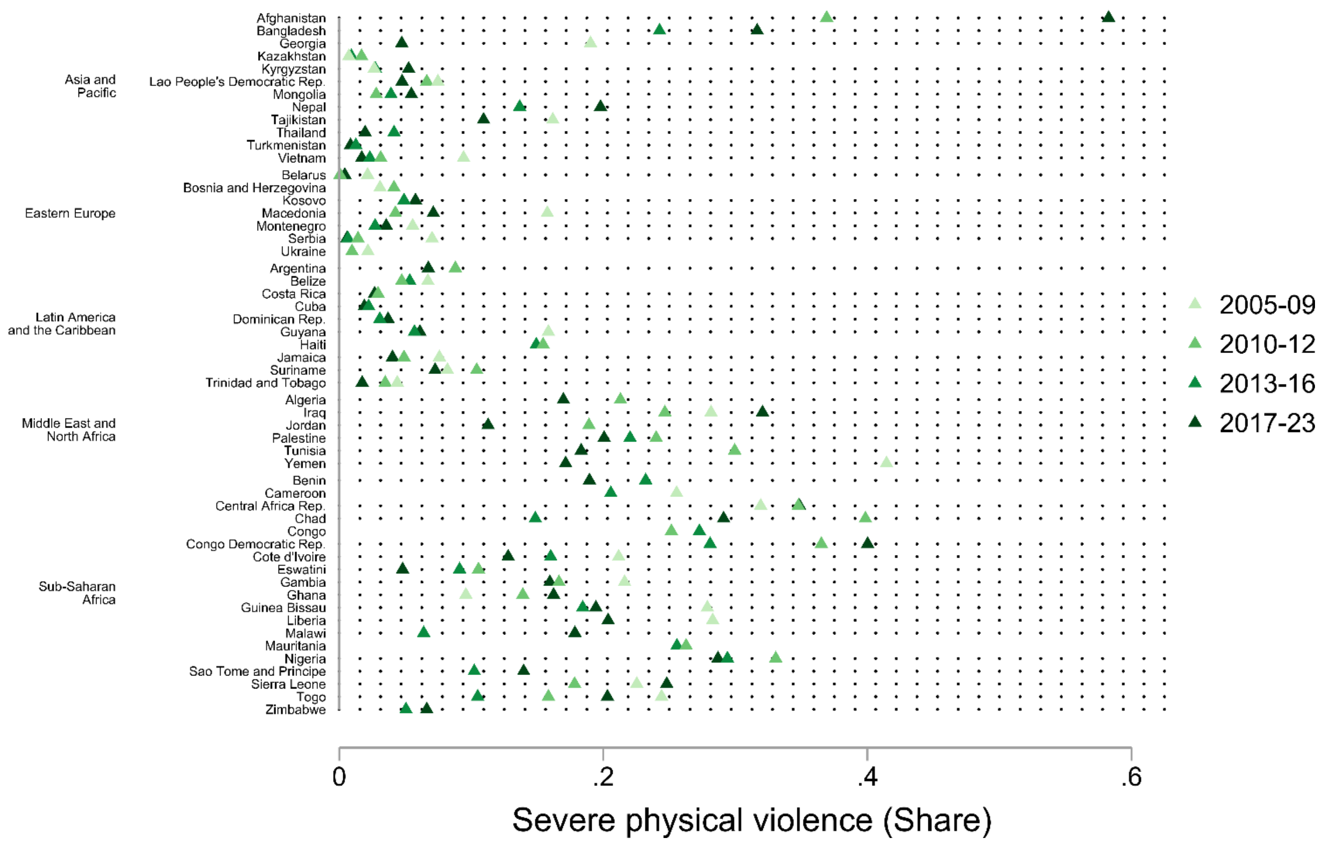

Asia and Pacific
Afghanistan
Bangladesh
Georgia
Kazakhstan
Kyrgyzstan
Lao People's Democratic Rep.
Mongolia
Nepal
Tajikistan
Thailand
Turkmenistan
Vietnam
Eastern Europe
Belarus
Bosnia and Herzegovina
Kosovo
Macedonia
Montenegro
Serbia
Ukraine
Latin America and the Caribbean
Argentina
Belize
Costa Rica
Cuba
Dominican Rep.
Guyana
Haiti
Jamaica
Suriname
Trinidad and Tobago
Middle East and North Africa
Algeria
Iraq
Jordan
Palestine
Tunisia
Yemen
Sub-Saharan Africa
Benin
Cameroon
Central Africa Rep.
Chad
Congo
Congo Democratic Rep.
Cote d'Ivoire
Eswatini
Gambia
Ghana
Guinea Bissau
Liberia
Malawi
Mauritania
Nigeria
Sao Tome and Principe
Sierra Leone
Togo
Zimbabwe
0
.2
.4
.6
Severe physical violence (Share)
2005-09
2010-12
2013-16
2017-23

**Figure B4: Trends in each type of discipline by country and age groups. The x-axis represents the cumulative share of each country's sample population within a given wave, ordered from highest to lowest values of the discipline type. Countries with at least three survey rounds are labelled. See online supplementary appendix B Figure B5 for estimates for the 54 countries.**

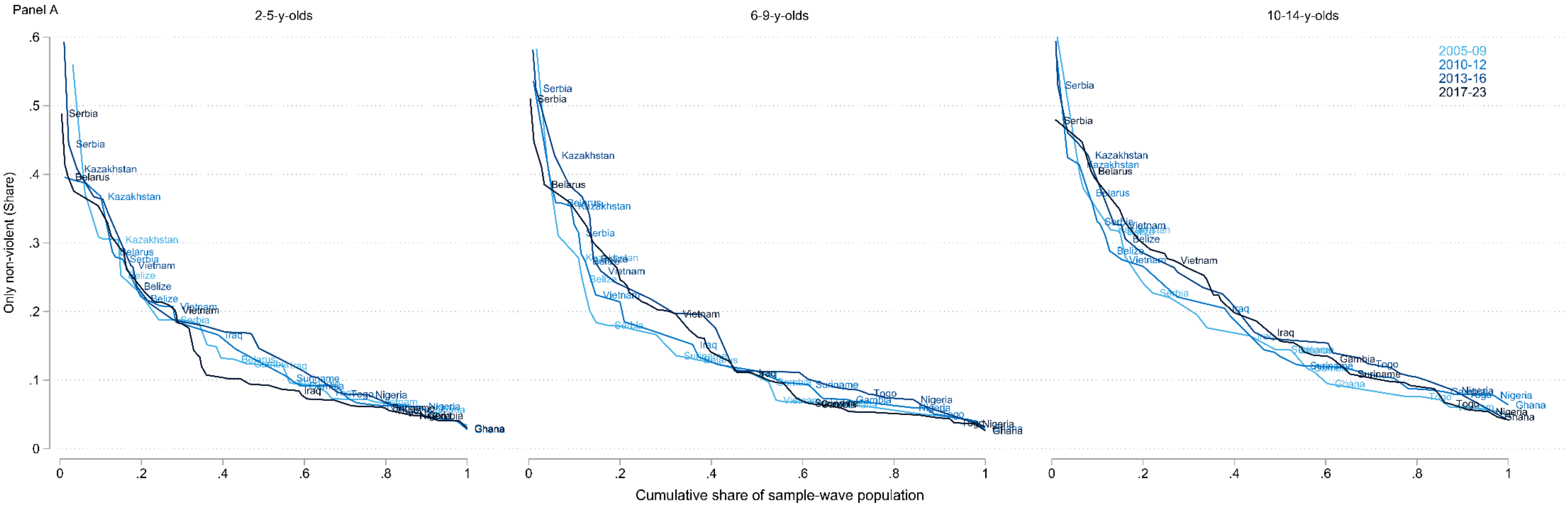

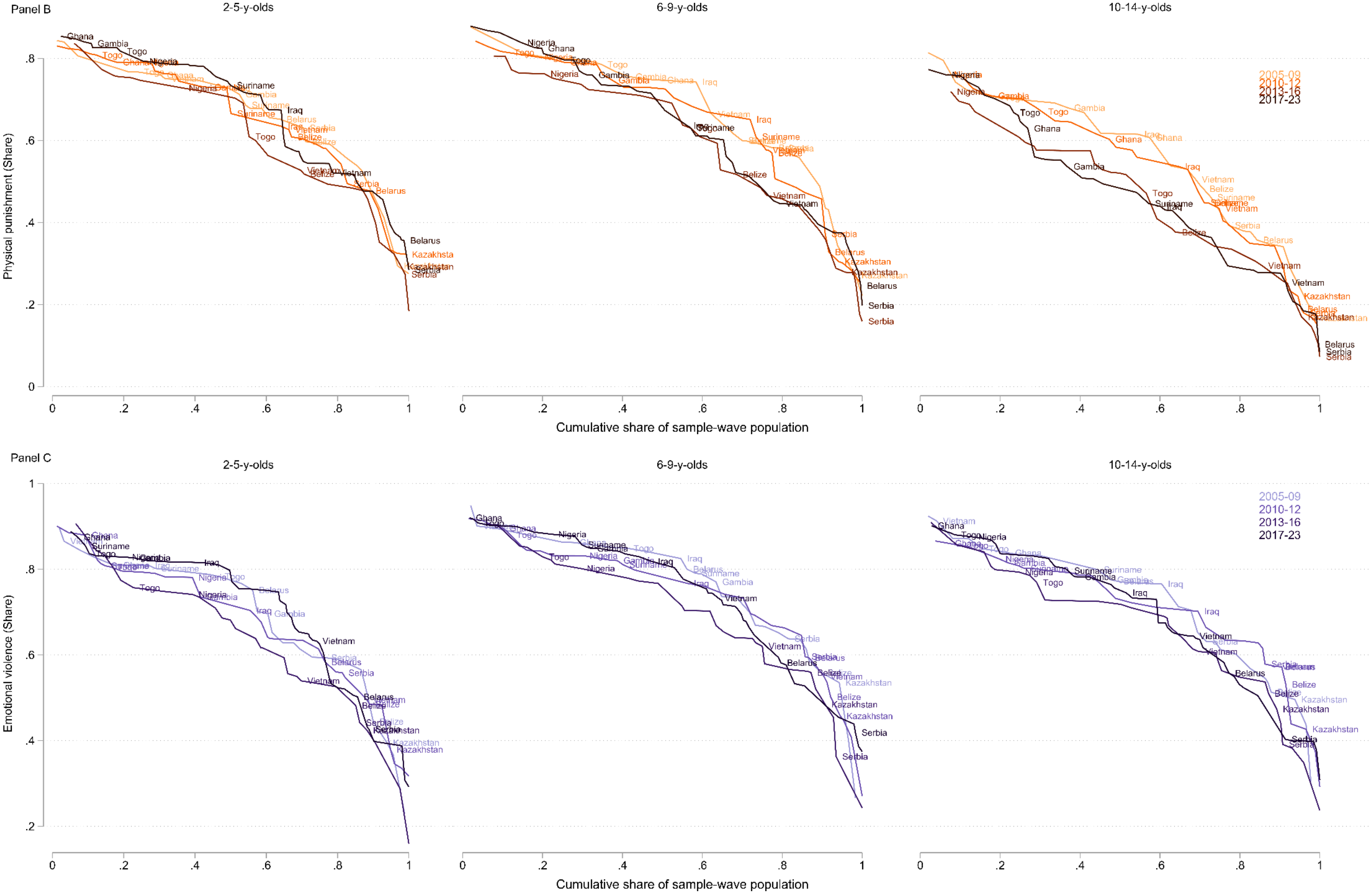
Panel B
2-5-y-olds
6-9-y-olds
10-14-y-olds
Physical punishment (Share)
Cumulative share of sample-wave population
2005-09
2010-12
2013-16
2017-23
Panel C
2-5-y-olds
6-9-y-olds
10-14-y-olds
Emotional violence (Share)
Cumulative share of sample-wave population
2005-09
2010-12
2013-16
2017-23

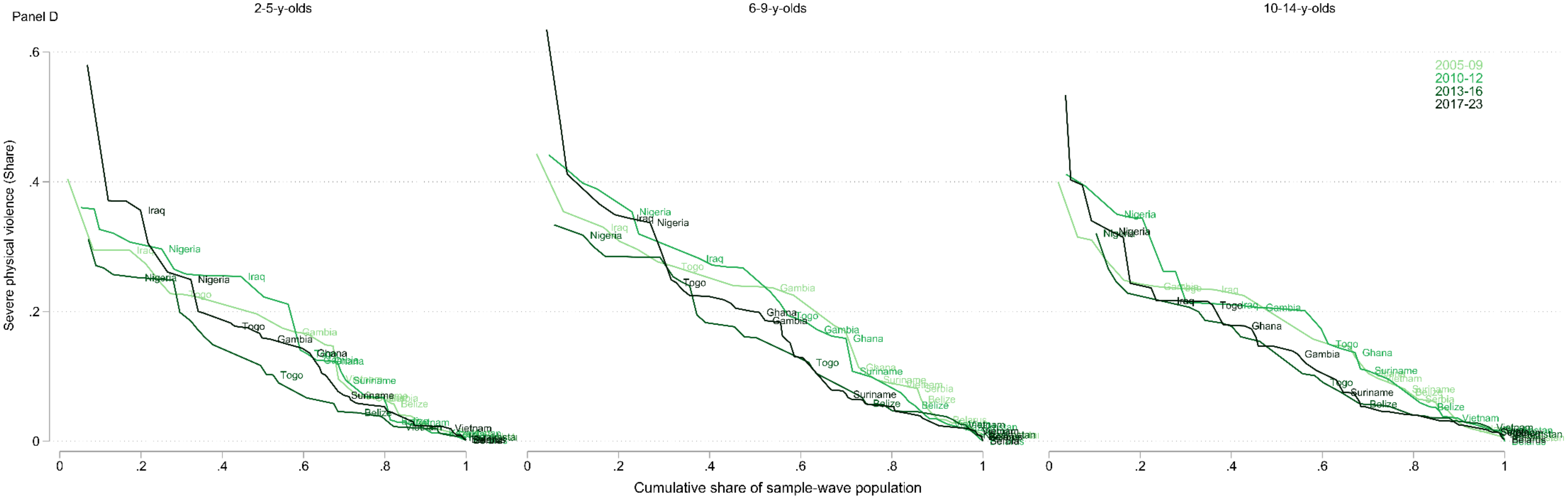

Panel D
2-5-y-olds
6-9-y-olds
10-14-y-olds
2005-09
2010-12
2013-16
2017-23
Severe physical violence (Share)
Cumulative share of sample-wave population

**Figure B5:** Evolution of each type of discipline by country and age group. Estimates are shown for all 54 countries with at least two survey waves.

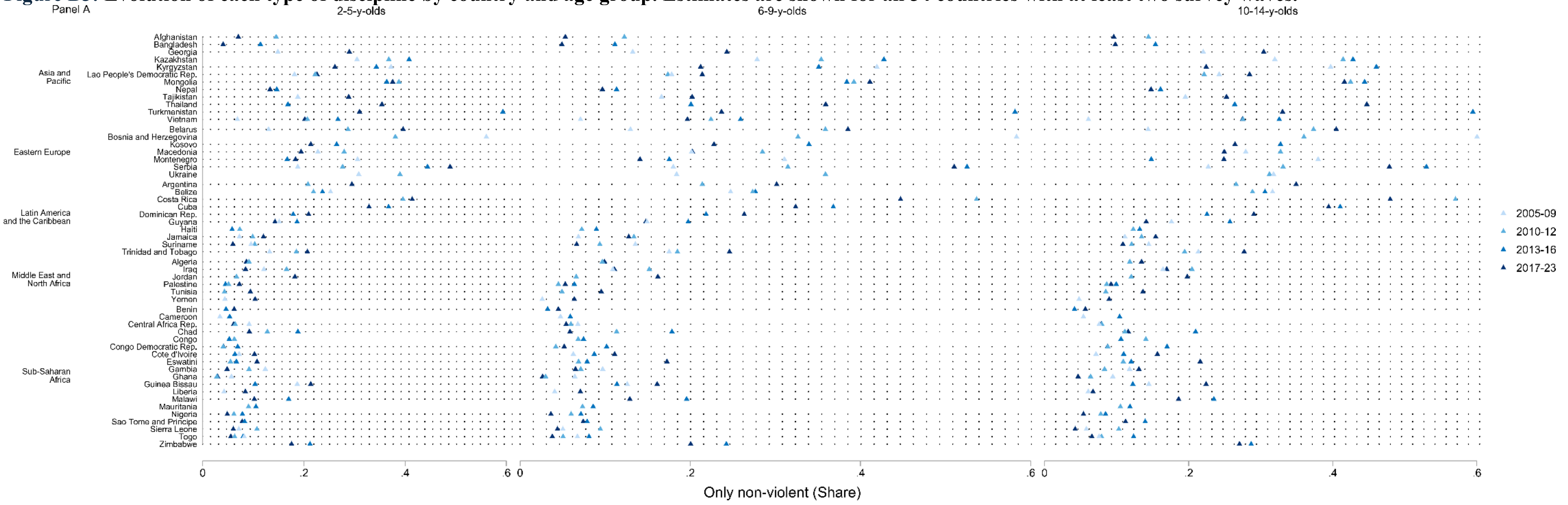

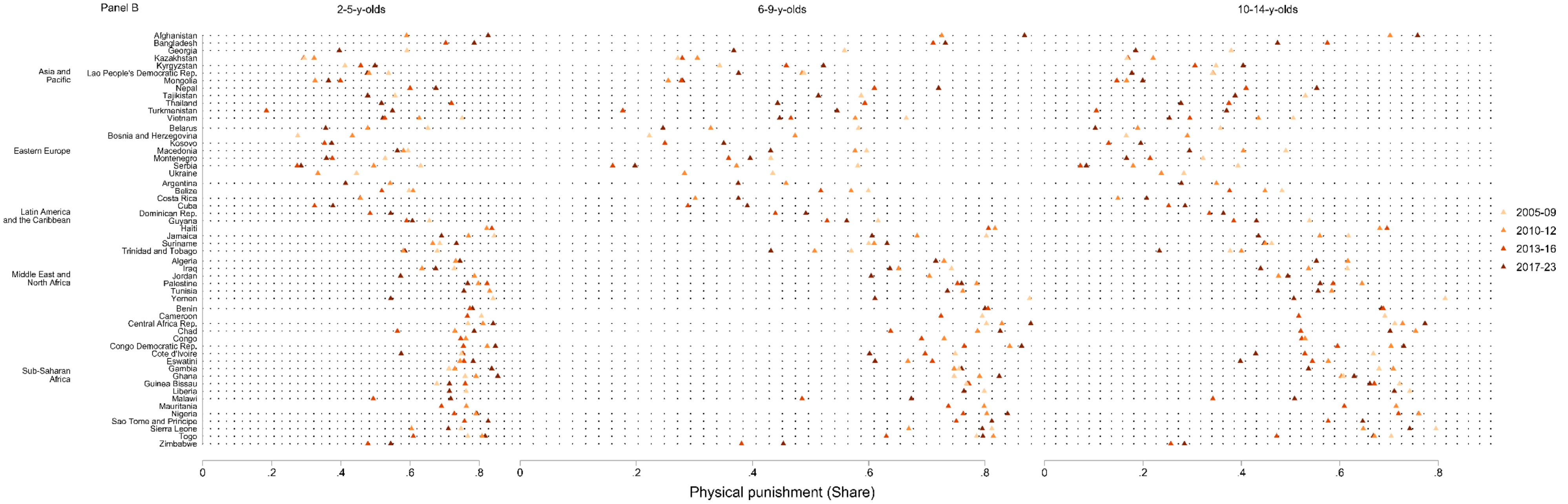

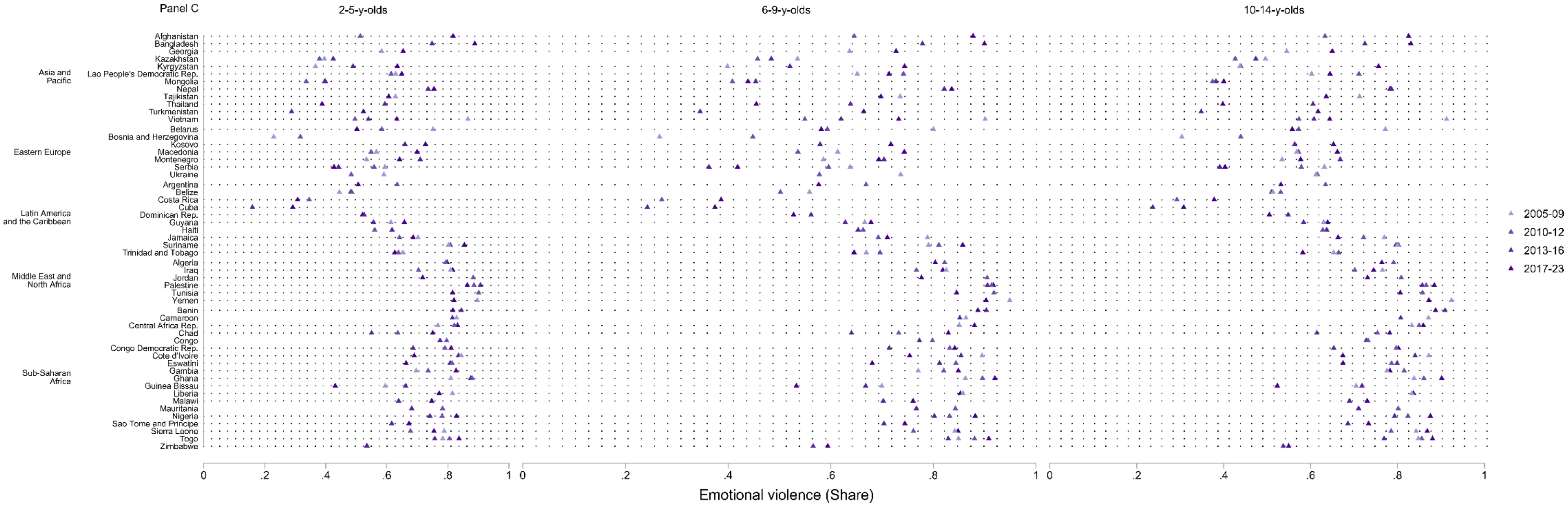

Panel C
2-5-y-olds
6-9-y-olds
10-14-y-olds
Asia and Pacific
Afghanistan
Bangladesh
Georgia
Kazakhstan
Kyrgyzstan
Lao People's Democratic Rep.
Mongolia
Nepal
Tajikistan
Thailand
Turkmenistan
Vietnam
Eastern Europe
Belarus
Bosnia and Herzegovina
Kosovo
Macedonia
Montenegro
Serbia
Ukraine
Latin America and the Caribbean
Argentina
Belize
Costa Rica
Cuba
Dominican Rep.
Guyana
Haiti
Jamaica
Suriname
Trinidad and Tobago
Middle East and North Africa
Algeria
Iraq
Jordan
Palestine
Tunisia
Yemen
Sub-Saharan Africa
Benin
Cameroon
Central Africa Rep.
Chad
Congo
Congo Democratic Rep.
Cote d'Ivoire
Eswatini
Gambia
Ghana
Guinea Bissau
Liberia
Malawi
Mauritania
Nigeria
Sao Tome and Principe
Sierra Leone
Togo
Zimbabwe
0 .2 .4 .6 .8 1
Emotional violence (Share)
2005-09
2010-12
2013-16
2017-23

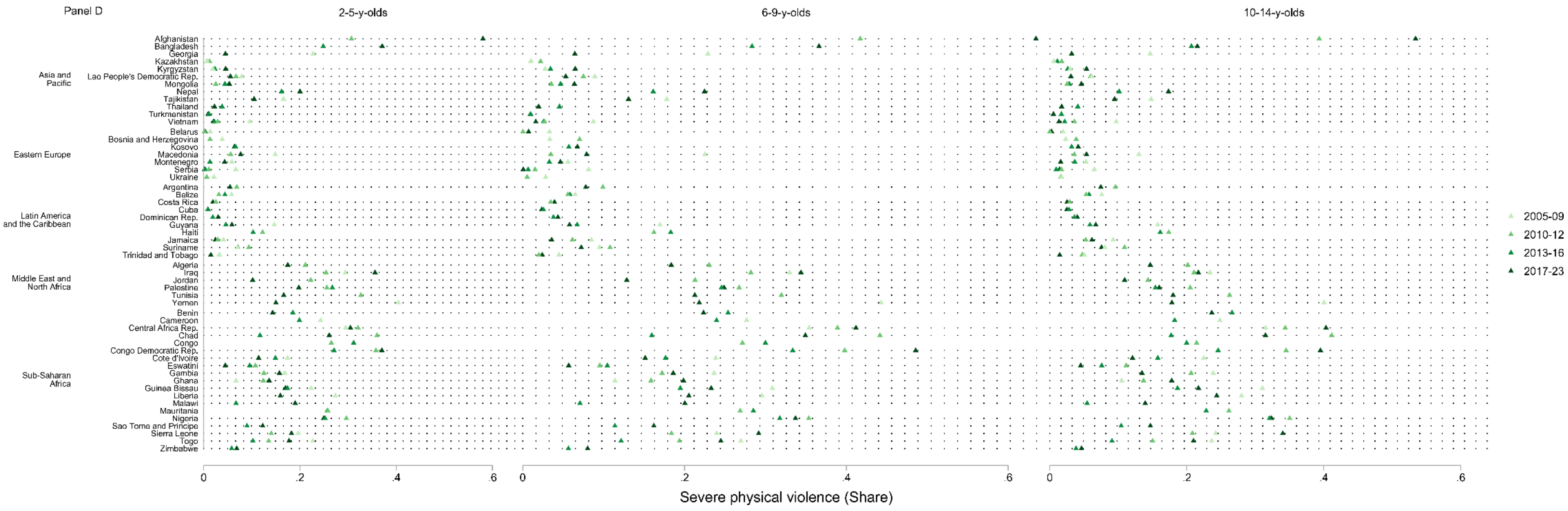

Panel D
2-5-y-olds
6-9-y-olds
10-14-y-olds
Asia and Pacific
Afghanistan
Bangladesh
Georgia
Kazakhstan
Kyrgyzstan
Lao People's Democratic Rep.
Mongolia
Nepal
Tajikistan
Thailand
Turkmenistan
Vietnam
Eastern Europe
Belarus
Bosnia and Herzegovina
Kosovo
Macedonia
Montenegro
Serbia
Ukraine
Latin America and the Caribbean
Argentina
Belize
Costa Rica
Cuba
Dominican Rep.
Guyana
Haiti
Jamaica
Suriname
Trinidad and Tobago
Middle East and North Africa
Algeria
Iraq
Jordan
Palestine
Tunisia
Yemen
Sub-Saharan Africa
Benin
Cameroon
Central Africa Rep.
Chad
Congo
Congo Democratic Rep.
Cote d'Ivoire
Eswatini
Gambia
Ghana
Guinea Bissau
Liberia
Malawi
Mauritania
Nigeria
Sao Tome and Principe
Sierra Leone
Togo
Zimbabwe
0
.2
.4
.6
Severe physical violence (Share)
2005-09
2010-12
2013-16
2017-23

**Figure B6: Share of 1-14y-olds exposed to "only physical punishment"; "only emotional violence"; and "both physical and emotional violence" by country. Online supplementary appendix B Table B9 presents the details of the estimates by country.**

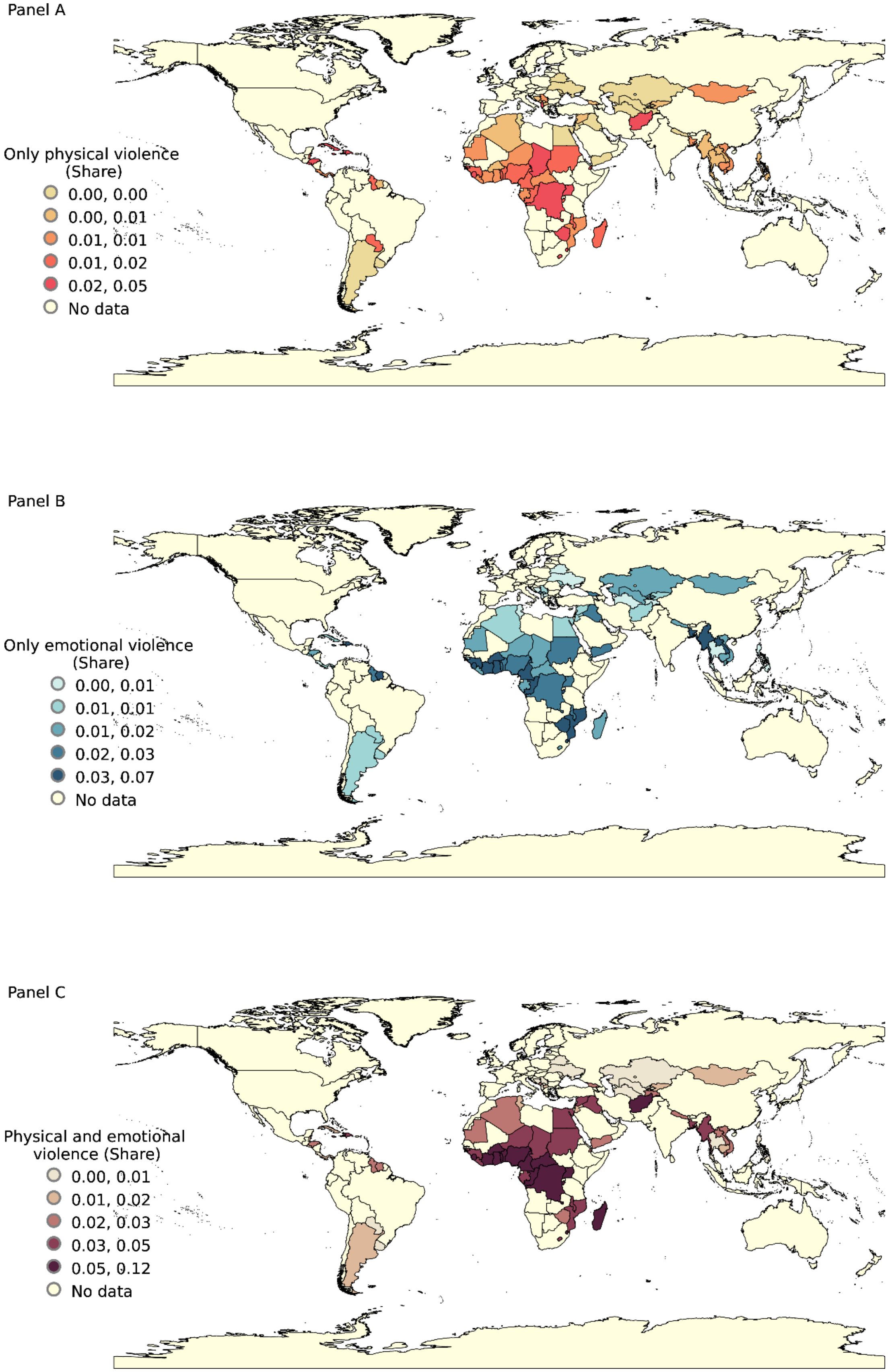

### Online supplementary appendix C: Additional child discipline measures

**Figure C1: Share of 1-14y-olds exposed to “non-violent and violent” and “neither violent nor non-violent” discipline by country.**

Panel A

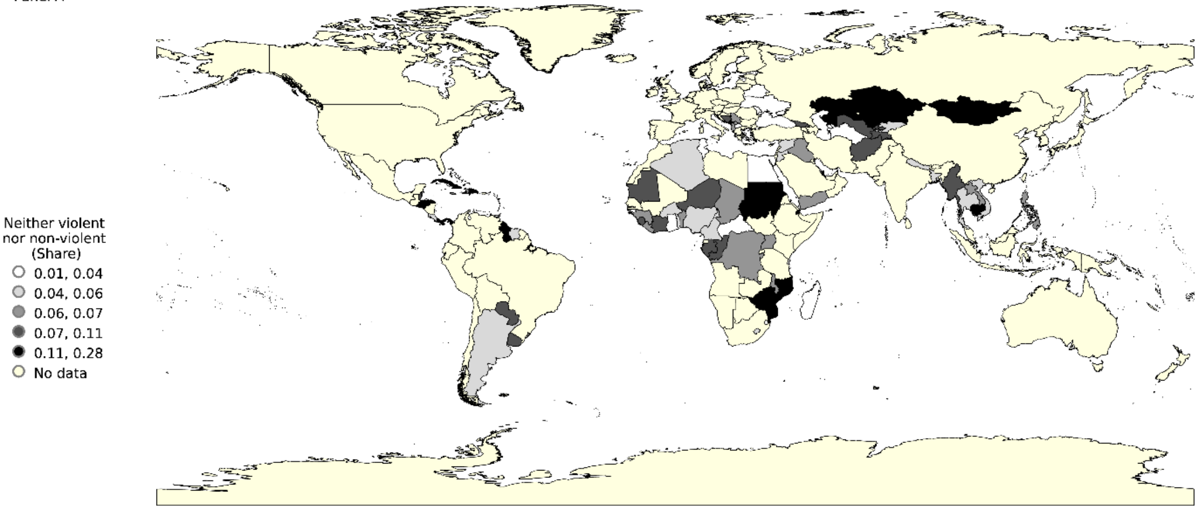

Panel B

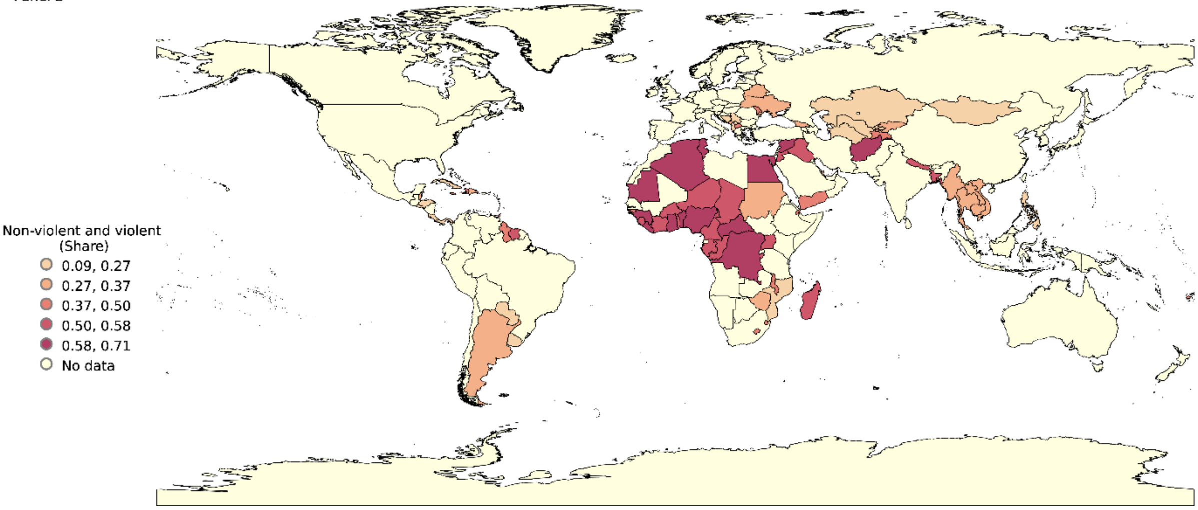
